# Fine-mapping across diverse ancestries drives the discovery of putative causal variants underlying human complex traits and diseases

**DOI:** 10.1101/2023.01.07.23284293

**Authors:** Kai Yuan, Ryan J. Longchamps, Antonio F. Pardiñas, Mingrui Yu, Tzu-Ting Chen, Shu-Chin Lin, Yu Chen, Max Lam, Ruize Liu, Yan Xia, Zhenglin Guo, Wenzhao Shi, Chengguo Shen, The Schizophrenia Workgroup of Psychiatric Genomics Consortium, Mark J. Daly, Benjamin M. Neale, Yen-Chen A. Feng, Yen-Feng Lin, Chia-Yen Chen, Michael O’Donovan, Tian Ge, Hailiang Huang

**Author notes:** A list of members is available in the supplementary information (The Schizophrenia Workgroup of Psychiatric Genomics Consortium). These authors jointly supervised this work. (T.G.); (H.H.).

## Abstract

Genome-wide association studies (GWAS) of human complex traits or diseases often implicate genetic loci that span hundreds or thousands of genetic variants, many of which have similar statistical significance. While statistical fine-mapping in individuals of European ancestries has made important discoveries, cross-population fine-mapping has the potential to improve power and resolution by capitalizing on the genomic diversity across ancestries. Here we present SuSiEx, an accurate and computationally efficient method for cross-population fine-mapping, which builds on the single-population fine-mapping framework, Sum of Single Effects (SuSiE). SuSiEx integrates data from an arbitrary number of ancestries, explicitly models population-specific allele frequencies and LD patterns, accounts for multiple causal variants in a genomic region, and can be applied to GWAS summary statistics. We comprehensively evaluated SuSiEx using simulations, a range of quantitative traits measured in both UK Biobank and Taiwan Biobank, and schizophrenia GWAS across East Asian and European ancestries. In all evaluations, SuSiEx fine-mapped more association signals, produced smaller credible sets and higher posterior inclusion probability (PIP) for putative causal variants, and captured population-specific causal variants.

## INTRODUCTION

Genome-wide association studies (GWAS) of human complex traits or diseases often implicate genetic loci that span hundreds or thousands of genetic variants, many of which have similar statistical significance. These loci may contain one or a handful of causal variants, while the associations of other variants are driven by their linkage disequilibrium (LD) with the causal variant(s). Statistical fine-mapping refines a GWAS locus to a smaller set of likely causal variants to facilitate interpretation and computational and experimental functional studies. Fine-mapping studies in samples of European ancestries have made important advances, with some disease-associated loci resolved to single-variant resolution^1–3^. Since non-causal variants tagging causal signals have marginally different effects across populations due to differences in LD patterns, cross-population fine-mapping, which integrates data from multiple populations and capitalizes on the genomic diversity across ancestries (e.g., smaller LD blocks in African populations), holds the promise to further improve fine-mapping resolution.

Cross-population fine-mapping analysis can be broadly classified into three categories, namely the meta-analysis-based methods, the single-population combining methods, and Bayesian statistical methods (Figure 1). The meta-analysis-based method applies single-population fine-mapping methods to meta-analyzed GWAS summary statistics and LD matrices, and has been widely used in the field^4, 5^. This approach, however, assumes no heterogeneity in effect sizes and LD patterns across populations, which is often not true and may lead to false positives and miscalibration of the inferred probability of a variant being causal^6^. The single-population combining method analyzes data from each population independently and subsequently integrates single-population fine-mapping results. While conducive to identifying population-specific causal variants^7^, this approach fails to leverage the increased sample size, potential genetic correlations and LD diversity across populations to facilitate loci discovery and improve fine-mapping resolution, and may be sensitive to the choice of methods that combine population-specific results. Bayesian methods^8, 9^ provide a principled way to fine-map causal variants across populations and have been employed in the analyses of several complex traits or diseases^8–12^. That said, current cross-population Bayesian fine-mapping methods often suffer from inflated false positive rates, poor computational scalability, and inability to distinguish multiple causal signals in the same genomic locus, impeding their applications to emerging biobank-scale datasets of diverse ancestries.

**Figure 1:**
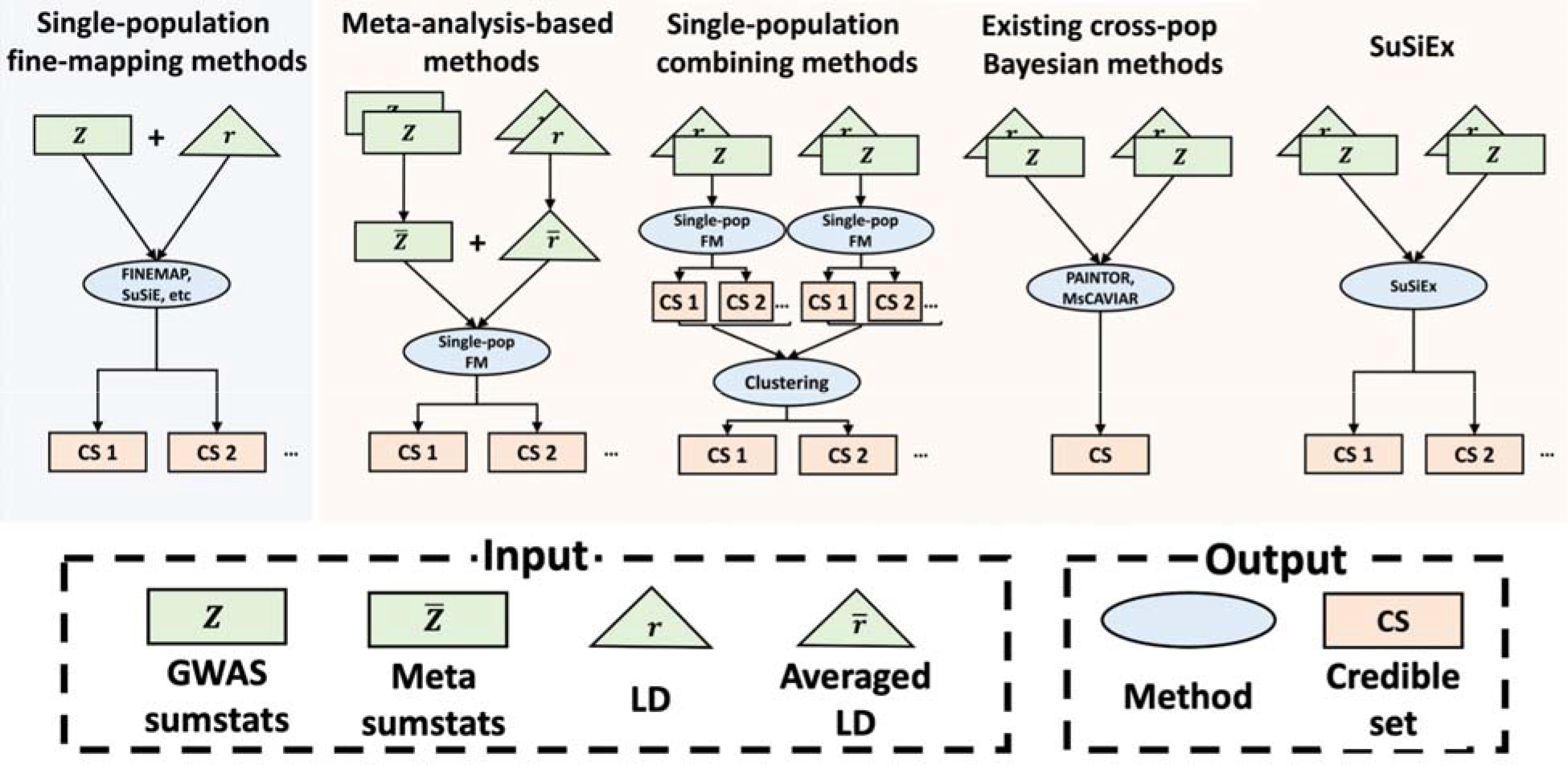
Overview of fine-mapping methods. An illustration of the inputs and outputs for single-population and cross-population fine-mapping methods, the latter of which includes meta-analysis-based methods, single-population combining methods, previously published Bayesian fine-mapping methods as well as SuSiEx. “CS 1” and “CS 2” represent the output of fine-mapping methods that can separate distinct causal signals into different credible sets, while “CS” represents a single credible set output that contains all causal variants as featured in some fine-mapping methods.

Recently, Wang *et al.* proposed a single-population fine-mapping method, SUm of SIngle Effects (SuSiE)^13^, which improved the calibration, computational efficiency and interpretation of statistical fine-mapping. Here, we extend the SuSiE model to a cross-population fine-mapping method, SuSiEx, which integrates multiple population-specific GWAS summary statistics and LD reference panels to enable more powerful and accurate fine-mapping. We evaluated the calibration, power, resolution and computational scalability of SuSiEx along with alternative fine-mapping methods via extensive simulations. We further used SuSiEx to fine-map 25 quantitative traits shared between the UK Biobank^14^ and Taiwan Biobank^15^, and to fine-map schizophrenia genetic risk loci across European and East Asian ancestries.

## RESULTS

### Overview of SuSiEx

SuSiEx extends the single-population fine-mapping model, SuSiE^13^, by integrating population-specific GWAS summary statistics and LD reference panels from multiple populations. In SuSiE, the genetic influence on a trait or disease within a genomic locus is modeled as the summation of several distinct effects, each contributed by a single causal variant, which naturally allows for the modeling of multiple association signals and assigns each inferred putative causal variant to a credible set with a posterior inclusion probability (PIP) (Figure 1). Building on this framework, SuSiEx couples each single effect by assuming that the causal variants are shared across populations (i.e., we report a single PIP rather than population-specific PIPs for each variant in a credible set), while allowing them to have varying effect sizes (including null effects) across ancestries (Supplementary Note). In addition, SuSiEx allows for a variant to be missing in an ancestry (e.g., due to its low allele frequency), in which case the ancestry does not contribute to the PIP estimate. Similar to SuSiE, SuSiEx builds on the Bayesian variable selection in regression^16, 17^ and applies the iterative Bayesian stepwise selection^13^ to model fitting. Once converged, for each inferred credible set, SuSiEx further estimates its causal probability for each population. This probability reflects the *current* evidence of the credible set having a causal effect in the respective population. A small probability, akin to null results from GWAS, does not preclude the credible set to become causal when the sample size grows. Additional modeling and computational details for SuSiEx are discussed in Methods and Supplementary Note.

Compared with the meta-analysis-based fine-mapping methods^4, 5^, SuSiEx explicitly models population-specific GWAS summary statistics and LD patterns (Figure 1; Extended Data Figure 1a), which is expected to improve fine-mapping resolution and more accurately control the false positive rate, while allowing for heterogeneous effect sizes and capturing population-specific causal variants (Extended Data Figure 1c). Compared with the method that combines single-population fine-mapping results^7^, SuSiEx leverages the sample size, genetic correlation and LD diversity across ancestries to improve the resolution of fine-mapping, especially for loci that are under-powered to fine-map in individual datasets (Figure 1; Extended Data Figure 1b). Compared with other Bayesian cross-population fine-mapping methods such as PAINTOR^9, 18^ and MsCAIVAR^8^, SuSiEx infers distinct credible sets for each causal signal (Figure 1), facilitating the interpretation of fine-mapping results, and is orders of magnitudes more scalable computationally (see below), enabling the analysis of large, complex loci and biobank-scale datasets across many complex traits and diseases.

### SuSiEx outperformed single-population and non-Bayesian cross-population fine-mapping methods in simulations

We conducted a series of simulations to systematically evaluate the performance of SuSiEx. We randomly selected 100 1Mb regions from chromosome 1 with an average of 6,548 variants per region (Supplementary Table 1), and simulated individual-level genotypes of European (EUR), East Asian (EAS) and African (AFR) populations using HAPGEN2^19^ and 1000 Genomes Project^20^ samples as the reference panel. For each genomic region, we generated phenotypic data under different numbers of causal variants (*n_csl_*; assuming that they are shared across populations), cross-population genetic correlations (*r_g_*) and local SNP heritability (*h*^2^). We then performed GWAS in each region and calculated in-sample LD matrices, which were used as inputs to different summary statistics based fine-mapping methods. To examine the impact of these genetic parameters on fine-mapping results, we defined a standard simulation setting with *n_csl_* = 1, *r_g_* = 0.7 and *h*^2^ = 0.1%, and varied these parameters to produce a range of local genetic architectures (Supplementary Table 2). Given a set of genetic parameters, we further assessed the impact of different population and discovery sample size combinations (Supplementary Table 3) on fine-mapping results. For each simulation setting (different combinations of genetic parameters and discovery sample sizes), the simulation was replicated 5 times for each genomic region, producing a total of 500 simulated datasets for analysis.

Throughout the simulation study, in single-population fine-mapping, we analyzed loci that reached genome-wide significance in population-specific GWAS (P<5E-8); in cross-population fine-mapping, we analyzed loci that reached genome-wide significance in at least one of the population-specific GWAS or in the cross-population fixed-effect meta-analysis. We assessed the performance of different fine-mapping methods using an array of metrics: (i) Coverage/Calibration: the proportion of 95% credible sets that include at least one true causal variant across simulation replicates; (ii) Power: the number of true causal variants identified (i.e., covered by a 95% credible set); (iii) Resolution: the size of 95% credible sets and the number of fine-mapped variants with high confidence (e.g., PIP >95%); (iv) Scalability: the computational cost/feasibility to perform fine-mapping in large genomic loci; (v) Robustness: the proportion of runs in which the fine-mapping algorithm converges and returns sensible results (defined later). We focused on constructing and evaluating 95% credible sets (i.e., each credible set has at least 95% probability of containing a true causal variant) in simulations.

As expected, in the standard simulation setting (Figure 2; Supplementary Figures 1 & 2; Supplementary Table 4), compared with single-population fine-mapping, integrating data across populations using SuSiEx led to better power (i.e., more true causal variants being identified; Figure 2a) and had higher resolution (i.e., smaller credible sets and more causal variants with high PIP; Figure 2b & 2d). Meanwhile, SuSiEx had well-calibrated coverage at 95%, regardless of the populations from which data were combined (Figure 2c). The magnitude of improvements in power and resolution depends on both the increase in the total sample size and the LD diversity in the discovery samples (Figure 2; Supplementary Table 4). For example, adding 50K EUR individuals to an existing EUR sample of 50K individuals increased the number of identified causal variants with PIP >95% from 18 to 26 (out of 500 causal variants simulated), and reduced the median size of the credible set from 11 to 8. The yield of causal variants with PIP >95% was much greater (increased from 18 to 78 out of 500 causal variants simulated) and the median size of the credible set was much smaller (reduced from 11 to 5) if the added 50K individuals were of AFR instead of EUR ancestry, demonstrating the importance of genetic diversity in cross-population fine-mapping. The inclusion of 50K individuals of EAS ancestry also provided a greater yield of causal variants with PIP >95% (increased from 18 to 44 out of 500 causal variants simulated) and smaller credible sets (reduced from 11 to 7) relative to adding 50K EUR samples, although the benefits were less pronounced than when the AFR samples were added, due to the smaller LD blocks in the African ancestries^21, 22^.

**Figure 2:**
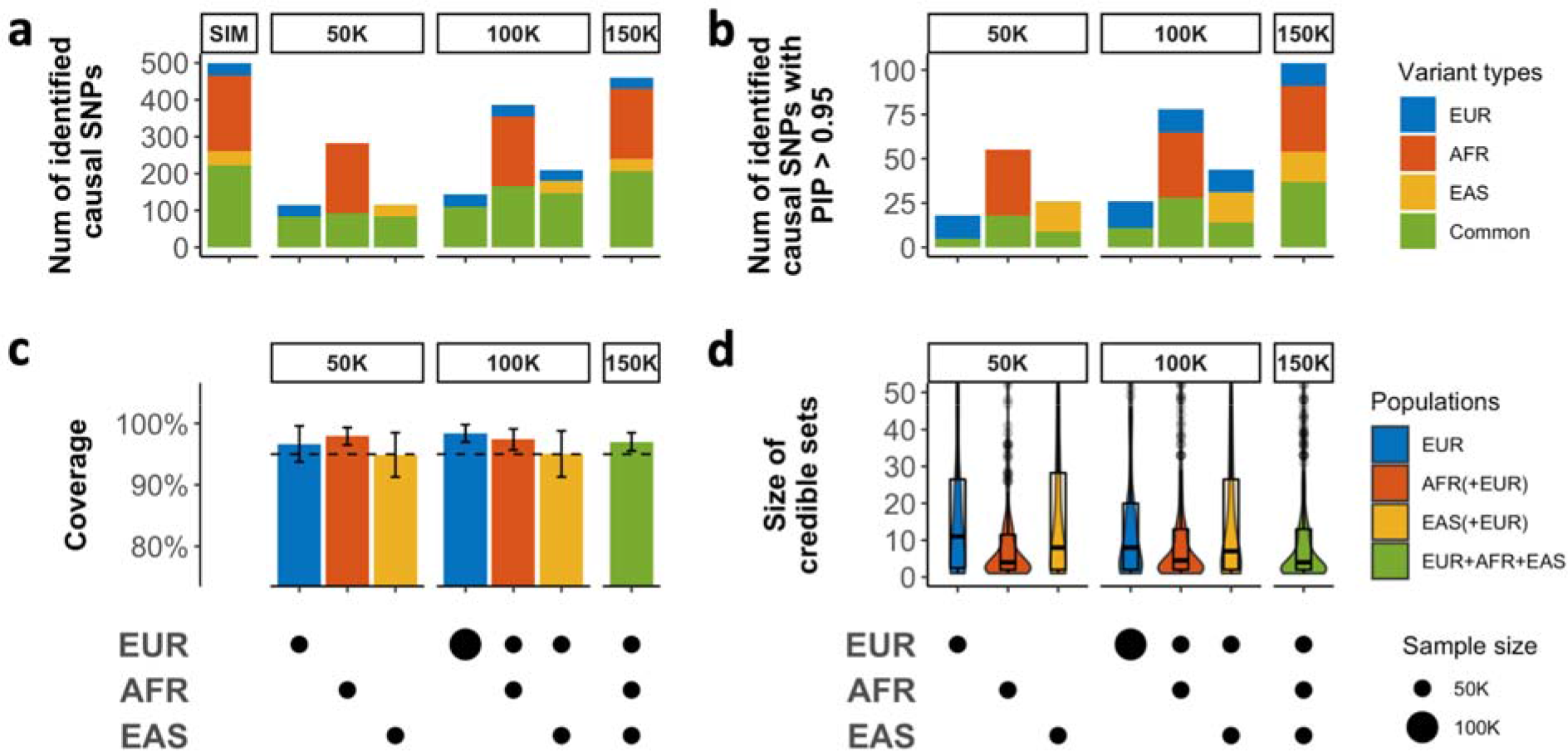
The performance of SuSiEx in simulations. Simulated data were generated under the standard parameter setting (Methods). **a**, The number of identified true causal variants (true causal variants covered by a 95% credible set) when integrating data from different populations with different sample sizes for fine-mapping. SIM indicates the number of different types of causal variants simulated. **b**, The number of true causal variants mapped to PIP >95%. **c**, The coverage of 95% credible sets (the proportion of credible sets that contain a true causal variant). The dashed line represents 95% coverage. The error bars represent 95% confidence intervals. **d**, Distribution of the size of credible sets. The upper and lower bounds of the box indicate the 75th and 25th percentiles, respectively. The middle line in the box indicates the median. In **a-d**, the top label of each subpanel indicates the total sample size, and the bottom panels indicate the sample size from each population. In **a** and **b**, variants with MAF >0.5% only in one population were defined as specific to that population, and all other variants were defined as “common” (i.e., shared variants across populations).

A widely used approach in recent multi-ancestry genetic studies^4^ is to apply a single-population fine-mapping method to meta-analyzed GWAS summary statistics and LD matrices. Despite of its convenience, this method can be miscalibrated and does not unleash the full potential of genomic diversity, likely due to its over-simplified modeling of LD across populations, the presence of population-specific variants, and the strong assumption in fixed-effect meta-analysis that cross-population effect sizes are homogeneous^6^. We confirmed, using the standard simulation setting, that fine-mapping using meta-analyzed GWAS and sample size weighted LD suffered substantial loss in both power and coverage (Supplementary Figure 3; Supplementary Table 5). In contrast, SuSiEx, through explicit and flexible modeling of population-specific association statistics and LD, identified many more causal variants (Supplementary Figure 3a) and was always well-calibrated (Supplementary Figure 3b). Additional simulations showed that when causal effect sizes or LD patterns differ, meta-analysis-based methods can lead to inflated false positive rates and power loss even when combining subpopulation groups within a continent (e.g., CEU and FIN; YRI and LWK; CHB and JPT; Supplementary Figures 4 & 5; Supplementary Table 6). Overall, meta-analysis-based methods are only valid and powerful when combining independent datasets from the same population (Supplementary Figure 6; Supplementary Table 7).

A recent study used a clustering method to combine single-population fine-mapping results, which was applied to multiple large-scale biobanks with promising biological discoveries^7^. However, this approach does not make use of subthreshold association signals, and does not leverage LD diversity to improve the resolution of fine-mapping. In simulations, SuSiEx identified more true causal variants especially when the GWAS sample size was moderate or small, as expected for current non-EUR GWAS (Supplementary Table 5). For example, when analyzing 50K EUR and 50K AFR individuals under the standard simulation setting, the single-population combining method identified a smaller number of causal variants compared with SuSiEx (375 vs. 386). Although the number of true causal variants discovered by both methods became comparable when the GWAS sample sizes became larger, SuSiEx outperformed the combining method in fine-mapping resolution, as measured by the size of credible sets and the number of true causal variants with high PIP (>50% or >95%; Supplementary Figure 7; Supplementary Table 5). For example, under the standard simulation setting, SuSiEx produced smaller credible sets on average (median size 2 vs. 4) and identified a larger number of causal variants with PIP >95% (161 vs. 140) compared with the single-population combining method when analyzing 200K EUR and 200K AFR individuals (Supplementary Table 5).

### SuSiEx outperformed existing Bayesian cross-population fine-mapping methods in simulations

We next compared SuSiEx with two published Bayesian cross-population fine-mapping methods, PAINTOR^9, 18^ and MsCAVIAR^8^, using the standard simulation setting. For the vast majority of the loci and simulation replicates, SuSiEx converged within a minute and a small number of iterations using a single CPU (Supplementary Figures 8 & 9; Supplementary Table 8). In contrast, neither PAINTOR nor MsCAVIAR was capable of analyzing all common variants (MAF >1% in EUR, EAS or AFR) in a 1Mb locus (6,548 variants per locus on average; Figure 3a, left column). In particular, MsCAVIAR is not computationally scalable and cannot complete analyzing a genetic locus within 24 hours, while PAINTOR always returned unreasonable results, in which the sum of PIP across variants in a genomic locus >5 or <0.1. We note that in the standard simulation setting, the number of true causal variants was set to one in each locus, and thus a sum of PIP >5 or <0.1 appears “unreasonable” and may indicate severe model fitting issues such as failure to converge. We then filtered the discovery summary statistics to fewer variants to enable performance evaluation across methods. Specifically, we created three input datasets with increasingly stringent selection criteria: “p < 0.05”, “top 500” and “top 150”, corresponding to marginal P <0.05, the top 500 and the top 150 most associated variants, respectively. With these filtered input datasets, the “enumerate” mode of PAINTOR, with the number of causal variants set to one (which matched the simulation parameter, and was thus a favorable setting for PAINTOR) still returned unreasonable results (sum of PIP >5 or <0.1) for approximately 25% of the analyses, while the “MCMC” mode of PAINTOR returned unreasonable results for almost all the analyses, with zero PIP for every variant (Figure 3a; Supplementary Table 9). The “enumerate” mode of PAINTOR was also highly sensitive to the parameter “maximum number causal SNPs”, which is typically unknown *a priori* and difficult to set in practice (Extended Data Figure 2). The other Bayesian fine-mapping method, MsCAIVAR, was only able to analyze the smallest input dataset (“top 150”), as larger dataset took more than 24 hours per locus (Figure 3a), although the results were generally “reasonable” (Extended Data Figure 2; Supplementary Table 9).

**Figure 3:**
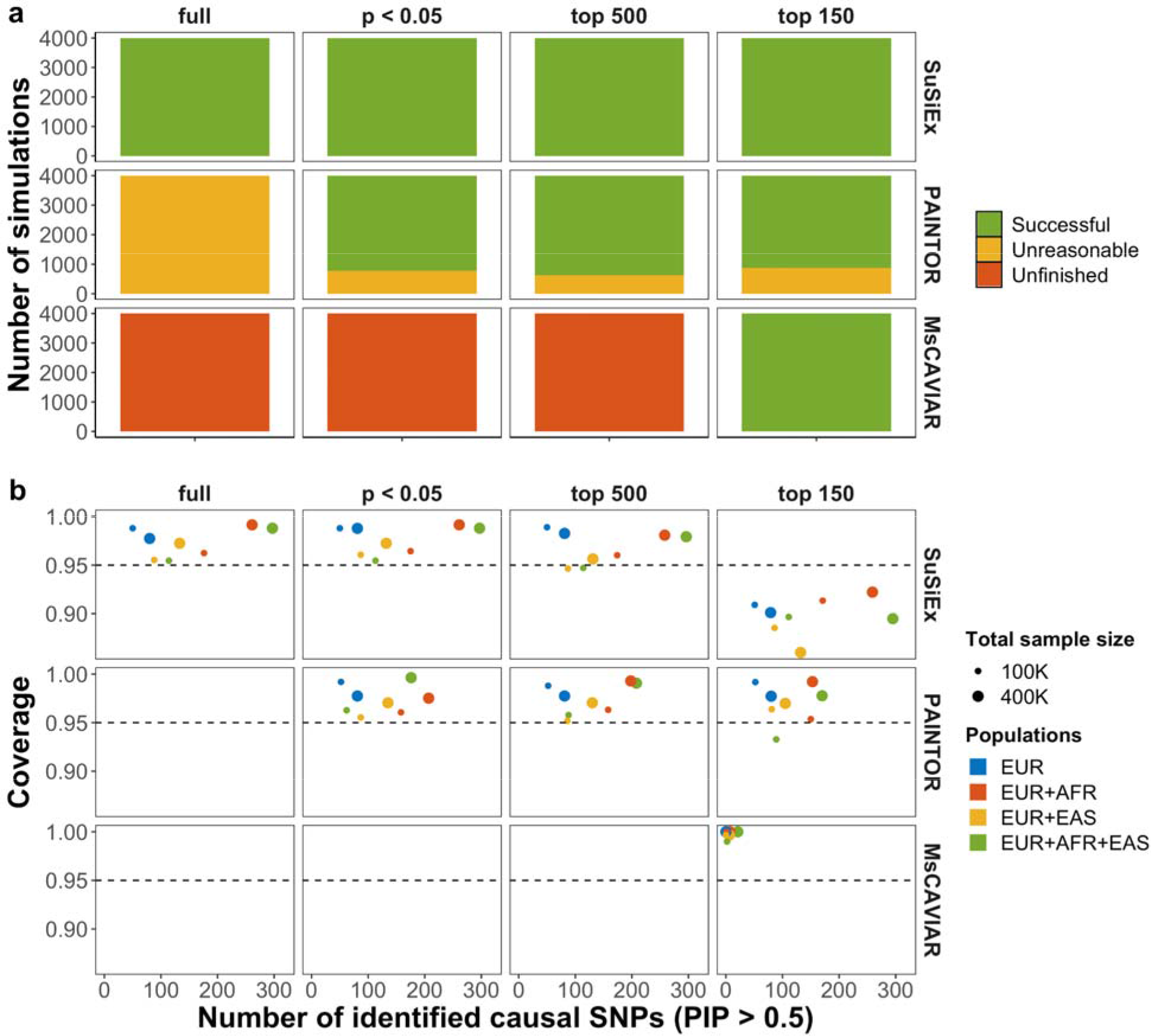
Comparison of SuSiEx, PAINTOR and MsCAVIAR in simulations. **a**, The job completion summary (scalability and robustness) for Bayesian fine-mapping methods using different numbers of input variants. PAINTOR was run using the “enumerate” mode with “-enumerate=1” (which matched the simulation parameter). Unfinished: jobs taking longer than 24 hours wall time. Unreasonable: jobs returning unreasonable results, defined as the sum of PIP across variants in the genomic locus >5 or <0.1 (1 is expected). Successful: jobs completed within 24 hours of wall time and returned reasonable results. **b**, Number of identified true causal variants with PIP >50% (x-axis) versus the coverage of credible sets (y-axis) for different input datasets and fine-mapping methods. Only simulation runs that were completed within 24 hours and returned reasonable results were included.

For each method, we then focused on simulation runs that returned reasonable PIP estimates. PAINTOR, with the “enumerate” mode and the number of causal variants set to one, had calibrated results at 95% coverage and identified a similar number of high-PIP causal variants to SuSiEx in the EUR-only and EUR + EAS fine-mapping (PIP >50%; Figure 3b). MsCAVIAR, however, identified much fewer causal variants with PIP >50% (Figure 3b). This is because MsCAVIAR tends to return large credible sets containing almost all the variants in the input dataset, each having a small PIP (Supplementary Table 10). SuSiEx outperformed PAINTOR and MsCAVIAR in the number of causal variants identified with PIP >50%, when AFR samples were included in the discovery GWAS (Figure 3b), suggesting that SuSiEx can leverage genomic diversity to fine-map more causal variants with high accuracy. For example, when combining 200K EUR and 200K AFR samples, SuSiEx identified 261 unique causal variants with PIP >50% using the full GWAS summary statistics, comparing with 209 identified by PAINTOR and 7 identified by MsCAIVAR across the four input datasets (Figure 3b; Supplementary Table 10). We note that the coverage for SuSiEx was well-calibrated in most settings but dropped below 95% when the top 150 most associated variants were used as input, likely due to information loss from variant filtering. Since using the full GWAS summary statistics as input was computationally tractable and yielded optimal results for SuSiEx, we do not consider this a limitation for SuSiEx and do not recommend any prefiltering of variants when using SuSiEx in practice.

### SuSiEx is robust to varying cross-population genetic architectures and model misspecifications

We examined the calibration, power and resolution of SuSiEx by varying key parameters in the standard simulation setting. The cross-population genetic correlation (*r_g_*) can be less than one for many complex traits and diseases^23^. SuSiEx accounts for imperfect genetic correlation by allowing for varying genetic effects across populations. Using simulated data with *r_g_* of 0.4, 0.7, and 1.0, we confirmed that SuSiEx was robust to a range of *r_g_*values, with good calibration and similar power and resolution (Supplementary Figures 10-14; Supplementary Table 11). The local SNP heritability (*h*^2^) and the number of causal variants (*n_csl_*) per locus can differ across the genome for a given trait or disease^1, 24–26^. We set the heritability per locus to 0.05%, 0.1%, 0.2%, 0.3%, 0.4% and 0.5%, and for a given per-locus heritability, varied *n_csl_*from 1 to 5 with each genetic effect drawn from a normal distribution (Methods). As expected, SuSiEx performed better when *h*^2^ increased (Supplementary Figures 15-19; Supplementary Table 12) and *n_csl_* decreased (Supplementary Figures 20-24; Supplementary Table 13), which corresponds to higher per-variant heritability and thus larger statistical power. Nonetheless, SuSiEx was always well-calibrated (Supplementary Figures 16 & 21), and was able to capture multiple causal variants in the same locus as *n_csl_* increased. SuSiEx was also robust to different values of the hyperparameters in the model (Supplementary Figures 25 & 26; Supplementary Table 14).

We additionally assessed the robustness of SuSiEx under model misspecifications. SuSiEx assumes that causal variants are shared across populations. As the frequency or allelic effect size of a causal variant becomes smaller in a population, the benefits of including data from that population in fine-mapping become smaller. At the extreme of this continuous frequency and effect size spectrum, a causal variant in some population(s) may be missing or null in other populations, violating the assumption of the SuSiEx model and adding noise to cross-population fine-mapping. However, Figure 2a and 2b show that SuSiEx can capture population-specific causal variants that are rare (MAF <0.5%) in all but one population. We further evaluated the robustness of SuSiEx by simulating causal variants that had non-zero effect sizes in one population but were null in other populations, including a scenario where a variant was only causal in the smaller AFR sample (N=50K) and was null in both EUR and EAS with much larger sample sizes (N=500K and 100K, respectively). We found that, by allowing for different effect sizes (including null effects) of a causal variant across populations, the performance of SuSiEx, including calibration and power, was minimally impacted by adding null data to the analysis (Supplementary Figures 27 & 28; Supplementary Table 15), confirming the robustness of SuSiEx to model misspecifications.

### Inference of population-specific causal status

Once converged, SuSiEx estimates the population-specific causal probability for each inferred credible set, and uses a probability threshold of 0.8 to infer population-specific causal status. We performed simulations across a wide range of settings to assess whether this inference is accurate. For example, in the scenario of two populations (EUR and AFR) with a balanced sample size, we simulated three causal configurations: (i) a EUR-specific causal variant (non-zero effect in EUR, zero effect in AFR); (ii) an AFR-specific causal variant (zero effect in EUR, non-zero effect in AFR); (iii) a shared causal variant (non-zero effect in both populations), using *r_g_* = 0.7 and *h*^2^ = 0.1%. Supplementary Figure 29 shows that when the causal variant was EUR-specific or AFR-specific, SuSiEx accurately inferred the causal configuration with >98% accuracy. When the causal variant was shared between EUR and AFR populations, the overall accuracy of the inference became lower especially when the GWAS sample size was small, but the vast majority of the misclassified causal variants had low allele frequencies and/or small effect sizes, and were thus underpowered to be identified (Supplementary Figure 30). As the frequency, effect size or GWAS sample size became larger, providing increasing evidence to support the causal effect, the accuracy of the inference substantially improved (Supplementary Figures 29 & 30; Supplementary Table 16). Additional simulations across different numbers of causal variants, cross-population genetic correlations, SNP heritability and GWAS sample sizes, as well as different causal configurations in three populations confirmed that SuSiEx can accurately infer whether a well-powered signal is causal in a population (Supplementary Figures 31-38; Supplementary Tables 16 & 17).

### The impact of LD mismatch on the performance of SuSiEx

Lastly, we note that in-sample LD is preferred in fine-mapping as it matches the correlation pattern between variants in the discovery GWAS sample. Unfortunately, in-sample LD is not always available, especially in large-scale GWAS comprising multiple cohorts. Using an external LD reference panel from a genetically close population can be a pragmatic solution despite its limitations^6, 27–29^. Here, we evaluated the impact of LD mismatch on SuSiEx. Consistent with previous findings, analysis using in-sample LD produced excellent calibration and power, while using external LD led to coverage and power loss as the genetic distance between the external reference panel and the discovery sample increased (Supplementary Figure 39; Supplementary Table 18).

### SuSiEx increased the power and resolution of fine-mapping in biobank analysis

We applied SuSiEx to data from the Pan-UKBB project and the Taiwan Biobank (TWB). The Pan-UKBB project is a multi-ancestry resource derived from the UK Biobank (UKBB)^14^ by analyzing six continental ancestry groups across 7,228 phenotypes. We included summary statistics of EUR and AFR ancestries from Pan-UKBB (*N*_EUR_ up to 419,807; *N*_AFR_ up to 6,570, Supplementary Table 19). We additionally included TWB, one of the largest biomedical databases in East Asia (*N*_EAS_ = 92,615)^15, 30^. We selected 25 quantitative traits shared between Pan-UKBB and TWB (Supplementary Table 19), and defined 13,420 genomic loci that reached genome-wide significance in at least one of the single-population association analysis or a fixed-effect meta-analysis across the three populations (Methods; Supplementary Table 20). A random-effect meta-analysis did not identify additional loci (Method). We then performed single-population fine-mapping using SuSiE, and cross-population fine-mapping using SuSiEx, combining EUR, AFR and EAS data. In both SuSiE and SuSiEx analyses, we used in-sample LD as the input and examined 99% credible sets.

SuSiEx identified a total of 14,361 credible sets across 9,827 loci, among which 14,115, 725 and 5,699 credible sets were inferred as causal (with population-specific causal probability >0.8) in the EUR, AFR and EAS populations, respectively (Supplementary Table 20). In contrast, single-population fine-mapping identified 13,205, 51 and 1,672 credible sets for the EUR, AFR and EAS populations, respectively (Supplementary Table 20). We note that the population-specific causal probability estimated by SuSiEx is affected by statistical power. A small probability inferred from the current dataset can become greater when the sample size increases, and thus should not be used to rule out the potential causal role of the variant, akin to a non-significant *P*-value from GWAS when the sample size is limited. We expect that, with increasing sample sizes, many of the credible sets will be inferred as causal in non-European populations.

Aligning credible sets across analyses (Methods) led to 2,278 (15.9%) credible sets identified by SuSiEx that were not identified by single-population fine-mapping (Supplementary Table 20), suggesting that SuSiEx can capture more putative causal signals than single-population fine-mapping. In addition to identifying and mapping more genetic associations through integrating data from multiple populations, SuSiEx also improved fine-mapping resolution. Relative to single-population fine-mapping in the EUR population, adding AFR and EAS data increased the average of the maximum PIP across all aligned credible sets from 0.42 to 0.46 (*P* = 7.3E-12; two-sided *t* test), and reduced the average size of credible sets from 30.5 to 28.0 (*P* = 6.7E-3; two-sided *t* test; Figure 4a & 4b; Supplementary Table 21; Extended Data Figure 3a as an example). Additionally, cross-population fine-mapping identified 2,466 putative causal variants with PIP >95% (Figure 4c; Supplementary Table 22), among which 437 were not discovered by any single-population fine-mapping. For example, SuSiEx identified a credible set containing a single missense variant of *TRIM5* (rs11601507; 11:5701074:C:A) associated with total bilirubin at PIP >99%. Due to the limited statistical power, this credible set failed to reach genome-wide significance in any population and was missed in single-population fine-mapping (GWAS *P*-value: *P_EUR_* = 1.9E-6, *P_AFR_* = 2.4E-2, *P_EAS_* = 1.1E-4; Figure 5a; Extended Data Figure 4). However, SuSiEx estimated that the causal probabilities of this missense variant in EUR, AFR, and EAS were 0.998, 0.743 and 0.994, respectively, suggesting that this putative causal variant may be shared between populations (Supplementary Table 21). Similarly, SuSiEx identified a two-variant credible set associated with albumin that failed to reach genome-wide significance in any population (Figure 5b; Extended Data Figure 5). The lead variant (13:31312178:A:T) in this credible set is an intron variant of *ALOX5AP* with PIP 97.4%, which was shared between EUR and EAS populations (population-specific causal probability: *P_EUR_* = 1.000, *P_AFR_*= 0.405, *P_EAS_* = 0.998; Supplementary Table 21). This variant was fine-mapped to be an eQTL variant regulating the expression of *ALOX5AP* in whole blood (PIP >99%), artery aorta (PIP = 86.1%) and spleen (PIP = 77.9%) (Figure 5b; Extended Data Figure 5)^31^. In both examples, SuSiEx identified putative causal variants and resolved a genetic locus to its gene target that would have been missed if only single-population fine-mapping was performed.

**Figure 4:**
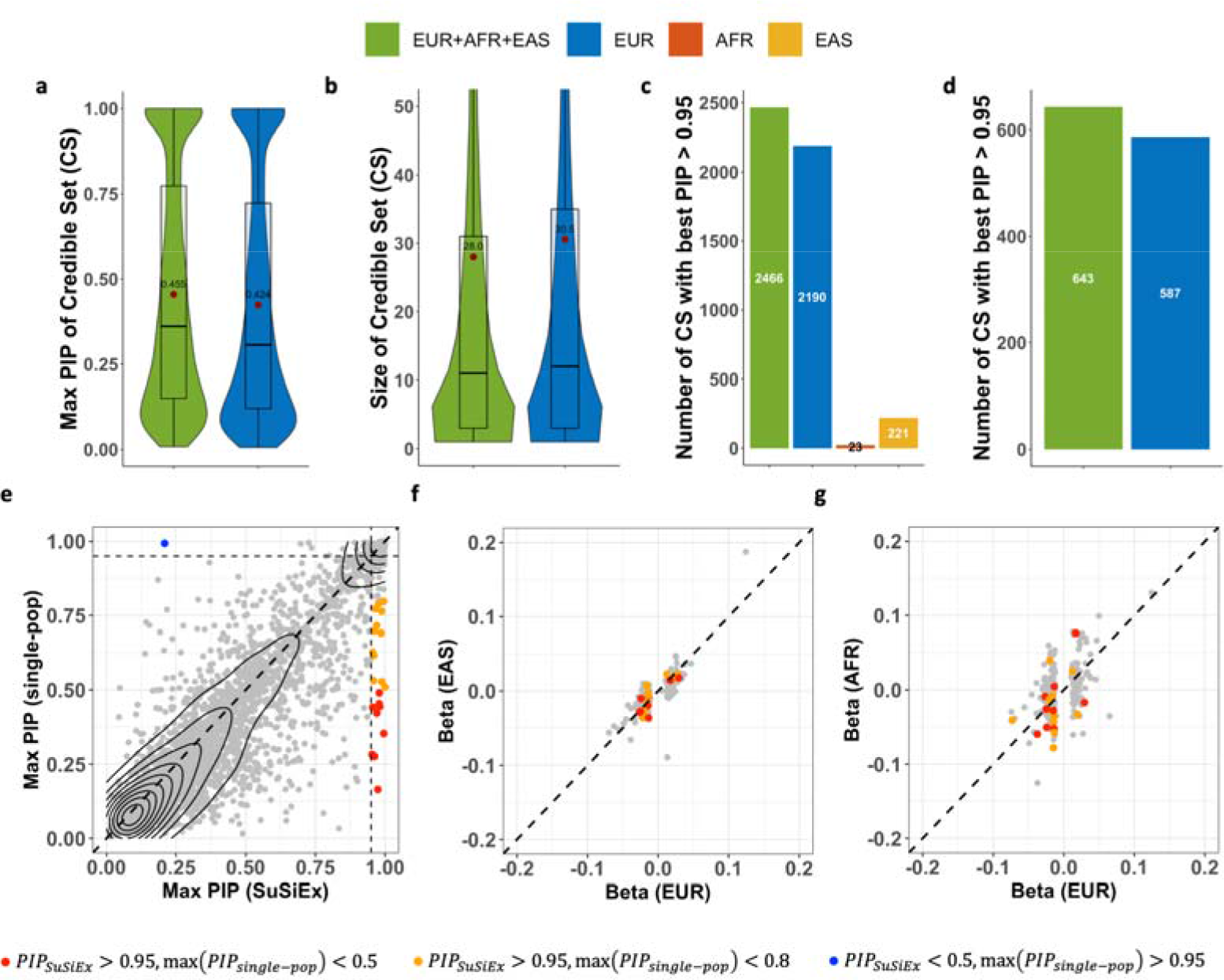
Cross-population fine-mapping analysis in biobanks. **a**, The distribution of the maximum PIP across 99% credible sets. **b**, The distribution of the size of 99% credible sets. **c**, The number of variants mapped to PIP >95% across 99% credible sets. **d**, The number of variants mapped to PIP >95% in single-credible-set loci. **e**, The maximum PIP from SuSiEx versus the maximum value of the maximum PIP in the three single-population fine-mapping using SuSiE. Only genomic loci with a single credible set aligned across analyses were included. **f** and **g**, The marginal per-allele effect size of the maximum PIP variant in EUR vs. EAS and EUR vs. AFR populations. Variants in single-credible-set loci with PIP >95% estimated by SuSiEx and minor allele frequencies >5% in all populations were included. In **a-b**, red dots represent the mean, the middle line in the box represents the median, and the upper and lower bounds of the box represent the 75th and 25th percentiles, respectively.

**Figure 5.**
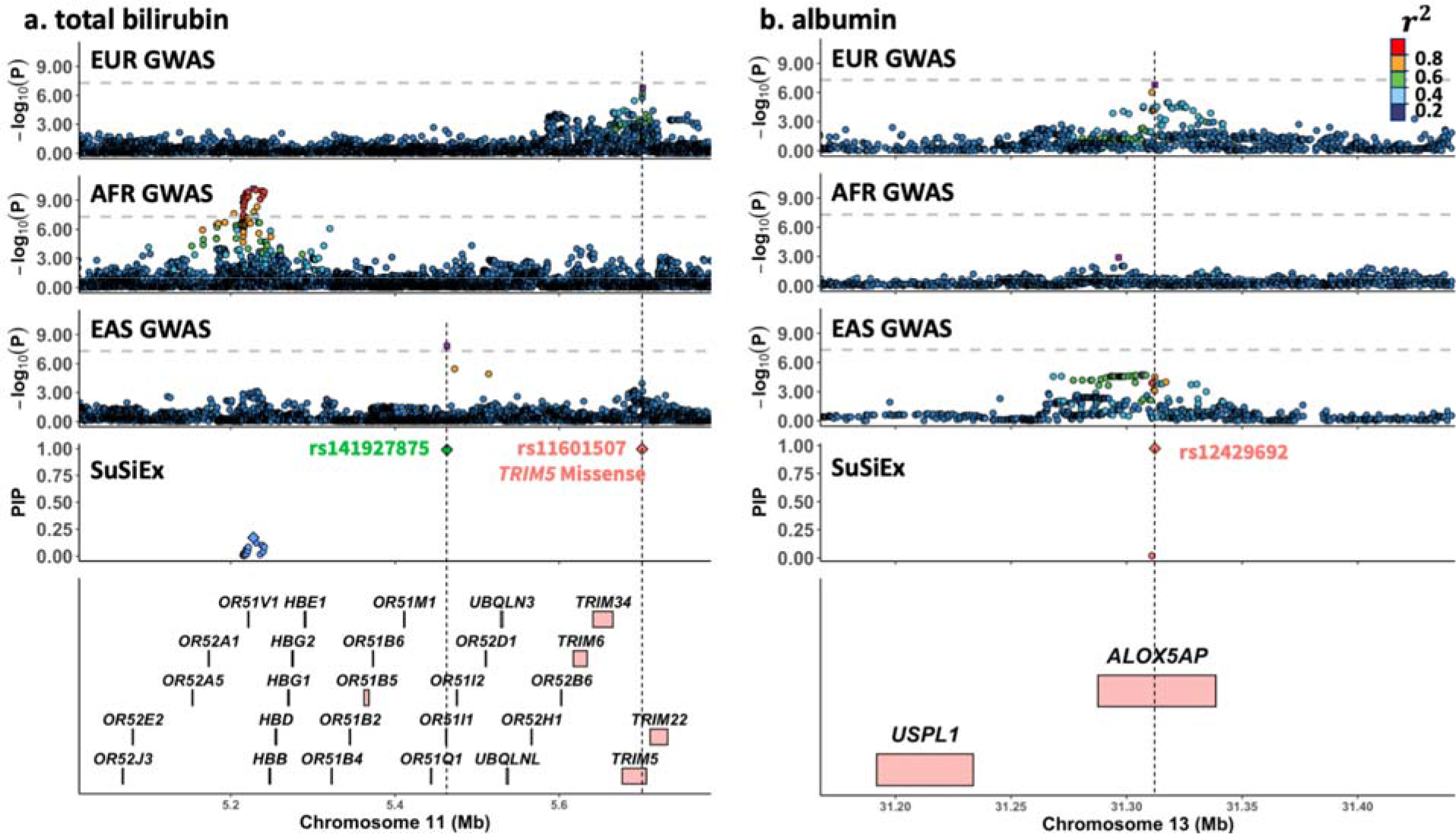
SuSiEx identified variants missed in single-population fine-mapping. Each sub-figure consists of five panels, which are aligned vertically, with the x-axis showing the genomic position. The top three panels visualize GWAS association statistics of the European (Pan-UKBB Europan), African (Pan-UKBB African) and East Asian (Taiwan biobank) populations following the LocusZoom^35^ style. The second to bottom panel visualizes the fine-mapping results from SuSiEx, which integrated GWAS summary statistics from the three populations.

Next, we restricted the comparison to loci that were mapped to a single credible set by both single- and cross-population fine-mapping such that our results were not affected by multiple causal variants in LD and the algorithm that aligns credible sets. In these single-credible-set loci, SuSiEx continued to outperform single-population fine-mapping in power and resolution, identifying more credible sets with high confidence (maximum PIP >95%; Figure 4d), and improving the maximum PIP of a credible set in general relative to single-population fine-mapping (*P* = 6.7E-5; two-sided *t* test; Figure 4e). In particular, SuSiEx improved the maximum PIP of 30 credible sets from <80% to >95% (Figure 4e; orange and red dots), among which 9 were improved from <50% to >95% (Figure 4e; red dots). We note that the maximum PIP for one credible set dropped substantially, from 99% to 21%, in the cross-population fine-mapping (Figure 4e; blue dot). Further investigation of this locus revealed that the putative causal variant (12:67643414:T:A) is located in a low complexity genomic region, where the quality of variant calling and imputation may be negatively affected^32^. This variant is also represented in fewer than 50% of individuals in gnomAD v2.1.1 genomes^33^, and violates Hardy-Weinberg equilibrium.

Biobank analyses further confirmed that SuSiEx can capture population-specific causal variants (Extended Data Figure 3c as an example). Despite a dominating EUR sample size, SuSiEx recaptured 87.5% of the findings from single-population fine-mapping. A non-trivial proportion of credible sets from single-population fine-mapping that were estimated to have lower maximum PIPs by SuSiEx may be attributed to quality-related concerns, including (i) the best PIP variant is in the low complexity region (LCR); (ii) the best PIP variant is in allelic imbalance or violates Hardy Weinberg equilibrium in gnomAD^33^; or (iii) the best PIP variant is multi-allelic or colocalizes with indels at the same genomic position, which might influence imputation quality. For example, 17.5% (29/166) of the putative causal variants with PIPs dropped by 10-20% in cross-population fine-mapping relative to single-population fine-mapping had quality issues, compared with 41.2% (7/17) of the variants with PIPs dropped by >40% (Extended Data Figure 6). These results suggest that, through the joint modeling of multiple populations and datasets, SuSiEx provides the additional benefit of identifying and removing likely low-quality findings from single-population analyses. Removing variants with potential quality issues before fine-mapping improved both single-population and cross-population fine-mapping, and SuSiEx continued to outperform single-population fine-mapping methods (Supplementary Figure 40).

We used Ensembl Variant Effect Predictor (VEP)^34^ to annotate each variant into high, moderate or low functional impact, as well as modifiers. As the inferred PIPs increased, the proportion of variants with high impact clearly increased (Extended Data Figure 7), suggesting that confidently fine-mapped variants were enriched among mutations of functional importance. In total, we identified 2,112 high or moderate impact variants in 99% credible sets located in 1,529 genes. Among these variants, 403 had a PIP greater than 50% (Supplementary Table 23), and 261 had a PIP greater than 95% (Supplementary Table 24). There were 25 genes containing at least two high/moderate impact SNPs with PIP greater than 95%, while only 23 were detected in the three single-population fine-mapping analyses. In particular, *IQGAP2* and *PIEZO1* carried 3 missense variants associated with multiple blood biomarkers with PIPs >95%.

Lastly, we compared the per-allele effect sizes of high-confidence putative causal variants (PIP >95% in single- or cross-population fine-mapping) located in single-credible-set loci among EUR, AFR and EAS populations (Figure 4f & 4g). As no secondary association was found in these loci, we used marginal effect sizes in the comparison. Overall, the effect sizes were highly concordant between EUR and EAS populations (*r* = 0.79) but less consistent between EUR and AFR populations (*r* = 0.21). Downsampling the EAS dataset to TWB batch 1 samples (N=27,033) and rerunning the SuSiEx analysis produced a substantially lower effect size correlation (*r* = 0.29) between EUR and EAS populations, suggesting that the limited non-European GWAS sample size could be a major factor that influenced the correlation estimate (Supplementary Figure 41). Expanding non-European genomic resources will be critical to investigate the consistency of causal effect sizes across populations.

The bottom panel shows gene annotations. For the GWAS panels, the left y-axis shows the - log_10_(p-value) of each SNP. The gray horizontal dash line represents the genome-wide significance threshold (5E-8). The purple rectangle for each locus represents the lead (most associated) variant. Variants are colored by descending LD with the lead variant (ordered red, orange, green, light blue, and dark blue dots). For the fine-mapping panels, different colors were used to distinguish different credible sets. The diamond represents the maximum PIP variant of each credible set. **a,** Association with total bilirubin on chr11: 5,100,000-5,700,000. **b,** Association with albumin on chr13: 31,150,000-31,450,000.

### SuSiEx identified additional putative causal candidates for schizophrenia

We applied SuSiEx to schizophrenia GWAS summary statistics of EUR (*N_case_* = 53,251, *N_control_* = 77,127) and EAS (*N_case_*= 14,004, *N_control_* = 16,757) ancestries from the Psychiatric Genomics Consortium (PGC), and fine-mapped the same 250 autosomal loci in the recent PGC publication^4^. SuSiEx successfully identified 215 99% credible sets out of 193 loci (not all loci converged to a credible set, as in all fine-mapping analyses), among which 213 were inferred as causal in EUR (population-specific causal probability >0.8) and 95 were causal in the EAS population. Out of the 213 credible sets, 11 contained a SNP with PIP >95% (Figure 6a; Supplementary Tables 25 & 26). As expected, SuSiEx outperformed published PGC fine-mapping results, which applied a single-population fine-mapping method, FINEMAP^36^, to meta-analyzed GWAS summary statistics and sample size weighted LD^4^. Specifically, SuSiEx mapped 57% (33 vs. 21) more signals to a single variant with PIP >50% in single-credible-set loci (Figure 6). Most of the SuSiEx-improved credible sets had a marginally genome-wide significant signal (*P*-value between 5E-8 and 1E-15; Figure 6b & 6c). SuSiEx also produced credible sets for three loci that could not be resolved by FINEMAP in the original analysis. In these loci, FINEMAP inferred five independent credible sets, each containing a single variant that was not statistically significant in the GWAS, likely due to inaccurate reference panel^37^. Furthermore, SuSiEx substantially increased the resolution of fine-mapping by reducing the average size of credible sets from 87.1 to 60.3 (*P* = 0.015; paired two-sided *t* test), and increasing the average of maximum PIP across credible sets from 0.25 to 0.27 (*P* = 0.012; paired two-sided *t* test). As an example, SuSiEx successfully fine-mapped the *FURIN* gene region, pinpointing the 3’UTR variant rs4702, which has been reported as a causal variant in earlier studies^38, 39^. Although this variant did not reach genome-wide significance in the EAS population (*P* = 1.06E-3) likely due to limited statistical power, SuSiEx strongly supported that this causal variant is shared between EUR and EAS populations, with population-specific causal probabilities of 1.000 and 0.976, respectively.

**Figure 6:**
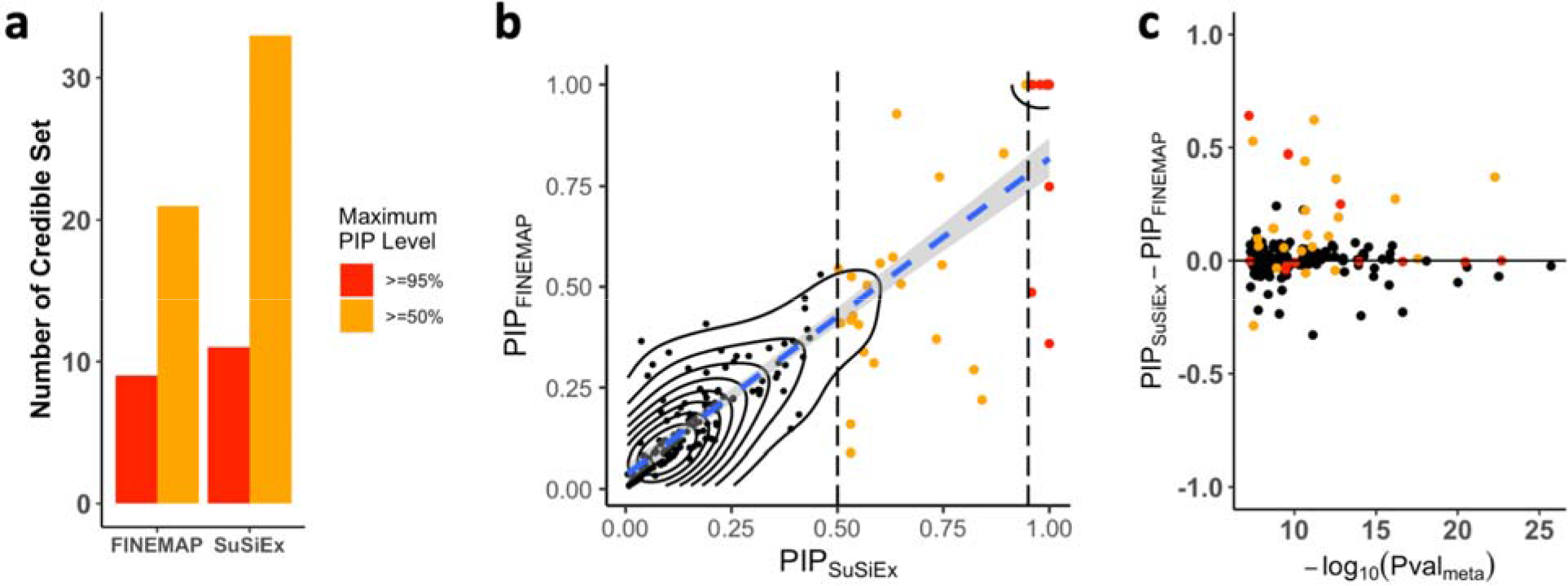
Fine-mapping of schizophrenia risk loci across European and East Asian populations. **a**, The number of putative causal variants mapped to PIP >50% and >95% by FINEMAP and SuSiEx in single-credible-set loci. **b**, The maximum PIP for each credible set within single-credible-set loci, estimated by SuSiEx and FINEMAP. **c**, The difference of the maximum PIP, estimated by SuSiEx and FINEMAP (y-axis), within each single-credible-set locus, plotted against the -log_10_(p-value) of the most associated variant in the cross-population meta-analysis. In **b** and **c**, red dots represent credible sets with a maximum PIP >95% estimated by SuSiEx; orange dots represent credible sets with a maximum PIP >50% estimated by SuSiEx.

## DISCUSSION

We presented SuSiEx, a cross-population fine-mapping method which links multiple population-specific sum of single effects (SuSiE) models by assuming the sharing of underlying causal variants. Through flexible and accurate modeling of varying population-specific causal effect sizes and LD patterns, SuSiEx improves the power and resolution of fine-mapping while producing well-calibrated false positive rates and retaining the ability to identify population-specific causal variants. We showed, via comprehensive simulation studies, that SuSiEx is highly computationally efficient, outperforms alternative cross-population fine-mapping methods in calibration, power and resolution, and is robust to model misspecifications. In particular, as the two state-of-the-art Bayesian cross-population fine-mapping methods, PAINTOR is sensitive to the predefined (yet unknown) number of causal variants, while MsCAVIAR is computationally intractable when the total number of input variants is greater than a few hundred. Moreover, neither method has the capacity to analyze summary statistics from a comprehensive set of common variants in loci greater than 1Mb. SuSiEx overcomes these limitations and offers effective and efficient cross-population fine-mapping that can be applied on biobank-scale datasets for the first time.

A key assumption made by the SiSiEx model is that causal variants are shared across populations, which enables the integration of data from multiple populations to improve the power and resolution of fine-mapping. As SuSiEx allows the causal effect size to vary across populations without restrictions and does not penalize small or zero effects, the model remains valid and well-calibrated for arbitrarily small or even null effects. An alternative modeling strategy may be explicitly modeling the causal configuration of each credible set across populations (i.e., whether a signal is causal in a population or not), similar to the methods developed for multi-trait colocalization^40, 41^. However, since the number of possible causal configurations grows exponentially with both the number of populations and the number of causal variants, existing techniques become computationally impractical when analyzing a genomic region with a handful of causal variants in more than two populations. Given that the causal effect size varies along a continuous spectrum, in contrast to making a binarized inference of whether a causal signal is significant or not in a population, SuSiEx focuses on estimating the causal effect in each population. That said, with a fitted model, SuSiEx estimates a population-specific causal probability for each credible set, which quantifies whether the signal is causal in a given population. While this probability can be helpful to reveal the cross-population genetic architecture, we note that it heavily depends on statistical power, which is influenced by multiple factors including discovery GWAS sample size and the frequency and effect size of the causal variant. Therefore, the larger number of credible sets that were inferred as causal in EUR relative to AFR and EAS populations (14,115 vs. 725 and 5,699) in our biobank analysis does not necessarily imply that the majority of identified causal variants are specific to Europeans. As non-European datasets continue to expand, we expect increasing evidence supporting that many of these signals are causal in non-European populations.

We recommend using SuSiEx to model population groups separately whenever cross-population differences in causal effect sizes and LD patterns are expected (e.g., when modeling Finnish vs. non-Finnish EUR populations) to avoid inflated false positive rates and power loss. Fixed-effect meta-analysis should only be used when GWAS are conducted in independent samples from the same population where effect sizes and LD patterns are highly concordant. That said, in reality, GWAS summary statistics and LD reference panels for subpopulations are often not available, in which case a meta-analysis followed by a single-population fine-mapping method remains to be a reasonable alternative when the subgroups that constitute a GWAS study are reasonably genetically close.

Throughout this work we tried to use in-sample LD reference panels for fine-mapping because mismatch between the LD of the discovery sample and the reference panel may produce spurious credible sets and causal signals, especially in genomic loci that harbor strong association signals. This has been shown in prior work^37^ and our simulations studies, and is a limitation of all fine-mapping methods. We therefore recommend using in-sample LD for SuSiEx whenever possible, and applying aggressive filtering of low-quality variants and secondary credible sets in complex genomic loci if external LD reference panels have to be used. We also note that all fine-mapping efforts heavily rely on data quality and assume that the true underlying causal variants have been captured and all common variants within the locus have been genotyped or imputed with reasonable accuracy.

Kanai et al. showed that PIP can be miscalibrated due to heterogeneities in the study design^42^. SiSuEx was able to model many heterogeneous factors across studies and estimate PIP more accurately as we have shown in simulations. However, as all fine-mapping methods, SuSiEx relies on high quality input data. Often, the challenge for multi-ancestry fine-mapping stems from the heterogeneity in the array types, imputation pipelines and reference panels across cohorts and studies. Simulations showed that the heterogeneity in the imputation pipeline and reference panel has a dominating impact on the quality of fine-mapping compared with the differences in array types^42^. Managing this heterogeneity is feasible through a single state-of-the-art imputation pipeline with a diverse and well-powered reference panel. Currently, the best publicly available resource is TOPMed^43^, although there remains much room for improvement. We also recommend reviewing each locus for quality issues, either manually or through tools such as SLALOM^42^. For example, if a locus misses a large proportion of common variants, it should not be included in fine-mapping. We expect that data quality issues will be mitigated as the size and diversity of the imputation reference panel continue to increase and low-pass whole-genome sequencing becomes increasingly affordable and rapidly adopted.

There are several limitations of the SuSiEx method and the present study. First, we restricted our analyses to SNPs to avoid potential strand flippings and alignment errors when analyzing indels across biobanks. This may produce false positives if fine-mapped SNP(s) are proxies for causal indels or structural variations (SV). Second, we did not incorporate functional annotations into SuSiEx. Adding functional priors to the model may improve fine-mapping resolution when multiple variants in strong LD have similar statistical significance, and may aid prioritization of follow-up functional studies. That said, the biology underlying the observed variant-phenotype association may be complex, and the modeling of functional data may be error-prone. Extending the Bayesian framework of SuSiEx to leverage functional or other omics data by introducing a proper prior to the model can be a promising future direction. Third, our cross-population fine-mapping in biobanks had an encouraging but modest improvement over the resolution of credible sets identified by European-only analyses, which was largely due to the limited discovery sample size of the African GWAS. However, we have shown that the largest improvements of SuSiEx come with the most diverse datasets, and thus expect that SuSiEx will become increasingly useful as the scale of genomic research in underrepresented populations continues to grow in global biobanks^44^ and disease-focused consortia. Lastly, it remains unclear how SuSiEx performs in admixed samples, in which the local ancestry (and thus the causal variants and their effect sizes) may vary from individual to individual. Developing and evaluating statistical fine-mapping methods in populations with complex genetic ancestries is an important future direction.

In summary, SuSiEx provides robust, accurate and scalable fine-mapping that integrates GWAS summary statistics from diverse populations. Together with the ability to distinguish multiple causal variants within a genomic region, SuSiEx enables the analysis of large, complex genomic loci and aids the interpretation of fine-mapping results. Future work that combines SuSiEx with the rapidly expanding non-European genomic resources may facilitate the discovery of functionally-important disease-causing variants computationally and experimentally.

## METHODS

### The Cross-population Sum of Single Effect (SuSiEx) Model

#### The Cross-population Sum of Single Effects (SuSiEx) model

We extend the “Sum of Single Effects” regression to a cross-population fine-mapping model:

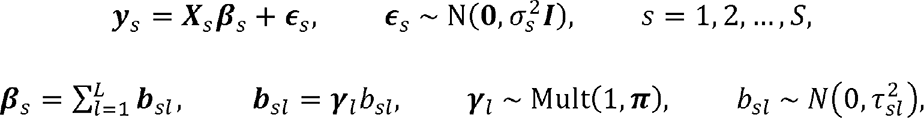

where for population *s* (e.g., European, Asian or African), *y*_s_ is a vector of standardized phenotypes (zero mean and unit variance) from *N_s_* individuals, *X_s_* = [*x*_*s*1_, *x_s_*_2_, …, *x_sM_*] is an *N_s_ ×M* matrix of standardized genotypes (each column *x_sj_* is mean centered and has unit variance) in a genomic region that harbors at least one strong association signal, β_s_ is a vector of SNP effect sizes, and ε_*s*_ is a vector of residuals with i.i.d. elements, each following a normal distribution with zero mean and variance 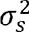. We assume that the overall effect vector β*_s_* is the sum of *L* single-effect vectors *b_s_*_1_, *l* = 1, 2,…, *L*, each has exactly one non-zero element with effect *b_sl_*. The position of the non-zero element is determined by the binary vector *γ_l_*, which follows a multinomial distribution. π = [*π*_1_, *π*_2_, …,*π*_3_]^T^ is a vector that gives the prior probability of a SNP being causal, and 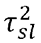 is the prior variance on the effect size *b_sl_* of the causal SNP. We note that all populations share the same underlying causal SNPs (i.e., *γ_l_* does not depend on *s*), but the effect sizes of the causal SNP are allowed to vary across populations (i.e., *b_sl_* depends on *s*).

The SuSiEx model can be fitted using an extension of the iterative Bayesian stepwise selection (IBSS) algorithm, which is equivalent to a coordinate ascent algorithm that maximizes the evidence lower bound (ELBO) of a variational approximation to the SuSiEx model. Both the IBSS algorithm and the ELBO can be computed using GWAS summary statistics, and thus the SuSiEx mode can be fitted without access to any individual-level data (Supplementary Note). For each single-effect *l* = 1, 2, …, *L*, SuSiEx estimates the posterior inclusion probability (PIP) for each variant *α*_*l*_ = [*α*_*l*1_, *α*_*l2*_, …, *α*_*lM*_]^T^, which can be used to compute a level-ρ credible set CS(*α*_l_; ρ) that has a probability no less than ρ of containing at least one causal variant. When L exceeds the number of detectable effects in the data, some *α*_*l*_ become diffuse and the corresponding credible sets will be large, containing many weakly SNPs. Such credible sets have no inferential value and can be discarded if they have purity below a threshold (e.g., 0.5), where purity is defined as the smallest absolute correlation among all pairs of variants within the credible set. Given a converged model fit, SuSiEx additionally estimates a population-specific causal probability for each identified credible set. We use a probability threshold of 0.8 to produce a binarized inference of whether a fine-mapped signal is causal in a given population.

#### The multi-step model fitting approach

To determine the maximum number of single effects *L*, we designed a heuristic, multi-step model fitting approach. Specifically, we start with *L* = 5 and fit the SuSiEx model. If the model does not converge, we sequentially reduce *L* by 1 until the algorithm converges. If the model converges with *L* = 5 and returns 5 credible sets, suggesting that more than 5 credible sets may exist, we set *L* = 10 and rerun the model fitting algorithm. If the model does not converge with *L* = 10, we sequentially reduce *L* by 1 until the algorithm converges.

### Simulations

#### Genomic data

We simulated individual-level genotypes of EUR, EAS and AFR populations using HAPGEN2^19^ with ancestry-matched 1000 Genomes Project (1KG) Phase III^20^ superpopulation samples as the reference panel. We grouped CEU, IBS, FIN, GBR and TSI into the EUR superpopulation, CDX, CHB, CHS, JPT and KHV into the EAS superpopulation, and ESN, MSL, LWK, GWD and YRI into the AFR superpopulation. To calculate the genetic map (cM) and recombination rate (cM/Mb) for each superpopulation, we downloaded the maps and rates for their constituent subpopulations (Data availability), linearly interpolated the genetic map and recombination rate at each position (Code availability), and averaged the genetic maps and recombination rates across the subpopulations in each superpopulation. We simulated 400,000 EUR samples, 200,000 EAS samples and 200,000 AFR samples, and confirmed that the allele frequencies and LD patterns of the simulated genotypes were highly similar to those of the 1KG reference panels. We randomly selected 100 1Mb regions from chromosome 1 (Supplementary Table 1), and filtered for bi-allelic common (MAF >1%) SNPs in at least one of the three superpopulations. To inform the optimal strategy to integrate data from population groups that are genetically close, we additionally simulated samples from subpopulations within a continent (EUR: CEU and FIN; AFR: YRI and LWK; EAS: CHB and JPT) using HAPGEN2 and 1KG subpopulation data as the reference.

#### Phenotypic data

We randomly selected *n_csl_* causal variants within each genomic locus. The allelic effect sizes of each selected causal variant for the EUR, EAS and AFR populations were generated under a multivariate normal distribution N(**0**, Σ_3✕3_), where Σ_3✕3_ was defined as, Σ***_ij_*** *= 1*, if *i* = *j*, and Σ***_ij_*** *= r_g_,* if *i* ≠ *j*, where *r_g_* is the genetic correlation between populations. For each locus, we then generated the phenotype by adding a normally distributed noise term to the genetic component to produce the given per-locus heritability *h*^2^.

#### Simulation settings

To assess SuSiEx in a wide range of settings, we generated simulation data with varying numbers of causal variants (*n_csl_*) per locus, genetic correlations (*r_g_*), and per-locus SNP heritability (*h*^2^). We defined a standard simulation setting using *n_csl_* = 1, *r_g_* = 0.7 and *h*^2^ = 0.1%. We then varied *r_g_* (*r_g_* = 0.4 and 1.0) to reflect different levels of cross-population genetic correlations, varied *h*^2^ (*h*^2^ = 0.05%, 0.2%, 0.3%, 0.4% and 0.5%) to reflect different per-locus heritability values, and varied *n_csl_* (*n_csl_* = 2, 3, 4, 5) with *h*^2^ = 0.5% to reflect the scenario of multiple causal variants in a genomic locus. For each parameter setting, we replicated the simulation five times for each locus (Supplementary Table 2), producing a total of 500 simulated datasets.

To evaluate the robustness of SuSiEx to model misspecification, we simulated causal variants that had non-zero effect sizes in EUR but were null in AFR, and vice versa, and assessed the impact of adding null data on the performance of SuSiEx. We further simulated a scenario that mimicked an African-specific effect in real-world applications, where a variant was only causal in the smaller AFR sample (N=50K) and was null in both EUR and EAS populations with much larger sample sizes (N=500K and 100K, respectively). We also evaluated the robustness of SuSiEx to different values of the hyperparameter (prior variance of each single-effect) in the model.

To assess whether SuSiEx can accurately infer whether a signal is causal in a given population, we simulated different causal configurations (i.e., population-specific or shared causal variants) in two-population and three-population settings across different numbers of causal variants, cross-population genetic correlations, per-locus SNP heritability and GWAS sample sizes. In each setting, we calculated the proportion that SuSiEx can correctly infer the true causal configuration across simulation replicates, and examined how this accuracy was influenced by the allele frequency and allelic effect size of the causal variant.

#### Association analysis and LD calculation

We used the linear regression implemented in PLINK^45^ to generate GWAS summary statistics, and calculated in-sample LD for each genomic locus. To evaluate the impact of LD mismatch on fine-mapping results, we additionally calculated LD matrices using 1KG subpopulation samples within the EUR and AFR superpopulations.

#### Fine-mapping analysis

We compared different fine-mapping methods, including SuSiEx, SuSiE, PAINTOR, MsCAVIAR and the single-population combining method using the standard simulation setting. SuSiEx and SuSiE were performed and evaluated on additional settings beyond the standard simulations. As PAINTOR and MsCAVIAR are not computationally scalable to full GWAS summary statistics, we restricted the analysis to three filtered sets of variants: “p < 0.05”, “top 500” and “top 150”, corresponding to marginal *P* <0.05, the top 500 and the top 150 most associated variants from GWAS, respectively. PAINTOR provides two model fitting options, “MCMC” and “enumerate”. The “MCMC” mode automatically learns the number of causal variants in a locus while the “enumerate” mode requires predefining the maximum number of causal variants. We ran PAINTOR using “-mcmc”, “-enumerate=1”, “-enumerate=2” and “-enumerate=3”. All other parameters were set to default. We set the maximum runtime to 24 hours in our high-performance computing (HPC) system, the maximum memory to 8Gb, and the number of CPUs to one. We ran MsCAVIAR with the default parameters and set the confidence level of credible sets to 95%. For SuSiEx, we used the multi-step model fitting approach described above to determine the number of causal variants. Credible sets that did not contain any genome-wide significant variant (marginal *P* <5E-8) in any single-population GWAS or fixed-effect cross-population meta-GWAS were filtered out. The single-population combining method combines 95% credible sets inferred by SuSiE within each population separately using a hierarchical clustering algorithm. Specifically, for each pair of credible set, the PIP-weighted Jaccard similarity index was computedc as ∑*_i_* ■■■(■*_i_,* ■*_i_*) / ∑*_i_* ■■■(■*_i_,* ■*_i_*), where *x_i_* and *y_i_* are PIP values (or zero if missing) for the same variant *i* from the two credible sets. Pairs of credible sets with a similarity index greater than 0.1 were combined. If one credible set can be combined with multiple credible sets, the set with the highest similarity was selected^7^.

### Biobank analysis

#### Cohorts

GWAS summary statistics of 25 quantitative traits, available from both the UK Biobank (UKBB) and Taiwan Biobank (TWB), were used in the biobank fine-mapping analysis (Supplementary Table 19). European (EUR; *N*_EUR_ up to 419,807) and African (AFR; *N*_AFR_ up to 6,570) GWAS summary statistics were obtained from the Pan-ancestry genetic analysis of the UK Biobank (Pan-UKBB). East Asian GWAS summary statistics were obtained from the Taiwan Biobank (EAS; *N*_EAS_ up to 92,615).

#### Loci definition

We used a 6-way LD clumping-based method to define genomic loci, using 1KG data as the LD reference for clumping. CEU, GBR, TSI, FIN and IBS were combined as the reference for the EUR population; ESN, GWD, LWK, MSL and YRI were combined as the reference for the AFR population; CHB, CHS, CDX, JPT and KHV were combined as the reference for the EAS population. We extracted all variants with MAF >0.5%, and for each of the 25 traits, performed the LD clumping in the three populations using the corresponding reference panel and PLINK^45^. To include loci that reached genome-wide significance (*P* <5E-8) only in the meta-analysis, we further performed clumping for the meta-GWAS across the three populations, using the three reference panels, respectively. For each clumping, we set the p-value threshold of the leading variant as 5E-8 (--clump-p1) and the threshold of the tagging variant as 0.05 (--clump-p2), and set the LD threshold as 0.1 (--clump-r2) and the distance threshold as 250 kb (--clump-kb). We then took the union of the 6-way LD clumping results and extended the boundary of each merged region by 100 kb upstream and downstream. Finally, we merged adjacent loci if the LD (*r*^2^) between the leading variants was larger than 0.6 in any LD reference panel. To account for potential heterogeneity of causal effect sizes across populations, we also performed a random-effect meta-analysis across the three populations using METASOFT^46^. This method, followed by the same clumping procedure, did not identify additional loci for fine-mapping.

#### In-sample LD calculation

We used the in-sample LD of the three populations in the fine-mapping analysis. We extracted all variants with MAF >0.5% from each population and calculated the LD using PLINK^45^. Multi-allelic variants and indels were excluded to avoid potential strand flipping and alignment errors.

#### Fine-mapping

We applied SuSiEx to the 25 quantitative traits to integrate GWAS summary statistics derived from the three populations, and examined 99% credible sets. In both single-population and cross-population fine-mapping, we filtered out credible sets that did not contain any genome-wide significant variant (*P* <5E-8) in any population-specific GWAS or cross-population fixed-effect meta-GWAS.

#### Credible set alignment

To compare the results between single-population and cross-population fine-mapping, we aligned the inferred credible sets across the four sets of analyses using the same clustering algorithm based on the PIP-weighted Jaccard similarity index^7^ described above.

#### Functional annotations

The functional impact of each variant was annotated using VEP, with the definition and classification of functional impact obtained from https://useast.ensembl.org/info/genome/variation/prediction/predicted_data.html. The high impact category includes transcript ablation, splice acceptor variants and splice donor variants; the moderate impact category includes missense variants and protein-altering variants; the low impact category includes synonymous variants and splice region variants; the modifier impact category includes introns and intergenic variants among others.

### Cross-population fine-mapping in schizophrenia cohorts

Schizophrenia GWAS summary statistics of European (EUR; *N_case_* = 53,251, *N_control_* = 77,127) and East Asian (EAS; *N_case_*= 14,004, *N_control_* = 16,757) ancestries were obtained from the recently published Psychiatric Genomics Consortium (PGC) schizophrenia analysis^4^. We fine-mapped the same 250 autosomal loci defined in the PGC publication. We calculated LD by applying LD-Store v1.1^37^ to each cohort and locus, and then calculated an effective sample size weight LD matrix^47^ across cohorts for the EUR and EAS populations, respectively (Code availability; LDmergeFM). We applied SuSiEx to integrate EUR and EAS schizophrenia GWAS summary statistics to perform cross-population fine-mapping. Credible set level was set to 99%. Credible sets that did not contain any genome-wide significant variant (marginal *P* <5E-8) in single-population GWAS or cross-population meta-GWAS were filtered out.

## Data Availability

Publicly available data are available from the following sites: 1KG Phase 3 reference panels: https://mathgen.stats.ox.ac.uk/impute/1000GP_Phase3.html.
Genetic map for each subpopulation: https://ftp.1000genomes.ebi.ac.uk/vol1/ftp/technical/working/20130507_omni_recombination_rates. Individual-level genotypes for UKBB samples were obtained under application 32568. UKBB European and African GWAS summary statistics were obtained from the PanUKBB Project. https://pan.ukbb.broadinstitute.org/downloads.
The GWAS summary statistics and in-sample LD of European and Asian for the schizophrenia analysis were obtained from the Schizophrenia Working Group of the Psychiatric Genomic Consortium (PGC). https://figshare.com/articles/dataset/scz2022/19426775. Taiwan Biobank data used in this study contain protected health information and are thus under controlled access. Application to access such data can be made to the Taiwan Biobank. https://taiwanview.twbiobank.org.tw/data_appl.

## DATA AVAILABILITY

Publicly available data are available from the following sites:

1KG Phase 3 reference panels: https://mathgen.stats.ox.ac.uk/impute/1000GP_Phase3.html;

Genetic map for each subpopulation: https://ftp.1000genomes.ebi.ac.uk/vol1/ftp/technical/working/20130507_omni_recombination_rates;

PanUKBB summary statistics: https://pan.ukbb.broadinstitute.org/downloads;

TWB data used in this study contain protected health information and are thus under controlled access. Application to access such data can be made to the TWB (https://www.twbiobank.org.tw/new_web_en/);

PGC schizophrenia GWAS: https://pgc.unc.edu/for-researchers/download-results

## CODE AVAILABILITY

The code used in this study is available from the following websites: SuSiEx: https://github.com/getian107/SuSiEx;

PAINTOR: https://github.com/gkichaev/PAINTOR_V3.0;

MsCAVIAR: https://github.com/nlapier2/MsCAVIAR;

HAPGEN2: https://mathgen.stats.ox.ac.uk/genetics_software/hapgen/hapgen2.html;

PLINK1.9: https://www.cog-genomics.org/plink;

LDmergeFM: https://github.com/Pintaius/LDmergeFM;

METASOFT: http://genetics.cs.ucla.edu/meta

## ETHICS

Collection of the UKBB data was approved by the Research Ethics Committee of the UKBB. UKBB individual-level data used in the present work were obtained under application 32568. Collection of the TWB data was approved by the Ethics and Governance Council (EGC) of TWB and the Department of Health and Welfare, Taiwan (Wei-Shu-I-Tzu no.1010267471). TWB obtained informed consent from all participants for research use of the collected data. Access to, and use of, TWB data in the present work was approved by the EGC of TWB (approval number: TWBR10907-05) and the Institutional Review Board of National Health Research Institutes, Taiwan (approval number: EC1090402-E).

## ACKNOWLEDGMENTS

We thank Masahiro Kanai for helpful discussions. UKBB European and African GWAS summary statistics were obtained from the PanUKBB Project. We thank the Schizophrenia Working Group of the Psychiatric Genomic Consortium (PGC) for providing the GWAS summary statistics and in-sample LD for the schizophrenia analysis. H.H. acknowledges supports from National Institute of Diabetes and Digestive and Kidney Diseases (NIDDK) K01DK114379 and R01DK129364, National Institute of Mental Health (NIMH) U01MH109539 and R01MH130675, Brain and Behavior Research Foundation Young Investigator Grant (28450), the Zhengxu and Ying He Foundation, and the Stanley Center for Psychiatric Research. T.G. is supported by National Human Genome Research Institute (NHGRI) R01HG012354. Y.F.L. is supported by the National Health Research Institutes (NP-109-PP-09), and the Ministry of Science and Technology (109-2314-B-400-017) of Taiwan.

## AUTHOR CONTRIBUTIONS

T.G. and H.H. designed the project; T.G. developed the statistical methods. K.Y. and T.G. programmed the code for SuSiEx; K.Y., R.J.L. and T.G. conducted simulation studies; K.Y., R.J.L. and M.Y. performed the biobank fine-mapping analysis; T.T.C., S.C.L., Y.A.F., Y.F.L. and C.Y.C. performed the analysis in the Taiwan Biobank; K.Y. and A.F.P. performed the analysis in the schizophrenia cohorts; M.J.D. and B.M.N. provided critical suggestions for the study design; M.Y., Y.C., M.L., R.L. and Y.X. took part in the testing of the code; M.O.D. and Z.G. made significant contributions to the generation and management of schizophrenia data. K.Y., T.G. and H.H. wrote the manuscript; All the authors reviewed and approved the final version of the manuscript.

---

**Extended Data Figure 1:**
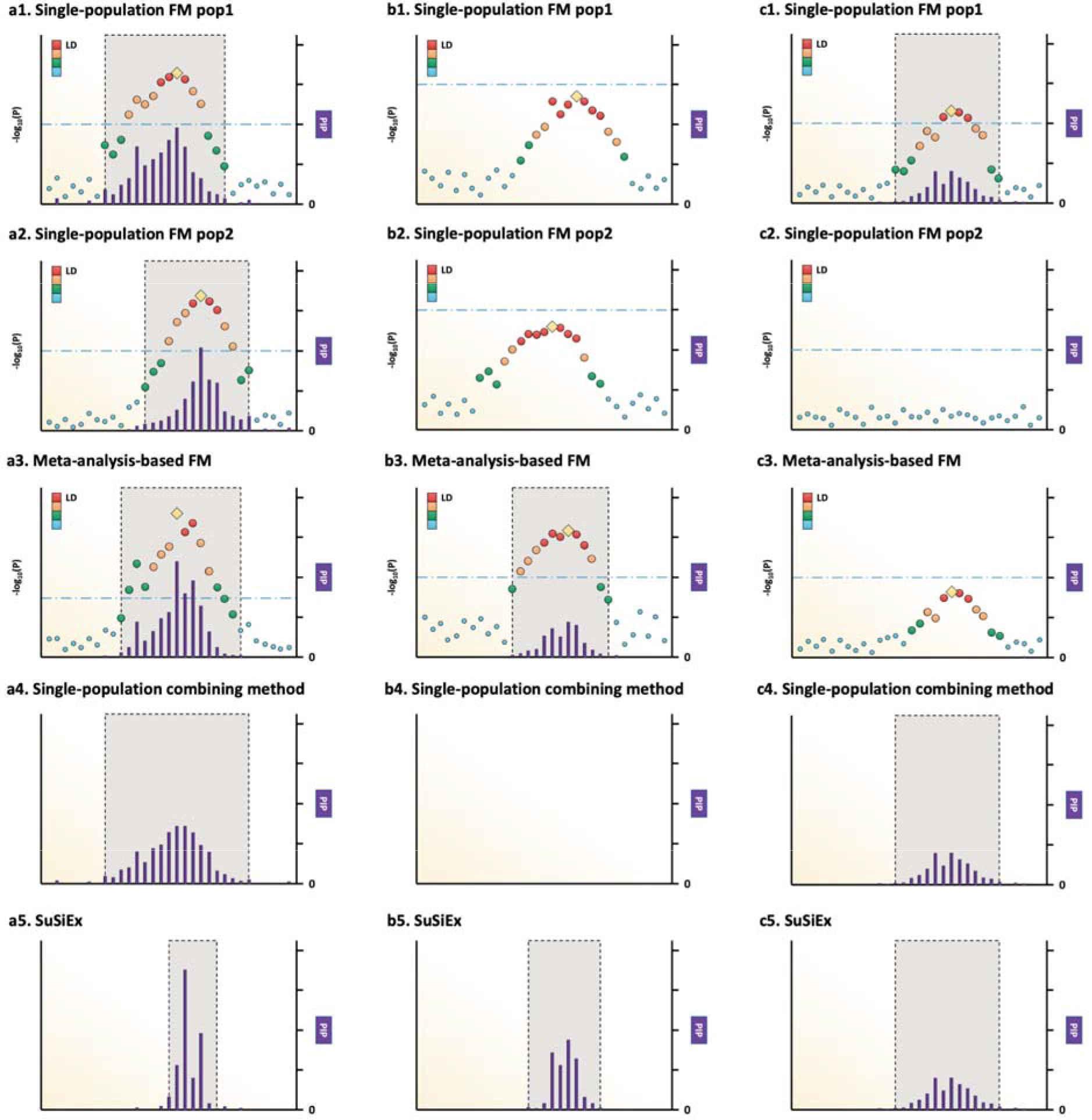
Schematic illustration of the meta-analysis-based fine-mapping method, single-population combining method, and SuSiEx. All panels were created following the LocusZoom style^17^. Variant positions are shown on the x axis. The gold diamond for each locus represents the lead (most associated) variant. The association strengths for other variants are colored by descending degrees of linkage disequilibrium (LD) with the lead variant (ordered red, orange, green, and blue dots). The purple bars represent the posterior inclusion probabilities (PIPs) inferred by fine-mapping methods. The light gray boxes represent the credible sets estimated by fine-mapping. **a1-a5**, Example of a strong causal signal shared across populations. **b1-b5**, Example of a weak causal signal shared across populations. **c1-c5**, Example of a population-specific causal signal.

**Extended Data Figure 2:**
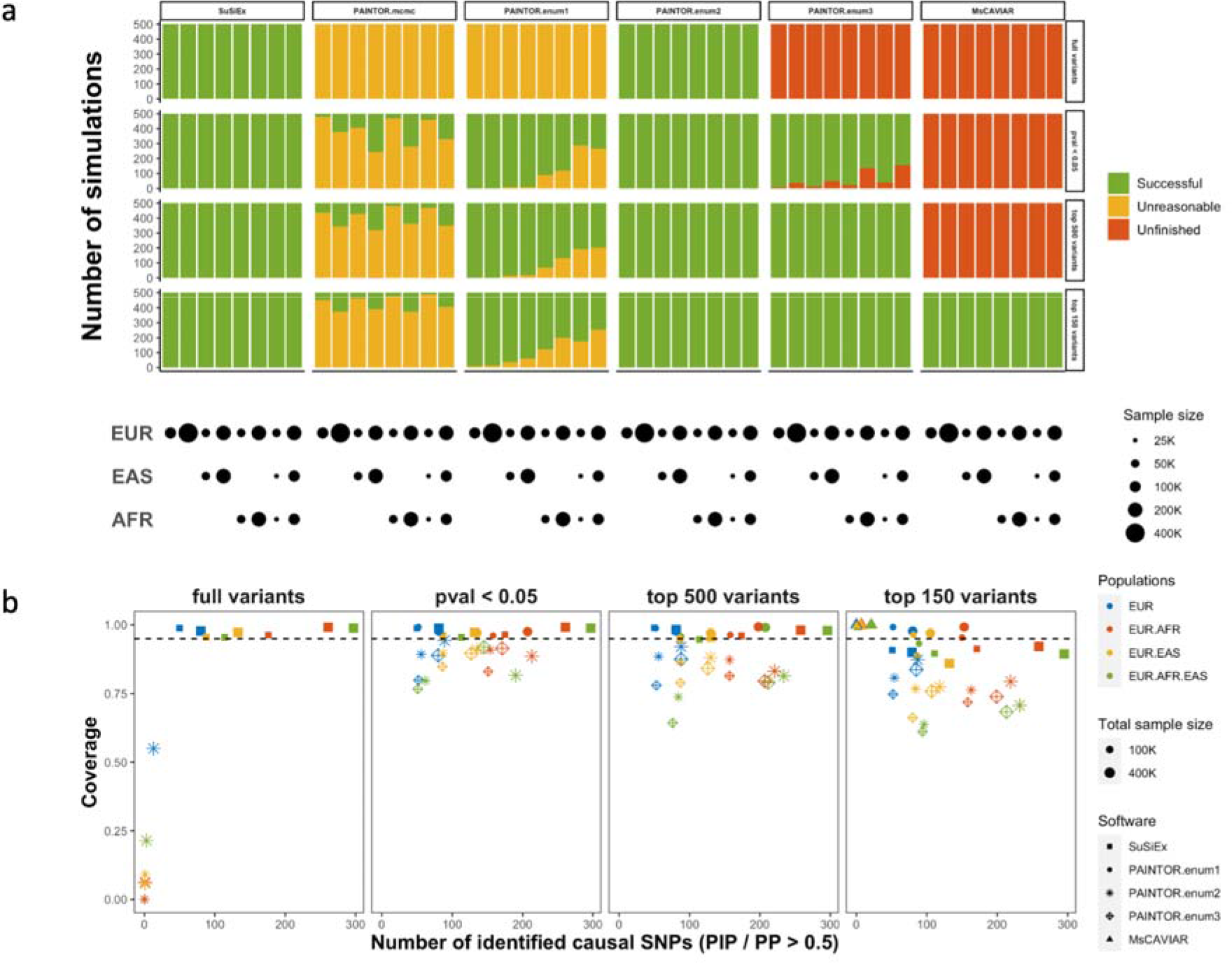
Comparison of SuSiEx, PAINTOR and MsCAVIAR in simulations. **a**, The job completion summary for the three Bayesian fine-mapping methods using different parameters and input datasets. Red represents jobs taking longer than 24 hours. Yellow represents jobs returning unreasonable results, defined as the sum of PIPs across variants in the genomic locus >5 or <0.1 (1 is expected). Green represents jobs that were completed within 24 hours and returned reasonable results. The lower panel represents different sample size combinations of the discovery GWAS. **b**, Number of identified true causal SNPs with PIP >0.5 (x-axis) versus the coverage of the credible sets (y-axis) for different input datasets and fine-mapping methods. Color represents the combination of discovery populations, the size of the symbols represents the total discovery sample size, and the shape of the symbols represents different methods and parameters. Only simulation runs that were completed within 24 hours and returned reasonable results were included.

**Extended Data Figure 3:**
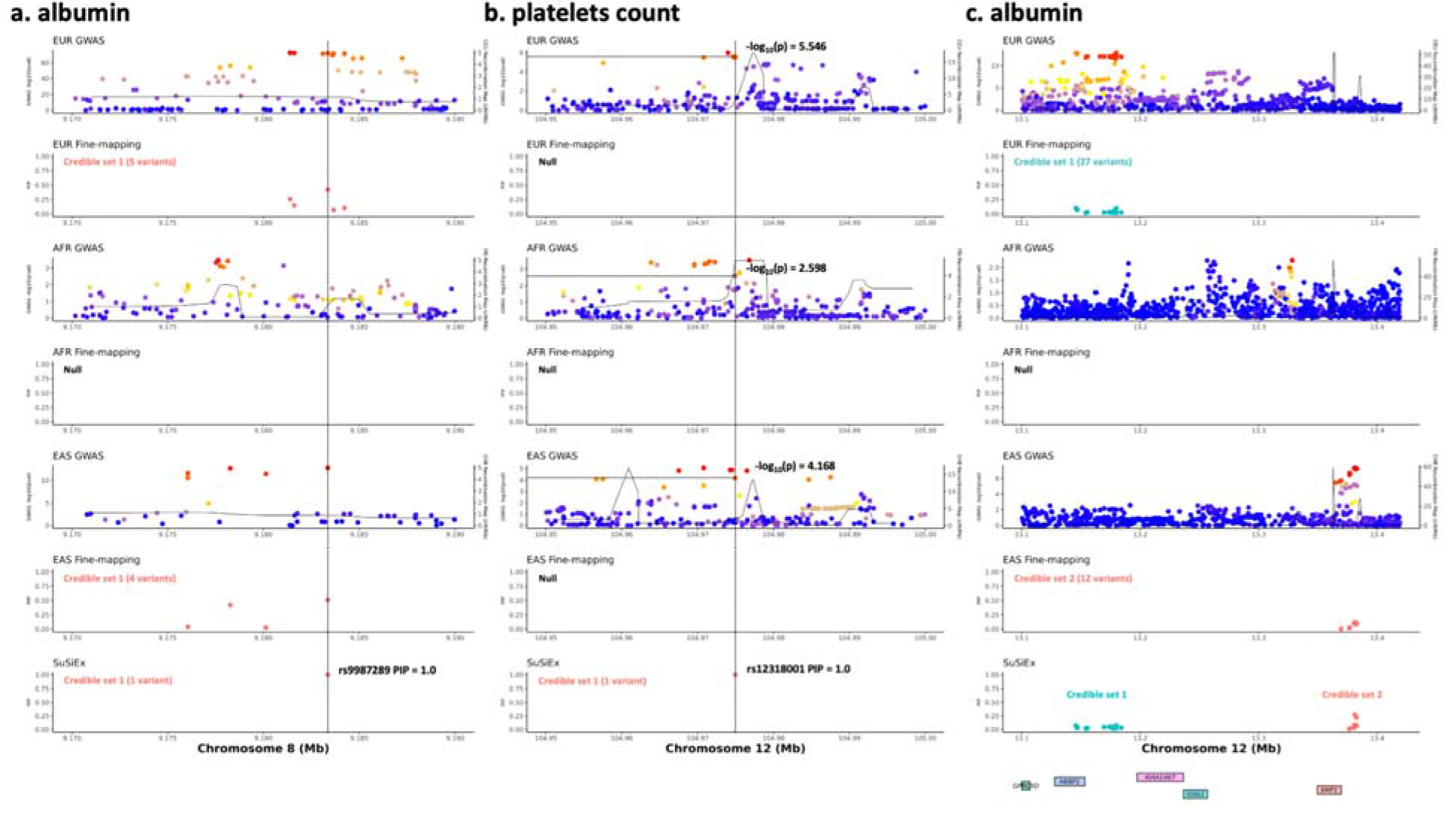
Examples of the improvement of SuSiEx over single-population fine-mapping in the biobank analysis. Each of the three sub-figures consists of eight panels, which are aligned vertically, with the x-axis showing the genomic position. The top six panels visualize GWAS association statistics and single-population fine-mapping results within the European (Pan-UKBB Europan), African (Pan-UKBB African) and East Asian (Taiwan biobank) populations. For association statistics, the left y-axis shows the -log_10_(p-value) of each SNP. The color represents the descending degrees of LD with the lead SNP (from red, orange to blue). The right y-axis shows the recombination rate in centimorgan per Megabase. The solid line indicates the population-specific recombination maps obtained from the 1000 Genomes Project. Different colors are used to distinguish different credible sets in the fine-mapping results. The second to bottom panel visualizes the results from SuSiEx. “Null” indicates that single-population fine-mapping did not obtain any reliable credible set. The bottom panel shows gene annotations if any. **a**, Association with albumin on chr8:9,170,000-9,190,000, an example of a strong causal signal shared across populations. **b**, Association with platelets count on chr12:104,900,000-105,050,000, an example of a weak causal signal shared across populations. **c**, Association with albumin on chr12:13,100,000-13,400,000, an example of population-specific causal signals.

**Extended Data Figure 4:**
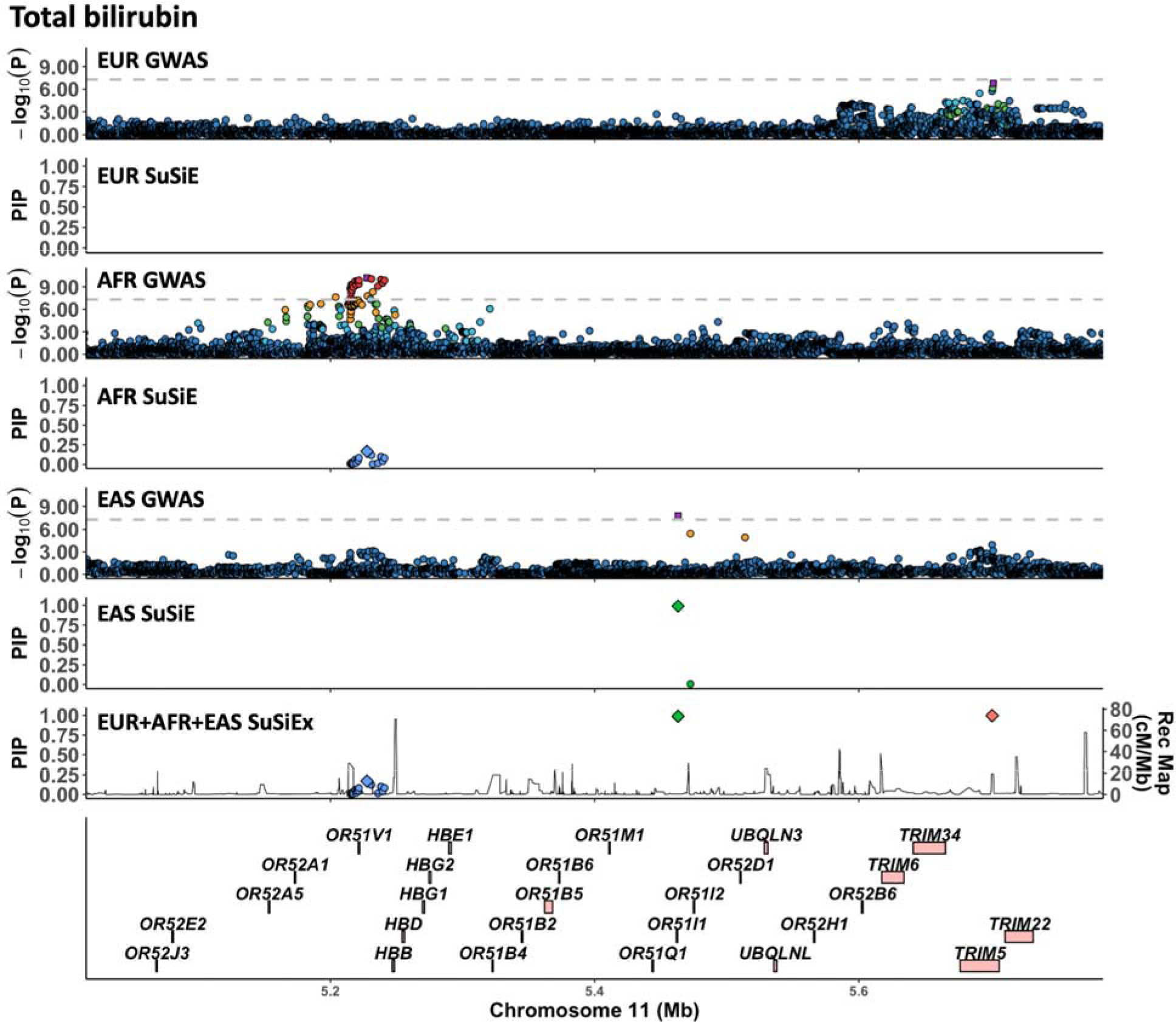
Association with total bilirubin on chr11: 5,100,000-5,700,000. Panels are aligned vertically, with the x-axis showing the genomic position. The top six panels visualize GWAS association statistics and single-population fine-mapping results of the European (Pan-UKBB Europan), African (Pan-UKBB African) and East Asian (Taiwan biobank) populations following the LocusZoom^37^ style. The second to bottom panel visualizes the fine-mapping results from SuSiEx, which integrated GWAS summary statistics from the three populations. The bottom panel shows gene annotations. For GWAS panels, the left y-axis shows the -log_10_(p-value) of each SNP. The gray horizontal dash line represents the genome-wide significance threshold (5E-8). The purple rectangle for each locus represents the lead (most associated) variant. Variants are colored by descending LD with the lead variant (ordered red, orange, green, light blue, and dark blue dots). For fine-mapping panels, different colors are used to distinguish different credible sets. The diamond represents the variant with the maximum PIP in each credible set. The left y-axis shows the PIP from fine-mapping and the right y-axis shows the recombination map obtained from the 1000 Genomes Project. For the SuSiEx panel, the average recombination rate across three populations is used.

**Extended Data Figure 5:**
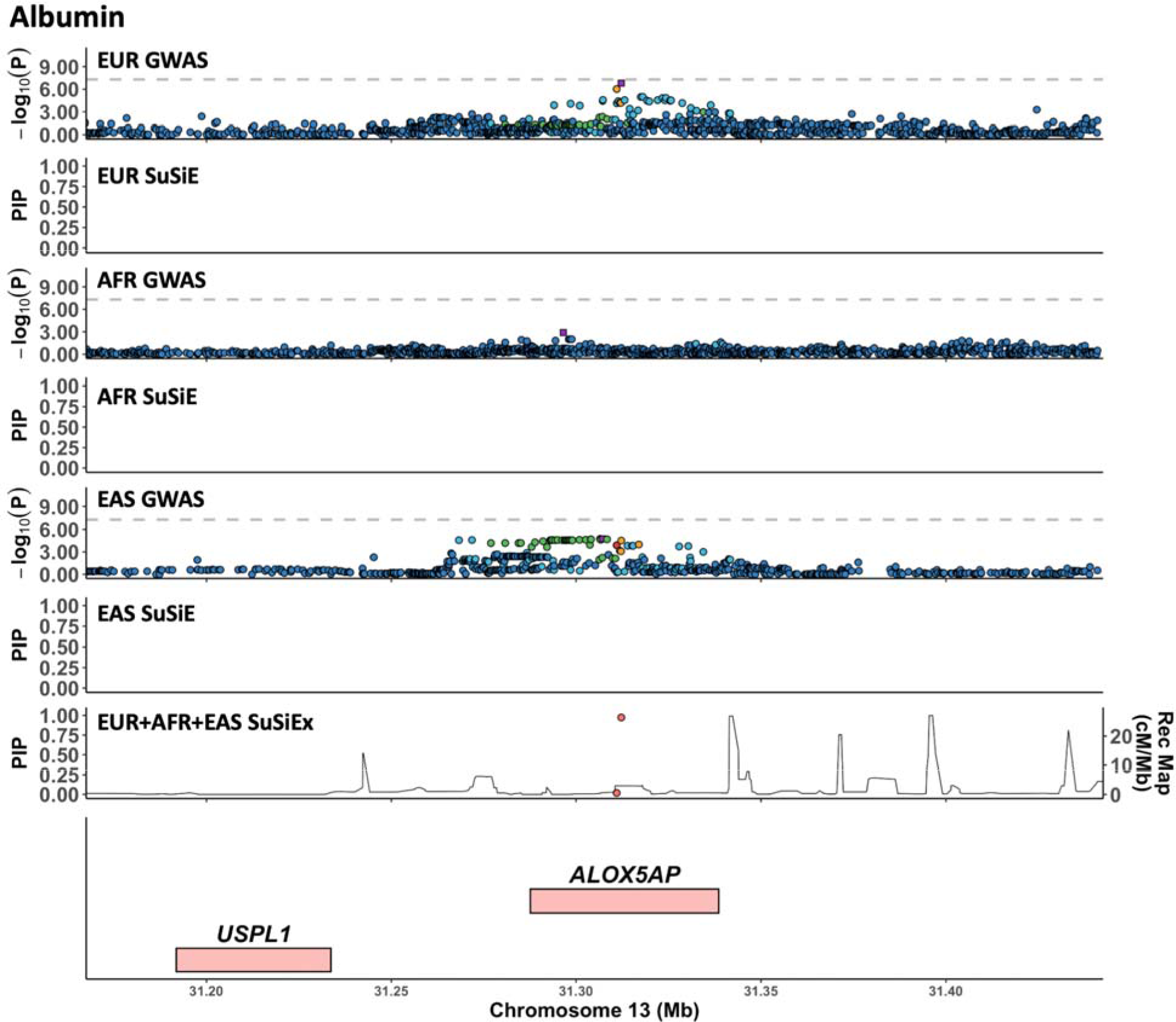
Association with albumin on chr13: 31,150,000-31,450,000. Panels are aligned vertically, with the x-axis showing the genomic position. The top six panels visualize GWAS association statistics and single-population fine-mapping results of the European (Pan-UKBB Europan), African (Pan-UKBB African) and East Asian (Taiwan biobank) populations following the LocusZoom^37^ style. The second to bottom panel visualizes the fine-mapping results from SuSiEx, which integrated GWAS summary statistics from the three populations. The bottom panel shows gene annotations. For GWAS panels, the left y-axis shows the -log_10_(p-value) of each SNP. The gray horizontal dash line represents the genome-wide significance threshold (5E-8). The purple rectangle for each locus represents the lead (most associated) variant. Variants are colored by descending LD with the lead variant (ordered red, orange, green, light blue, and dark blue dots). For fine-mapping panels, different colors are used to distinguish different credible sets. The diamond represents the variant with the maximum PIP in each credible set. The left y-axis shows the PIP from fine-mapping and the right y-axis shows the recombination map obtained from the 1000 Genomes Project. For the SuSiEx panel, the average recombination rate across three populations is used.

**Extended Data Figure 6:**
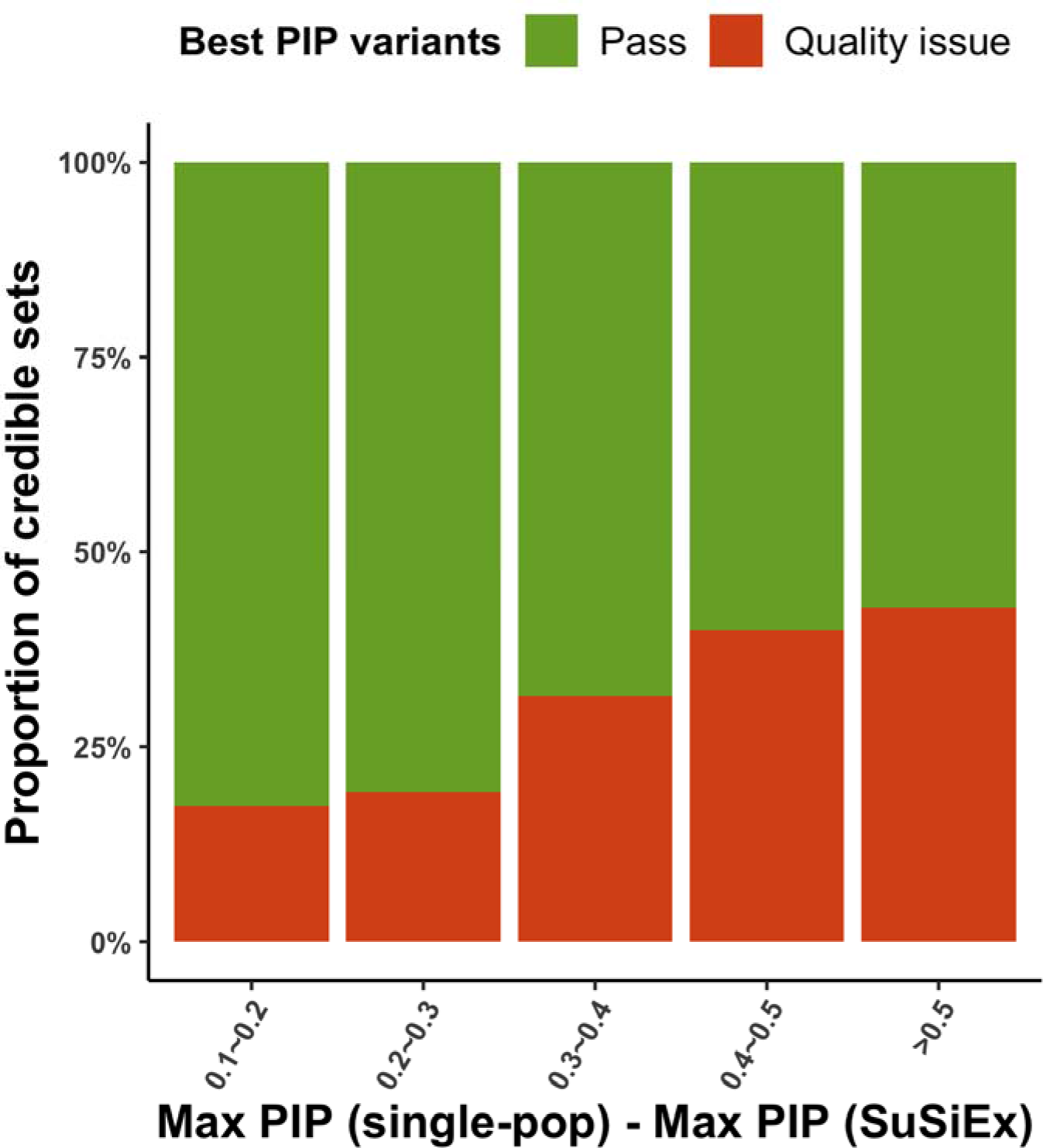
Proportion of variants showing quality issues binned by the drop in PIP between single- and multi-population fine-mapping. Quality issues were defined as (i) the best PIP variant is in the low complexity region; (ii) the best PIP variant is in allelic imbalance or violates Hardy Weinberg equilibrium in gnomAD^33^; or (iii) the best PIP variant is multi-allelic or colocalizes with indels at the same genomic position, which might influence imputation quality.

**Extended Data Figure 7:**
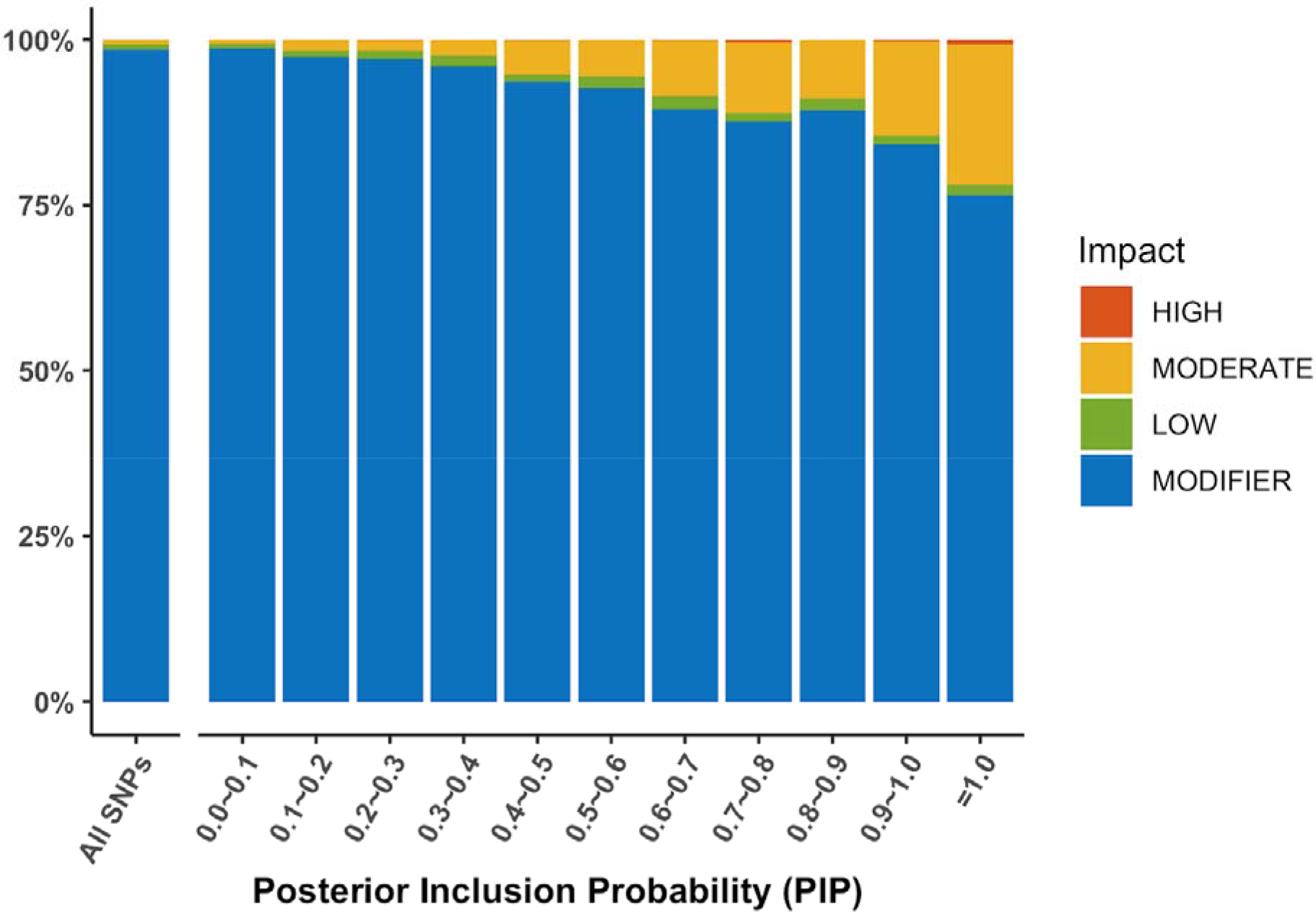
Proportion of variants with high/moderate functional impact in cross-population biobank fine-mapping analyses. The functional impact of each variant was annotated using VEP, with the definition and classification of functional impact obtained from https://useast.ensembl.org/info/genome/variation/prediction/predicted_data.html. The high impact category includes transcript ablation, splice acceptor variants and splice donor variants; the moderate impact category includes missense variants and protein-altering variants; the low impact category includes synonymous variants and splice region variants; the modifier impact category includes introns and intergenic variants among others.

