## Supplementary Notes for "Fine-mapping across diverse ancestries drives the discovery of putative causal variants underlying human complex traits and diseases"

**The Cross-population Sum of Single Effect (SuSiEx) Model**

**The multiple-population single effect regression (SER) model.** We first describe the SER model for multiple populations, which is the building block for the SuSiEx model. Consider the following linear regression:

$$\boldsymbol{y}_s = \boldsymbol{X}_s \boldsymbol{b}_s + \boldsymbol{\epsilon}_s, \qquad \boldsymbol{\epsilon}_s \sim \mathrm{N}(\boldsymbol{0}, \sigma_s^2 \boldsymbol{I}), \qquad s = 1, 2, \ldots, S,$$

$$\boldsymbol{b}_s = b_s \boldsymbol{\gamma}, \qquad \boldsymbol{\gamma} \sim \mathrm{Mult}(1, \boldsymbol{\pi}), \qquad b_s \sim \mathrm{N}(0, \tau_s^2),$$

where for population $s$ (e.g., European, Asian or African), $\boldsymbol{y}_s$ is a vector of standardized phenotypes (zero mean and unit variance) from $N_s$ individuals, $\boldsymbol{X}_s = [\boldsymbol{x}_{s1}, \boldsymbol{x}_{s2}, \ldots, \boldsymbol{x}_{sM}]$ is an $N_s \times M$ matrix of standardized genotypes (each column $\boldsymbol{x}_{sj}$ is mean centered and has unit variance) in a genomic region that harbors at least one strong association signal, $\boldsymbol{b}_s$ is a vector of SNP effect sizes, and $\boldsymbol{\epsilon}_s$ is a vector of residuals with i.i.d. elements, each following a normal distribution with zero mean and variance $\sigma_s^2$. The vector $\boldsymbol{b}_s$ has exactly one non-zero element with effect $b_s$. The position of the non-zero element is determined by the binary vector $\boldsymbol{\gamma}$, which follows a multinomial distribution. $\boldsymbol{\pi} = [\pi_1, \pi_2, \ldots, \pi_M]^{\mathrm{T}}$ is a vector that gives the prior probability of a SNP being causal, and $\tau_s^2$ is the prior variance on the effect size $b_s$ of the causal SNP. We note that all populations share the same underlying causal SNPs (i.e., $\boldsymbol{\gamma}$ does not depend on $s$), but the effect sizes of the causal SNP are allowed to vary across populations (i.e., $b_s$ depends on $s$).

**Posterior under the multi-population SER.** Conditional on the $j$-th SNP being causal, the least squares estimate of its effect size in population $s$ is $\hat{b}_{sj} = \left(\boldsymbol{x}_{sj}^{\mathrm{T}} \boldsymbol{x}_{sj}\right)^{-1} \boldsymbol{x}_{sj}^{\mathrm{T}} \boldsymbol{y}_s = N_s^{-1} \boldsymbol{x}_{sj}^{\mathrm{T}} \boldsymbol{y}_s$, with variance $v_{sj}^2 = \sigma_s^2 \left(\boldsymbol{x}_{sj}^{\mathrm{T}} \boldsymbol{x}_{sj}\right)^{-1} = \sigma_s^2 N_s^{-1}$ and the corresponding $z$ score $z_{sj} = \hat{b}_{sj} / v_{sj}$. The Bayes Factor (BF) for comparing this model with the null model (i.e., $b_s = 0$) in population $s$ is:

$$\mathrm{BF}_{sj} = \mathrm{BF}\left(\boldsymbol{y}_s, \boldsymbol{x}_{sj}\right) = \frac{p(\boldsymbol{y}_s \mid \boldsymbol{x}_{sj}, \sigma_s^2, \tau_s^2)}{p(\boldsymbol{y}_s \mid b_s = 0, \sigma_s^2)} = \sqrt{\frac{v_{sj}^2}{\tau_s^2 + v_{sj}^2}} \exp\left(\frac{z_{sj}^2}{2} \frac{v_{sj}^2}{\tau_s^2 + v_{sj}^2}\right).$$

Let $\boldsymbol{y} = \{\boldsymbol{y}_s\}_{s=1}^S$, $\boldsymbol{\sigma}^2 = \{\sigma_s^2\}_{s=1}^S$ and $\boldsymbol{\tau}^2 = \{\tau_s^2\}_{s=1}^S$, the collection of $\boldsymbol{y}_s$, $\sigma_s^2$ and $\tau_s^2$ across populations, the posterior distribution on $\boldsymbol{b}_s = b_s \boldsymbol{\gamma}$ can be computed as:

$$\boldsymbol{\gamma} \mid \boldsymbol{y}, \boldsymbol{\sigma}^2, \boldsymbol{\tau}^2 \sim \mathrm{Mult}(1, \boldsymbol{\alpha}),$$

$$b_s \mid \boldsymbol{y}_s, \sigma_s^2, \tau_s^2, \gamma_j = 1 \sim \mathrm{N}\left(\mu_{sj}, \phi_{sj}^2\right),$$

where $\boldsymbol{\alpha} = [\alpha_1, \alpha_2, \ldots, \alpha_M]^{\mathrm{T}}$ is a vector of posterior inclusion probabilities (PIPs) with

$$\alpha_j = p(\gamma_j = 1 \mid \boldsymbol{y}, \boldsymbol{\sigma}^2, \boldsymbol{\tau}^2) = \frac{\prod_{s=1}^S p(\boldsymbol{y}_s \mid \gamma_j = 1) p(\gamma_j = 1)}{\sum_{j'=1}^M \prod_{s=1}^S p\left(\boldsymbol{y}_s \mid \gamma_{j'} = 1\right) p\left(\gamma_{j'} = 1\right)}$$

$$= \frac{\pi_j \prod_{s=1}^S p(\boldsymbol{y}_s \mid \boldsymbol{x}_{sj})}{\sum_{j'=1}^M \pi_{j'} \prod_{s=1}^S p(\boldsymbol{y}_s \mid \boldsymbol{x}_{sj'})} = \frac{\pi_j \prod_{s=1}^S \mathrm{BF}(\boldsymbol{y}_s, \boldsymbol{x}_{sj})}{\sum_{j'=1}^M \pi_{j'} \prod_{s=1}^S \mathrm{BF}(\boldsymbol{y}_s, \boldsymbol{x}_{sj'})} = \frac{\pi_j \prod_{s=1}^S \mathrm{BF}_{sj}}{\sum_{j'=1}^M \pi_{j'} \prod_{s=1}^S \mathrm{BF}_{sj'}}$$

$$\phi_{sj}^2 = \left(v_{sj}^{-2} + \tau_s^{-2}\right)^{-1}, \qquad \mu_{sj} = \left(\frac{\phi_{sj}^2}{v_{sj}^2}\right)\hat{b}_{sj}.$$

The likelihood of the multi-population SER model is:

$$\ell_{\mathrm{SER}}(\boldsymbol{y}; \boldsymbol{\sigma}^2, \boldsymbol{\tau}^2) = \sum_{j=1}^M \prod_{s=1}^S p(\boldsymbol{y}_s \mid \gamma_j = 1)\, p(\gamma_j = 1) = \prod_{s=1}^S p_0(\boldsymbol{y}_s \mid \sigma_s^2) \sum_{j=1}^M \pi_j \prod_{s=1}^S \mathrm{BF}_{sj},$$

where $p_0(\boldsymbol{y}_s \mid \sigma_s^2) = p(\boldsymbol{y}_s \mid b_s = 0, \sigma_s^2) = (2\pi\sigma_s^2)^{-\frac{N_s}{2}} \exp\left(-\frac{1}{2\sigma_s^2} \boldsymbol{y}_s^{\mathsf{T}} \boldsymbol{y}_s\right)$.

**The Cross-population Sum of Single Effects (SuSiEx) model.** We extend the multi-population SER model to the SuSiEx model that allows for multiple causal effects in a genomic locus:

$$\boldsymbol{y}_s = \boldsymbol{X}_s \boldsymbol{\beta}_s + \boldsymbol{\epsilon}_s, \qquad \boldsymbol{\epsilon}_s \sim \mathrm{N}(\boldsymbol{0}, \sigma_s^2 \boldsymbol{I}), \qquad s = 1, 2, \dots, S,$$

$$\boldsymbol{\beta}_s = \sum_{l=1}^L \boldsymbol{b}_{sl}, \qquad \boldsymbol{b}_{sl} = \boldsymbol{\gamma}_l b_{sl}, \qquad \boldsymbol{\gamma}_l \sim \mathrm{Mult}(1, \boldsymbol{\pi}), \qquad b_{sl} \sim N(0, \tau_{sl}^2),$$

where the overall effect vector $\boldsymbol{\beta}_s$ is the sum of $L$ single-effect vectors $\boldsymbol{b}_{sl}$, $l = 1, 2, \dots, L$, each has exactly one non-zero element.

**Variational approximation to the SuSiEx model.** We use variational approximation to fit the SuSiEx model. Assuming that $\tau_{sl}^2$ is fixed, the evidence lower bound (ELBO) of the model is:

$$F(\boldsymbol{\sigma}^2, q; \boldsymbol{y}) = \log \ell(\boldsymbol{y}; \boldsymbol{\sigma}^2, \boldsymbol{\tau}^2) - D_{KL}(q \mid\mid p_{\mathrm{post}}),$$

where $\ell(\boldsymbol{y}; \boldsymbol{\sigma}^2, \boldsymbol{\tau}^2)$ is the likelihood of the SuSiEx model,

$$D_{KL}(q \mid\mid p) = \int q(\boldsymbol{\beta}) \log \frac{q(\boldsymbol{\beta})}{p(\boldsymbol{\beta})} \mathrm{d}\boldsymbol{\beta}$$

is the Kullback-Leibler (KL) divergence between the two distributions $p$ and $q$, and $p_{\mathrm{post}} = p(\boldsymbol{\beta} \mid \boldsymbol{y})$ is the posterior distribution of the causal effects. We fit the model by maximizing the ELBO $F$ over a class of distributions $Q$ that factorize over the $L$ single effects: $q(\boldsymbol{b}_{s1}, \boldsymbol{b}_{s2}, \dots, \boldsymbol{b}_{sL}) = \prod_{l=1}^L q_l(\boldsymbol{b}_{sl})$ for any $q$ in $Q$. Let $\mathrm{E}_q[\cdot]$ denote the expectation with respect to the distribution $q$,

$$F = \log[p(\boldsymbol{y})] - \mathrm{E}_q\left[\log \frac{q(\boldsymbol{\beta})}{p(\boldsymbol{\beta} \mid \boldsymbol{y})}\right] = \mathrm{E}_q\left[\log \frac{p(\boldsymbol{y}) p(\boldsymbol{\beta} \mid \boldsymbol{y})}{q(\boldsymbol{\beta})}\right] = \mathrm{E}_q[\log\, p(\boldsymbol{y} \mid \boldsymbol{\beta})] + \mathrm{E}_q\left[\log \frac{p(\boldsymbol{\beta})}{q(\boldsymbol{\beta})}\right]$$

$$= -\sum_{s=1}^{S} \frac{N_s}{2} \log(2\pi\sigma_s^2) - \sum_{s=1}^{S} \frac{1}{2\sigma_s^2} \mathrm{E}_q \left\| \boldsymbol{y}_s - \sum_{l=1}^{L} \boldsymbol{\eta}_{sl} \right\|^2 + \sum_{l=1}^{L} \mathrm{E}_{q_l} \left[ \log \frac{p_l(\boldsymbol{\eta}_l)}{q_l(\boldsymbol{\eta}_l)} \right]$$

$$= -\sum_{s=1}^{S} \frac{N_s}{2} \log(2\pi\sigma_s^2) - \sum_{s=1}^{S} \frac{1}{2\sigma_s^2} \mathrm{ERSS}_s + \sum_{l=1}^{L} \mathrm{E}_{q_l} \left[ \log \frac{p_l(\boldsymbol{\eta}_l)}{q_l(\boldsymbol{\eta}_l)} \right]$$

where ||·|| denotes the Euclidean norm, and $\mathrm{ERSS}$ is the expected residual sum of squares under the variational approximation $q$, $\boldsymbol{\eta}_{sl} = \boldsymbol{X}_s \boldsymbol{b}_{sl}$ and $\boldsymbol{\eta}_l = \{\boldsymbol{\eta}_{sl}\}_{s=1}^{S}$. We optimize $F$ over $q$ and $\sigma_s^2$ by iteratively updating each $q_l, l = 1, 2, \dots, L$, while keeping $\sigma_s^2$ and other elements of $q$ fixed, along with a separate step for updating $\sigma_s^2$ with all elements of $q_l$ fixed. Specifically, to update $\sigma_s^2$, we note that

$$\frac{\partial F}{\partial \sigma_s^2} = -\frac{N_s}{2\sigma_s^2} + \frac{1}{2\sigma_s^4} \mathrm{ERSS}_s.$$

Setting this derivative to zero and solving for $\sigma_s^2$ gives $\hat{\sigma}_s^2 = \mathrm{ERSS}_s / N_s$.

To update $q_l$ with $q_{l'}, l' \neq l$ fixed, we have

$$F = -\sum_{s=1}^{S} \frac{N_s}{2} \log(2\pi\sigma_s^2) - \sum_{s=1}^{S} \frac{1}{2\sigma_s^2} \mathrm{E}_q \|\boldsymbol{r}_{sl} - \boldsymbol{\eta}_{sl}\|^2 + \mathrm{E}_{q_l} \left[ \log \frac{p_l(\boldsymbol{\eta}_l)}{q_l(\boldsymbol{\eta}_l)} \right] + \mathrm{const}$$

$$= -\sum_{s=1}^{S} \frac{N_s}{2} \log(2\pi\sigma_s^2) - \sum_{s=1}^{S} \frac{1}{2\sigma_s^2} \mathrm{E}_{q_l} \|\bar{\boldsymbol{r}}_{sl} - \boldsymbol{\eta}_{sl}\|^2 + \mathrm{E}_{q_l} \left[ \log \frac{p_l(\boldsymbol{\eta}_l)}{q_l(\boldsymbol{\eta}_l)} \right] + \mathrm{const},$$

where $\mathrm{const}$ denotes terms that do not depend on $q_l$, $\boldsymbol{r}_{sl} = \boldsymbol{y}_s - \sum_{l' \neq l} \boldsymbol{\eta}_{sl}$, and $\bar{\boldsymbol{r}}_{sl} = \mathrm{E}_q[\boldsymbol{y}_s - \sum_{l' \neq l} \boldsymbol{\eta}_{sl}] = \boldsymbol{y}_s - \sum_{l' \neq l} \bar{\boldsymbol{\eta}}_{sl} = \boldsymbol{y}_s - \sum_{l' \neq l} \boldsymbol{X}_s \bar{\boldsymbol{b}}_{sl'}$ is the expected residual under $q_{l'}, l' \neq l$. We note that maximizing $F$ with $q_{l'}, l' \neq l$ fixed is equivalent to maximizing the ELBO for the multi-population SER model in which the observed phenotypes $\boldsymbol{y}_s$ are replaced by the expected residual $\bar{\boldsymbol{r}}_{sl}$. Further, the optimization of the SER model does not restrict the form of $q_l$ and has a closed form solution. Therefore, the SuSiEx model can be fitted using an extension of the iterative Bayesian stepwise selection (IBSS) algorithm.

**The iterative Bayesian stepwise selection (IBSS) algorithm.** With an initialization of the posterior mean effect size of $\boldsymbol{b}_{sl}$, denoted as $\bar{\boldsymbol{b}}_{sl}$ (e.g., $\bar{\boldsymbol{b}}_{sl} = 0$ for all $s$ and $l$), the fitting procedure updates the posterior of $\boldsymbol{b}_{sl}$, given estimates of other effects $\boldsymbol{b}_{sl'}, l' \neq l$, and then updates $\sigma_s^2$ with all estimates of $\boldsymbol{b}_{sl}$ fixed, until convergence:

- Compute the expected residuals:

$$\bar{\boldsymbol{r}}_{sl} = \boldsymbol{y}_s - \sum_{l' \neq l} \boldsymbol{X}_s \bar{\boldsymbol{b}}_{sl'}, \qquad s = 1, 2, \dots, S.$$

- Compute the posterior inclusion probabilities (PIPs):

$$\alpha_{lj} = p\big(\gamma_{lj} = 1 \mid \bar{\boldsymbol{r}}_{sl}, \boldsymbol{\sigma}^2, \boldsymbol{\tau}^2\big) = \frac{\pi_j \prod_{s=1}^{S} \mathrm{BF}(\bar{\boldsymbol{r}}_{sl}, \boldsymbol{x}_{sj})}{\sum_{j'=1}^{M} \pi_{j'} \prod_{s=1}^{S} \mathrm{BF}(\bar{\boldsymbol{r}}_{sl}, \boldsymbol{x}_{sj'})}, \qquad j = 1, 2, \dots, M,$$

where

$$\mathrm{BF}(\bar{\boldsymbol{r}}_{sl}, \boldsymbol{x}_{sj}) = \sqrt{\frac{v_{sj}^2}{\tau_{sl}^2 + v_{sj}^2}} \exp\left(\frac{z_{slj}^2}{2} \frac{v_{sj}^2}{\tau_{sl}^2 + v_{sj}^2}\right),$$

$$\hat{b}_{slj} = \left(\boldsymbol{x}_{sj}^{\mathsf{T}} \boldsymbol{x}_{sj}\right)^{-1} \boldsymbol{x}_{sj}^{\mathsf{T}} \bar{\boldsymbol{r}}_{sl} = N_s^{-1} \boldsymbol{x}_{sj}^{\mathsf{T}} \bar{\boldsymbol{r}}_{sl},\ v_{sj}^2 = \sigma_s^2 \left(\boldsymbol{x}_{sj}^{\mathsf{T}} \boldsymbol{x}_{sj}\right)^{-1} = \sigma_s^2 N_s^{-1},\ z_{slj} = \hat{b}_{slj} / v_{sj}.$$

- Update the posterior distribution for $b_{sl}$:

$$b_{sl} \mid \bar{\boldsymbol{r}}_{sl}, \sigma_s^2, \tau_s^2, \gamma_{lj} = 1 \sim N\big(\mu_{slj}, \phi_{slj}^2\big), \qquad \phi_{slj}^2 = \left(v_{sj}^{-2} + \tau_{sl}^{-2}\right)^{-1}, \qquad \mu_{slj} = \left(\frac{\phi_{slj}^2}{v_{sj}^2}\right) \hat{b}_{slj}.$$

- Update the posterior moments for $\boldsymbol{b}_{sl}$:

$$\bar{\boldsymbol{b}}_{sl} = \mathrm{E}[\boldsymbol{b}_{sl}] = \boldsymbol{\alpha}_l \circ \boldsymbol{\mu}_{sl}, \qquad \bar{\boldsymbol{b}}_{sl}^2 = \mathrm{E}[\boldsymbol{b}_{sl}^2] = \boldsymbol{\alpha}_l \circ (\boldsymbol{\mu}_{sl}^2 + \boldsymbol{\phi}_{sl}^2),$$

where $\boldsymbol{\alpha}_l = [\alpha_{l1}, \alpha_{l2}, \dots, \alpha_{lM}]^{\mathsf{T}}$, $\boldsymbol{\mu}_{sl} = [\mu_{sl1}, \mu_{sl2}, \dots, \mu_{slM}]^{\mathsf{T}}$, $\boldsymbol{\phi}_{sl} = [\phi_{sl1}, \phi_{sl2}, \dots, \phi_{slM}]^{\mathsf{T}}$, and $\circ$ is element-wise multiplication.

- Update $\sigma_s^2$:

$$\sigma_s^2 = \frac{1}{N_s} \mathrm{ERSS}_s.$$

**Model fitting with summary statistics.** To fit the SuSiEx model and calculate the ELBO $F$ to monitor the convergence of the IBSS algorithm using only GWAS summary statistics, we denote $\hat{\beta}_{sj} = \left(\boldsymbol{x}_{sj}^{\mathsf{T}} \boldsymbol{x}_{sj}\right)^{-1} \boldsymbol{x}_{sj}^{\mathsf{T}} \boldsymbol{y}_s = N_s^{-1} \boldsymbol{x}_{sj}^{\mathsf{T}} \boldsymbol{y}_s$ as the marginal least squares effect size estimate of the $j$-th SNP in populations $s$, and $\boldsymbol{D}_s = [\boldsymbol{d}_{s1}, \boldsymbol{d}_{s2}, \dots, \boldsymbol{d}_{sM}] = \boldsymbol{X}_s^{\mathsf{T}} \boldsymbol{X}_s / N_s$ as the LD matrix for population $s$, and note that when computing the expected residuals in the IBSS algorithm, we have:

$$\boldsymbol{x}_{sj}^{\mathsf{T}} \bar{\boldsymbol{r}}_{sl} = \boldsymbol{x}_{sj}^{\mathsf{T}} \boldsymbol{y}_s - \boldsymbol{x}_{sj}^{\mathsf{T}} \sum_{l' \neq l} \boldsymbol{X}_s \bar{\boldsymbol{b}}_{sl'} = N_s \hat{\beta}_{sj} - N_s \sum_{l' \neq l} \boldsymbol{d}_{sj}^{\mathsf{T}} \bar{\boldsymbol{b}}_{sl'}.$$

Therefore, instead of $\bar{\boldsymbol{r}}_{sl}$, we update $\boldsymbol{x}_{sj}^{\mathsf{T}} \bar{\boldsymbol{r}}_{sl}$ in each iteration, which can be calculated from summary statistics and can be used to update $\hat{b}_{slj} = N_s^{-1} \boldsymbol{x}_{sj}^{\mathsf{T}} \bar{\boldsymbol{r}}_{sl}$. In addition, we have

$$\mathrm{ERSS}_s = \mathrm{E}_q \left\| \boldsymbol{y}_s - \sum_{l=1}^{L} \boldsymbol{\eta}_{sl} \right\|^2 = \boldsymbol{y}_s^{\mathsf{T}} \boldsymbol{y}_s - 2 \sum_{l=1}^{L} \overline{\boldsymbol{\eta}}_{sl}^{\mathsf{T}} \boldsymbol{y}_s + \sum_{l,k=1}^{L} \mathrm{E}_q [\boldsymbol{\eta}_{sl}^{\mathsf{T}} \boldsymbol{\eta}_{sk}]$$

$$= \boldsymbol{y}_s^{\mathsf{T}} \boldsymbol{y}_s - 2 \sum_{l=1}^{L} \overline{\boldsymbol{b}}_{sl}^{\mathsf{T}} \boldsymbol{X}_s^{\mathsf{T}} \boldsymbol{y}_s + \sum_{l,k=1}^{L} \overline{\boldsymbol{b}}_{sl}^{\mathsf{T}} \boldsymbol{X}_s^{\mathsf{T}} \boldsymbol{X}_s \overline{\boldsymbol{b}}_{sk} - \sum_{l=1}^{L} \overline{\boldsymbol{b}}_{sl}^{\mathsf{T}} \boldsymbol{X}_s^{\mathsf{T}} \boldsymbol{X}_s \overline{\boldsymbol{b}}_{sl} + \sum_{l=1}^{L} \mathrm{E}_{q_l} [\boldsymbol{b}_{sl}^{\mathsf{T}} \boldsymbol{X}_s^{\mathsf{T}} \boldsymbol{X}_s \boldsymbol{b}_{sl}]$$

$$= N_s - 2N_s \sum_{l=1}^{L} \overline{\boldsymbol{b}}_{sl}^{\mathsf{T}} \widehat{\boldsymbol{\beta}}_s + N_s \sum_{l,k=1}^{L} \overline{\boldsymbol{b}}_{sl}^{\mathsf{T}} \boldsymbol{D}_s \overline{\boldsymbol{b}}_{sk} - N_s \sum_{l=1}^{L} \overline{\boldsymbol{b}}_{sl}^{\mathsf{T}} \boldsymbol{D}_s \overline{\boldsymbol{b}}_{sl} + N_s \sum_{l=1}^{L} \sum_{j=1}^{M} \overline{b}_{slj}^2 .$$

Lastly,

$$\mathrm{E}_{q_l} \left[ \log \frac{p_l(\boldsymbol{\eta}_l)}{q_l(\boldsymbol{\eta}_l)} \right] = \log \ell_{\mathrm{SER}}(\overline{\boldsymbol{r}}_{sl}) + \sum_{s=1}^{S} \frac{N_s}{2} \log(2\pi\sigma_s^2) + \sum_{s=1}^{S} \frac{1}{2\sigma_s^2} \mathrm{E}_{q_l} \| \overline{\boldsymbol{r}}_{sl} - \boldsymbol{\eta}_{sl} \|^2$$

$$= \log \ell_{\mathrm{SER}}(\overline{\boldsymbol{r}}_{sl}) + \sum_{s=1}^{S} \frac{N_s}{2} \log(2\pi\sigma_s^2) + \sum_{s=1}^{S} \frac{1}{2\sigma_s^2} \left[ \overline{\boldsymbol{r}}_{sl}^{\mathsf{T}} \overline{\boldsymbol{r}}_{sl} - 2\overline{\boldsymbol{b}}_{sl}^{\mathsf{T}} \boldsymbol{X}_s^{\mathsf{T}} \overline{\boldsymbol{r}}_{sl} + \mathrm{E}_{q_l} \left( \boldsymbol{b}_{sl}^{\mathsf{T}} \boldsymbol{X}_s^{\mathsf{T}} \boldsymbol{X}_s \boldsymbol{b}_{sl} \right) \right]$$

$$= \log \sum_{j=1}^{M} \pi_j \prod_{s=1}^{S} \mathrm{BF}\left( \overline{\boldsymbol{r}}_{sl}, \boldsymbol{x}_{sj} \right) + \sum_{s=1}^{S} \frac{1}{2\sigma_s^2} \left[ -2\overline{\boldsymbol{b}}_{sl}^{\mathsf{T}} \boldsymbol{X}_s^{\mathsf{T}} \overline{\boldsymbol{r}}_{sl} + N_s \sum_{j=1}^{M} \overline{b}_{slj}^2 \right].$$

Therefore, both the IBSS algorithm and the ELBO $F$ can be computed using GWAS summary statistics. In practice, the IBSS algorithm terminates when the increase in $F$ between successive iterations is smaller than a small non-negative number (e.g., 1e-4).

**Construction of credible sets.** The PIPs $\boldsymbol{\alpha}_l$ can be used to compute a level-$\rho$ credible set $CS(\boldsymbol{\alpha}_l; \rho)$, which has a probability no less than $\rho$ of containing at least one causal SNP. Specifically, let $(i_1, i_2, \dots, i_M)$ denote the indices that sort $\alpha_{lj}$ in decreasing order, i.e., $\alpha_{li_1} > \alpha_{li_2} > \cdots > \alpha_{li_M}$, and let $S_k = \sum_{j=1}^{k} \alpha_{li_j}$. Then $CS(\boldsymbol{\alpha}_l; \rho) \coloneqq \{i_1, i_2, \dots, i_{k_0}\}$, where $k_0 = \min\{k : S_k \geq \rho\}$. When $L$ exceeds the number of detectable effects in the data, some $\boldsymbol{\alpha}_l$ become diffuse and the corresponding credible sets will be large, containing many weakly SNPs. Such credible sets have no inferential value and can be discarded if they have purity below a threshold (e.g., 0.5), where purity is defined as the smallest absolute correlation among all pairs of variants within the credible set.

***Post hoc* probabilities of causal configurations.** After fitting the SuSiEx model, we provide a *post hoc* probability of whether a credible set is shared across populations or population-specific. Let $\boldsymbol{\omega}_{lm} = [\omega_{1lm}, \omega_{2lm}, \dots, \omega_{Slm}]^{\mathsf{T}}, m = 1, 2, \dots, 2^S$ denote a total of $W = 2^S$ activation configurations of the $l$-th credible set, with $\omega_{slm} \in \{0,1\}$ being a binary indicator of whether the signal in the $l$-th credible set is causal in population $s$ under the $m$-th configuration. We assume that the activation indicator vector $\boldsymbol{\omega}_{lm} \in \{0,1\}^S$ follows a multivariate Bernoulli distribution with a uniform prior for each configuration. For a given credible set, the posterior probability of a causal configuration, conditional on the posterior estimates of other credible sets $\overline{\boldsymbol{r}}_l = \{\overline{\boldsymbol{r}}_{sl}\}_{s=1}^{S}$, can be calculated as:

$$p(\boldsymbol{\omega}_{lm} \mid \bar{\boldsymbol{r}}_l) = \frac{\sum_{j=1}^{M} p(\bar{\boldsymbol{r}}_l \mid \boldsymbol{\omega}_{lm}, \gamma_{lj} = 1) p(\gamma_{lj} = 1) p(\boldsymbol{\omega}_{lm})}{\sum_{m'=1}^{W} \sum_{j=1}^{M} p(\bar{\boldsymbol{r}}_l \mid \boldsymbol{\omega}_{lm'}, \gamma_{lj} = 1) p(\gamma_{lj} = 1) p(\boldsymbol{\omega}_{lm'})}$$

$$= \frac{\sum_{j=1}^{M} \prod_{s:\omega_{slm=1}} p(\bar{\boldsymbol{r}}_{sl} \mid \boldsymbol{\omega}_{lm}, \gamma_{lj} = 1) p(\gamma_{lj} = 1)}{\sum_{m'=1}^{W} \sum_{j=1}^{M} \prod_{s:\omega_{slm'=1}} p(\bar{\boldsymbol{r}}_{sl} \mid \boldsymbol{\omega}_{lm'}, \gamma_{lj} = 1) p(\gamma_{lj} = 1)}$$

$$= \frac{\sum_{j=1}^{M} \pi_j \prod_{s:\omega_{slm=1}} \mathrm{BF}(\bar{\boldsymbol{r}}_{sl}, \boldsymbol{x}_{sj})}{\sum_{m'=1}^{W} \sum_{j=1}^{M} \pi_j \prod_{s:\omega_{slm'=1}} \mathrm{BF}(\bar{\boldsymbol{r}}_{sl}, \boldsymbol{x}_{sj})}.$$

The probability of the $l$-th credible set being causal in population $s_0$ can then be calculated as: $\sum_{m:\omega_{s_0lm=1}} p(\boldsymbol{\omega}_{lm} \mid \bar{\boldsymbol{r}}_{sl})$. We use a probability threshold of 0.8 to infer whether a fine-mapped signal is causal or not in population $s_0$.

**Additional modeling considerations.** There are several reasons why we did not explicitly model population specificity of each causal signal in SuSiEx (e.g., by including a binary indicator variable for each causal variant and each population). Our initial perspective was based on the understanding that proving a null hypothesis is inherently unattainable. Specifically, whether a causal signal can be detected in a population (i.e., whether it is population-specific) depends on the statistical power, which is in turn determined by multiple factors including the allele frequency and allelic effect size of the causal variant and the discovery sample size. All these factors vary continuously, and it is technically impossible to distinguish a population-specific causal variant (exactly zero genetic effect) from a population-shared causal variant with a tiny genetic effect. For instance, a causal variant with an effect close to zero can be classified as non-causal in a small study and flip to causal as the discovery sample size increases. Therefore, rather than using a cutoff to make a *binarized* inference of whether a variant is causal or not in a population, we focus on *estimating* its effect in each population, which also brings substantial computational simplicity and efficiency.

The rationale of our modeling assumption lies in the fact that as long as the causal variant has a non-zero (even tiny) effect, it can be considered as shared across populations and we can leverage the shared signal and data from multiple populations to improve the power and resolution of fine-mapping. Modeling the effect size (as a continuous variable) rather than the causal status (as a binary state) of an association signal allows us to take advantage of data from underrepresented populations which is still underpowered at this time. As the frequency and/or allelic effect size of a causal variant becomes smaller in a population (with zero effect on the extreme of this continuous spectrum), the benefits of including data from that population in fine-mapping become smaller, but since we allow the effect size to vary across populations without restrictions and does not penalize small or zero effects, the model remains valid and well-calibrated.

Our last consideration is computational cost. While there are methods developed for the problem of multi-trait colocalization that can explicitly infer the causal configuration across traits (e.g., Foley et al. 2021, Nature Communications; Giambartolomei et al. 2018, Bioinformatics), these methods all assumed that each trait has no more than one causal variant in the genetic locus being analyzed. Since the number of possible causal configurations grows exponentially with both the number of traits (in the multi-trait setting) or populations (in the multi-population setting) and the number of causal variants, existing techniques become computationally impractical when analyzing a genomic region with a handful of causal variants in more than two populations. For example, to fine-map a locus with a maximum of 5 causal variants (which is the default setting in this study) across 3 ancestries, there will be (2^5)^3 = 32768 causal configurations to search through, which is highly computationally expensive even with sophisticated approximation techniques and quickly exhausts the power of non-European datasets.
