## Supplementary Figures for "Fine-mapping across diverse ancestries drives the discovery of putative causal variants underlying human complex traits and diseases"

| Supplementary Figure | Title | Section |
|---|---|---|
| 1 | Performance of SuSiEx under the standard simulation setting. | Standard simulation setting |
| 2 | The number of confidently identified causal SNPs under the standard simulation setting. | |
| 3 | Comparison between SuSiEx and the meta-analysis-based fine-mapping method using the standard simulation setting. | SuSiEx vs. meta-analysis-based fine-mapping, the single-population combining method and meta+SuSiE |
| 4 | The coverage of Meta+SuSiE and SuSiEx when analyzing genetically close populations. | |
| 5 | The number of causal variants identified by Meta+SuSiE and SuSiEx when analyzing genetically close populations with identical causal effect sizes. | |
| 6 | The number of causal variants identified by Meta+SuSiE, SuSiEx and Mega+SuSiE when analyzing two independent samples from the same population. | |
| 7 | Comparison between SuSiEx and the single-population combining method under the standard simulation setting. | |
| 8 | The runtime of SuSiEx under varying sample sizes and population combinations. | The computational efficiency of SuSiEx |
| 9 | The number of iterations before SuSiEx converged under varying sample sizes and population combinations. | |
| 10 | The number of causal SNPs identified by SuSiEx under varying genetic correlations ($r_g$). | Simulation with varying $r_g$ |
| 11 | The coverage of SuSiEx under varying genetic correlations ($r_g$). | |
| 12 | The size of credible sets identified by SuSiEx under varying genetic correlations ($r_g$). | |
| 13 | The maximum PIP estimated by SuSiEx under varying genetic correlations ($r_g$). | |
| 14 | The number of confidently identified causal SNPs under varying genetic correlations ($r_g$). | |
| 15 | The number of causal SNPs identified by SuSiEx under varying local heritability ($h^2$). | Simulation with varying $h^2$ |

| | | |
|---|---|---|
| 16 | The coverage of SuSiEx under varying local heritability ($h^2$). | |
| 17 | The size of credible sets identified by SuSiEx under varying local heritability ($h^2$). | |
| 18 | The maximum PIP estimated by SuSiEx under varying local heritability ($h^2$). | |
| 19 | The number of confidently identified causal SNPs under varying local heritability ($h^2$). | |
| 20 | The number of causal SNPs identified by SuSiEx under varying numbers of causal SNPs per locus ($n_{csl}$). | Simulation with varying $n_{csl}$ |
| 21 | The coverage of SuSiEx under varying numbers of causal SNPs per locus ($n_{csl}$). | |
| 22 | The size of credible sets identified by SuSiEx under varying numbers of causal SNPs per locus ($n_{csl}$). | |
| 23 | The maximum PIP estimated by SuSiEx under varying numbers of causal SNPs per locus ($n_{csl}$). | |
| 24 | The number of confidently identified causal SNPs under varying numbers of causal SNPs per locus ($n_{csl}$). | |
| 25 | The impact of different $\tau_s^2$ values on the performance of SuSiEx. | Simulation with varying hyperparameters |
| 26 | The impact of different $\tau_s^2$ values on the calibration of SuSiEx. | |
| 27 | Performance of SuSiEx in the presence of population-specific causal SNPs. | Simulation of population-specific causal variants |
| 28 | Performance of SuSiEx in the presence of African-specific causal variants. | |
| 29 | The population-specific causal probability under the standard simulation setting. | |
| 30 | The impact of allele frequency and causal effect size on the classification of population-specific causal variants. | |
| 31 | The population-specific causal probability under varying numbers of causal SNPs per locus ($n_{csl}$) in two-population fine-mapping analysis. | |
| 32 | The population-specific causal probability under varying genetic correlations ($r_g$) in two-population fine-mapping analysis. | |

| | | |
|---|---|---|
| 33 | The population-specific causal probability under varying local heritability ($h^2$) in two-population fine-mapping analysis. | |
| 34 | The population-specific causal probability under the standard simulation setting in three-population fine-mapping analysis. | |
| 35 | The impact of allele frequency and causal effect size on the classification of population-specific causal variants in three-population fine-mapping analysis. | |
| 36 | The population-specific causal probability under varying numbers of causal SNPs per locus ($n_{csl}$) in three-population fine-mapping analysis. | |
| 37 | The population-specific causal probability under varying genetic correlations ($r_g$) in three-population fine-mapping analysis. | |
| 38 | The population-specific causal probability under varying local heritability ($h^2$) in three-population fine-mapping analysis. | |
| 39 | Comparison of fine-mapping results between in-sample LD and external reference LD. | In-sample LD vs. external LD |
| 40 | Cross-population fine-mapping analysis after removing the variants with quality issues in biobanks. | Biobank analysis |
| 41 | The marginal per-allele effect size of the maximum PIP variant across populations. | |

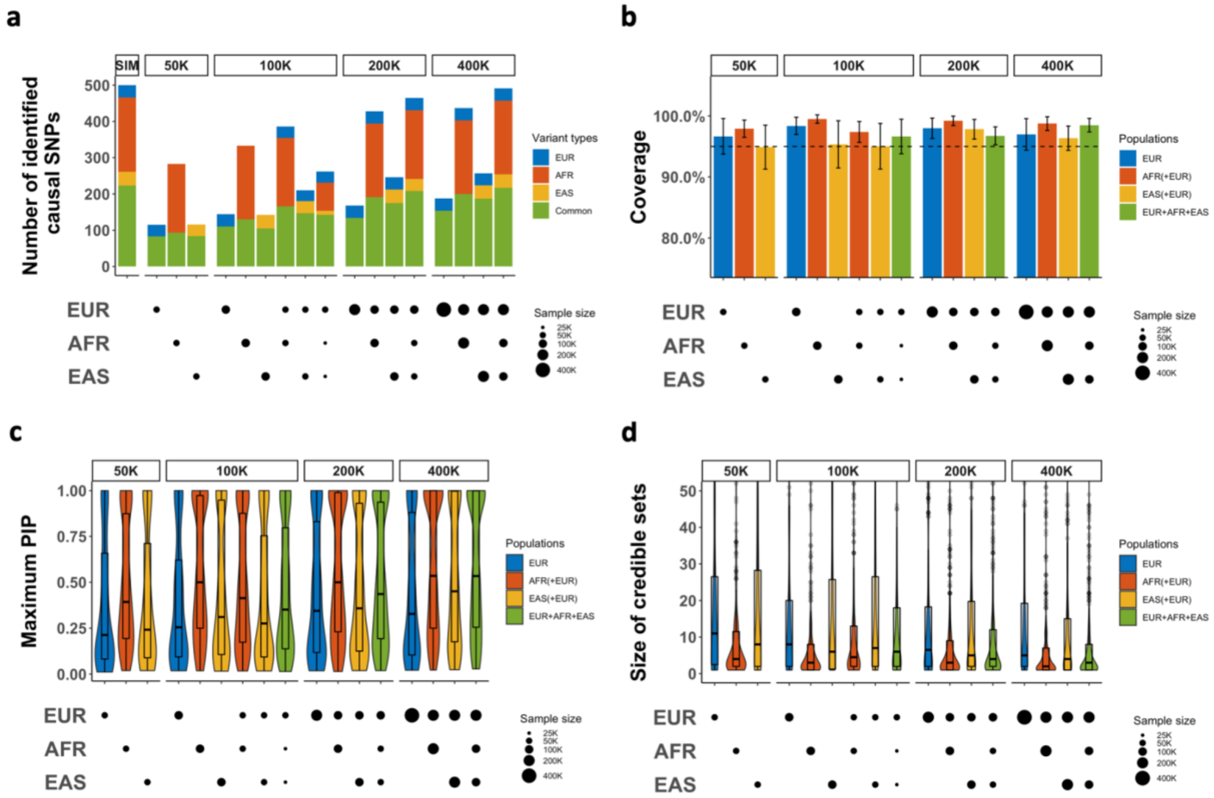


**Supplementary Figure 1: Performance of SuSiEx under the standard simulation setting.** **a**, The number of identified true causal variants when integrating data from different populations with different sample sizes for fine-mapping (true causal variants covered by a credible set). **b**, The coverage of credible sets. The dashed line represents the 95% coverage. The error bar represents the 95% confidence interval. **c**, Distribution of the maximum PIP. **d**, Distribution of the size of credible sets. The upper and lower bounds of the box indicate the 75th and 25th percentiles, respectively. The middle line in the box indicates the median. The top label of each subpanel indicates the total sample size. The bottom panels indicate the sample size from each population. Numerical results are available in Supplementary Table 4.

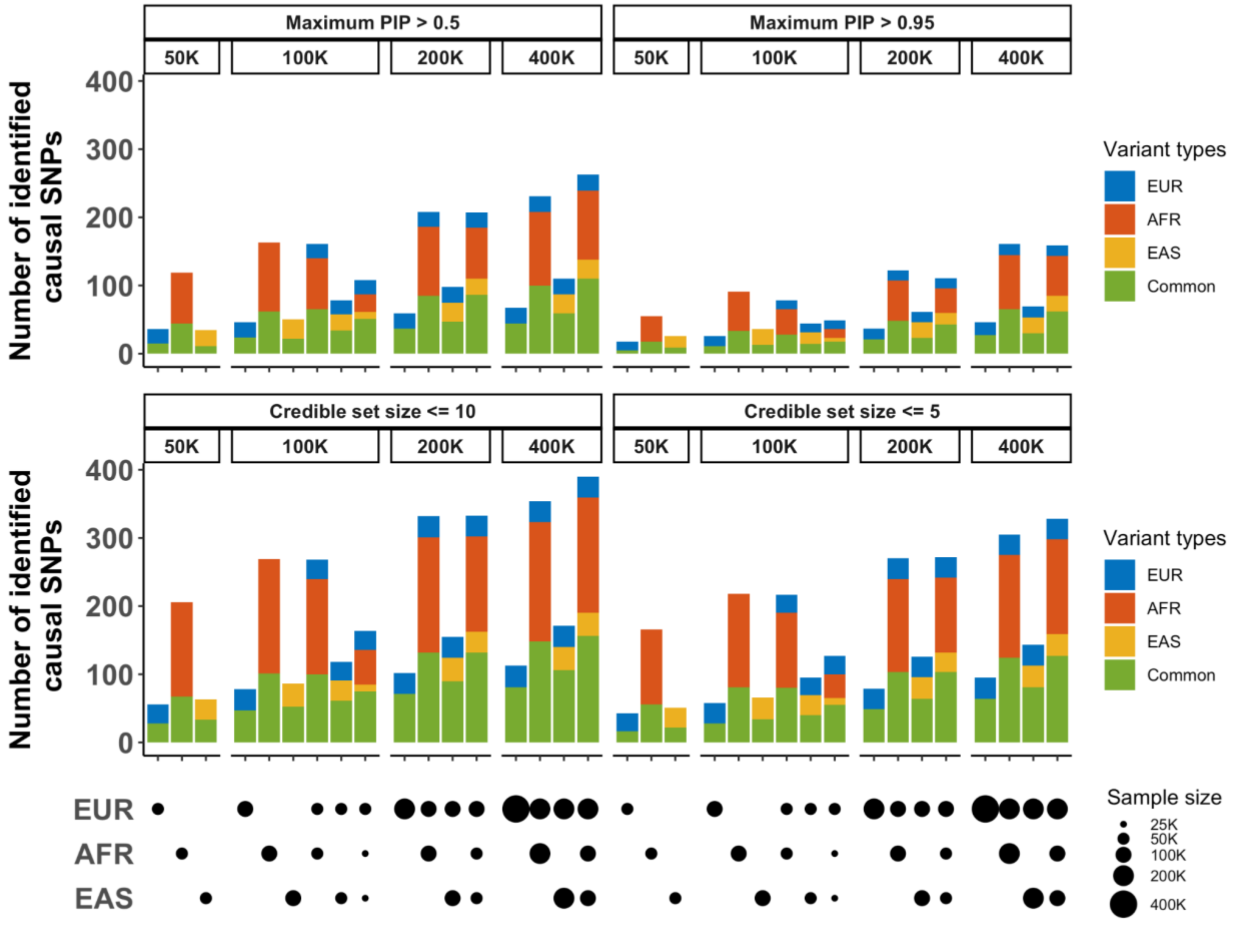


**Supplementary Figure 2: The number of confidently identified causal SNPs under the standard simulation setting.** The top label of each subpanel indicates the total sample size and the thresholds for selecting the credible sets. The bottom panels indicate the sample size from each population. Numerical results are available in Supplementary Table 4.

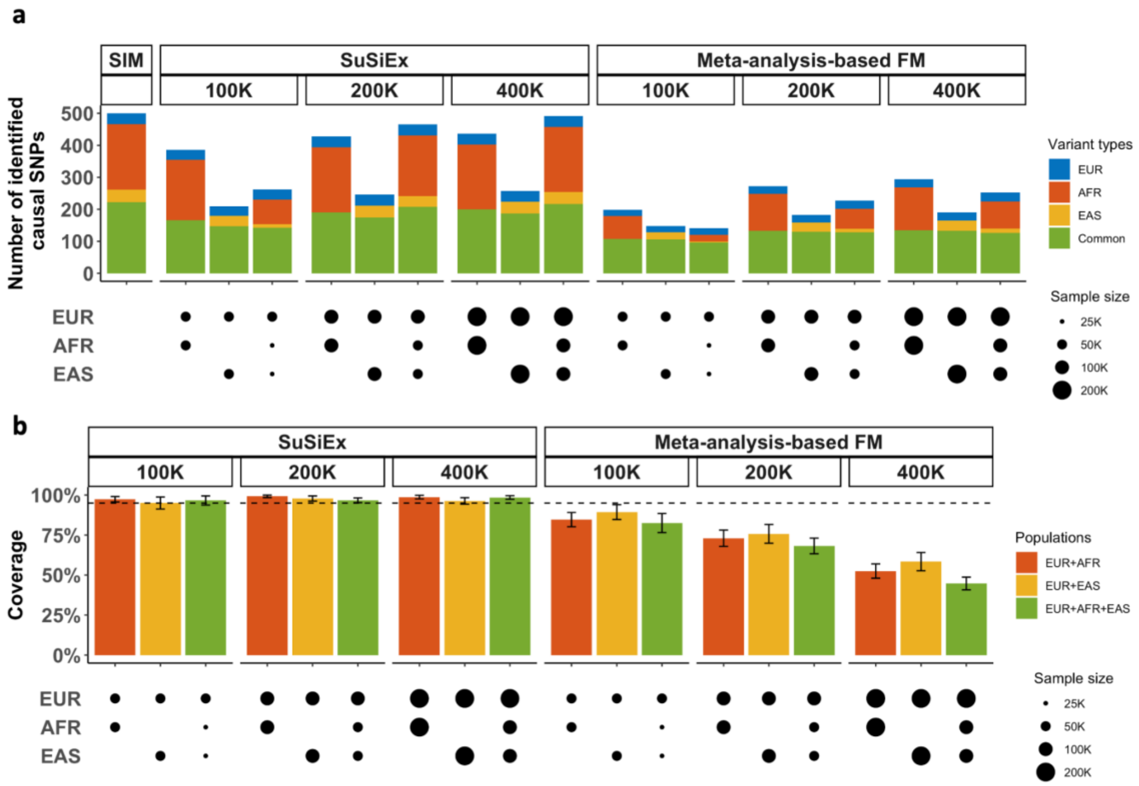


**Supplementary Figure 3: Comparison between SuSiEx and the meta-analysis-based fine-mapping method using the standard simulation setting. a**, The number of identified true causal variants (true causal variants covered by a credible set). **b**, The coverage of credible sets. The dashed line represents the 95% coverage. The error bar represents the 95% confidence interval. The top label of each subpanel indicates the total sample size and the fine-mapping method. The bottom panels indicate the sample size from each population. Numerical results are available in Supplementary Table 5.

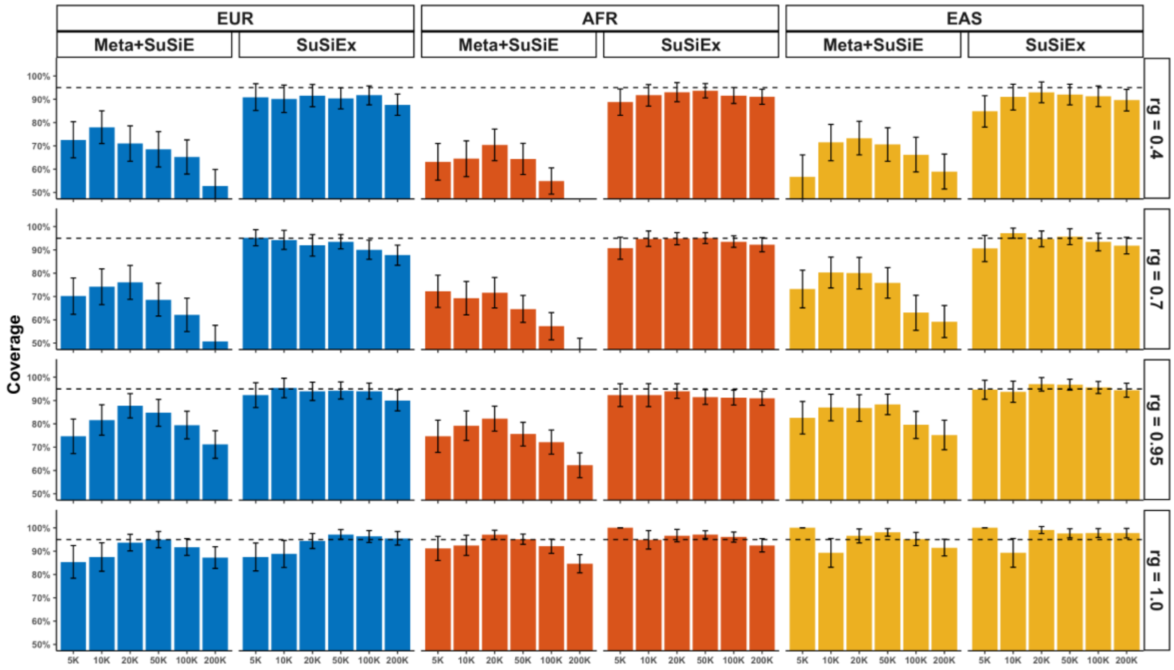


**Supplementary Figure 4: The coverage of Meta+SuSiE and SuSiEx when analyzing genetically close populations.** The top label of each subpanel indicates the continental population in which the analysis was performed and the fine-mapping method. The label on the right indicates the genetic correlation between the two subpopulations. The x-axis shows the discovery sample size of each subpopulation. The y-axis shows the coverage. The dashed line represents the 95% coverage. The error bar represents the 95% confidence interval. Numerical results are available in Supplementary Table 6.

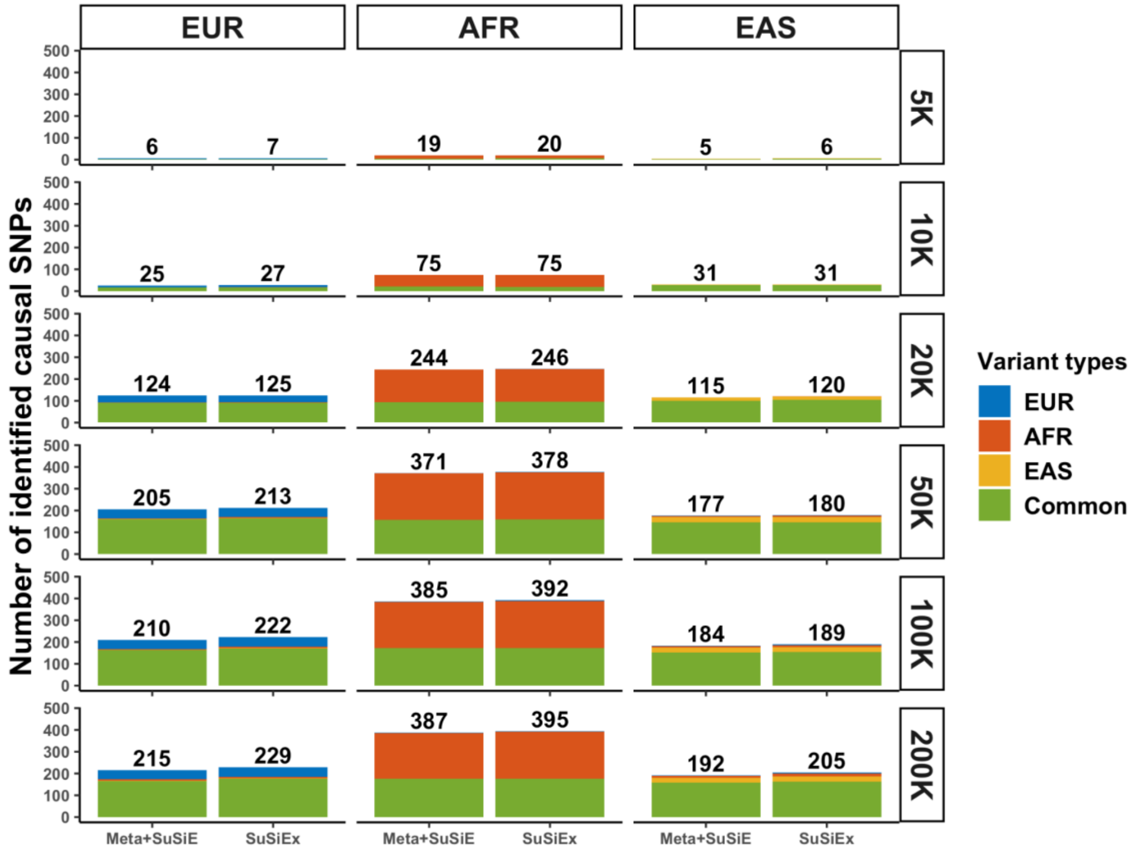


**Supplementary Figure 5: The number of causal variants identified by Meta+SuSiE and SuSiEx when analyzing genetically close populations with identical causal effect sizes.** The top label on each subpanel indicates the continental population in which the analysis was performed. The label on the right indicates the discovery sample size of each subpopulation. The x-axis shows the fine-mapping method. The y-axis shows the number of identified true causal variants. Numerical results are available in Supplementary Table 6.

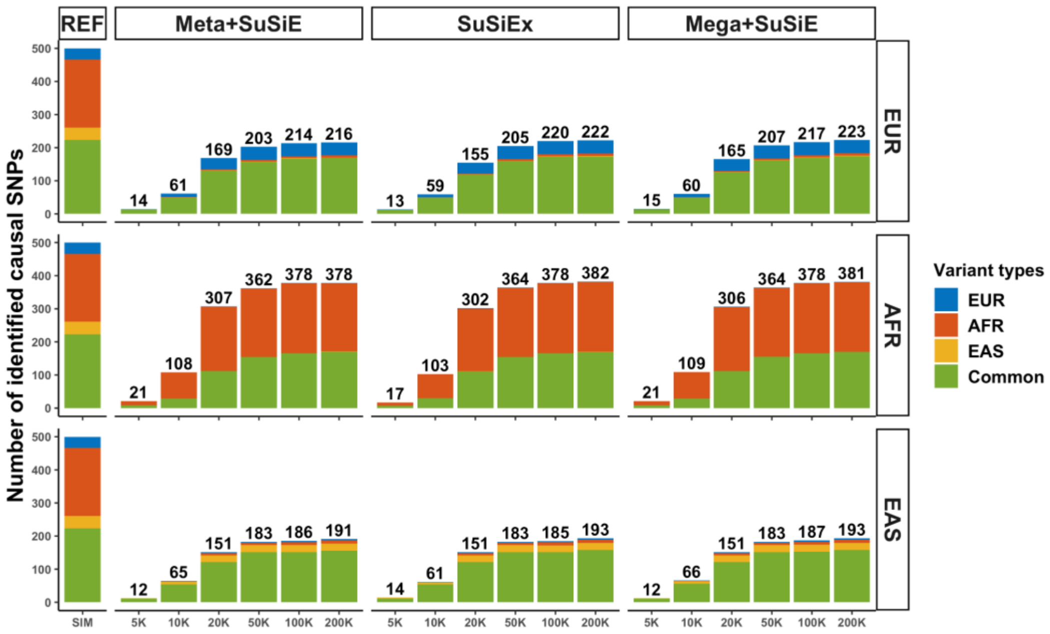


**Supplementary Figure 6: The number of causal variants identified by Meta+SuSiE, SuSiEx and Mega+SuSiE when analyzing two independent samples from the same population*.*** The top label on each subpanel indicates the fine-mapping method. The label on the right indicates the continental population in which analysis was performed. The x-axis shows the discovery sample size. The y-axis shows the number of identified true causal variants. The approach that performs GWAS on the merged dataset and applies SuSiE to the resulting GWAS is denoted as Mega+SuSiE. Numerical results are available in Supplementary Table 7.

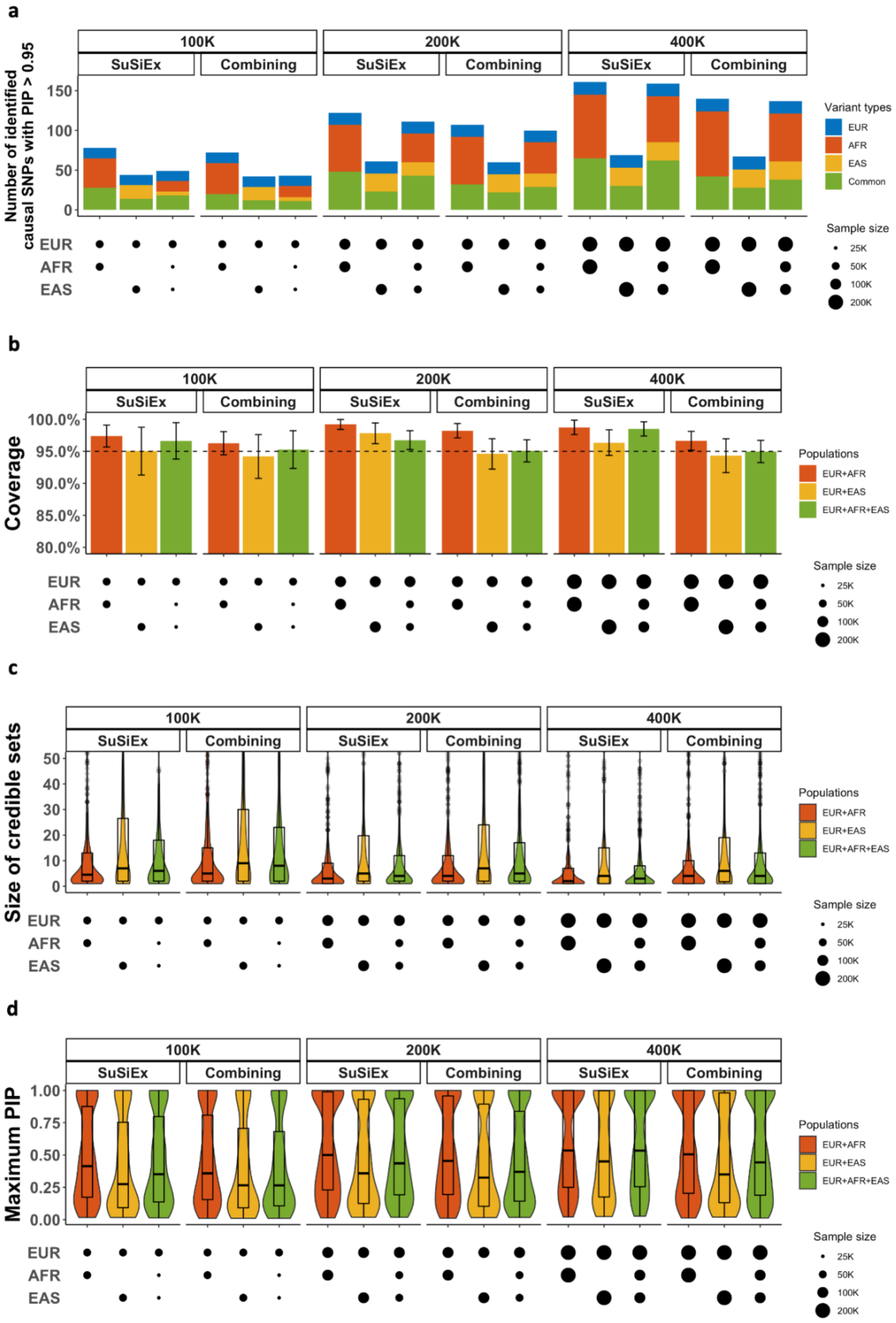
a
b
c
d
100K
200K
400K
SuSiEx
Combining
Number of identified causal SNPs with PIP > 0.95
Coverage
Size of credible sets
Maximum PIP
Variant types
EUR
AFR
EAS
Common
Populations
EUR+AFR
EUR+EAS
EUR+AFR+EAS
Sample size
25K
50K
100K
200K

**Supplementary Figure 7: Comparison between SuSiEx and the single-population combining method under the standard simulation setting. a**, The number of identified true causal variants with PIP >0.95 when integrating data from different populations with different sample sizes for fine-mapping. **b**, The coverage of credible sets. The dashed line represents the 95% coverage. The error bar represents the 95% confidence interval. **c**, Distribution of the size of credible sets. **d,** Distribution of the maximum PIP. The upper and lower bounds of the box indicate the 75th and 25th percentiles, respectively. The middle line in the box indicates the median. The top label of each subpanel indicates the total sample size and the fine-mapping method. The bottom panels indicate the sample size from each population. Numerical results are available in Supplementary Table 5.

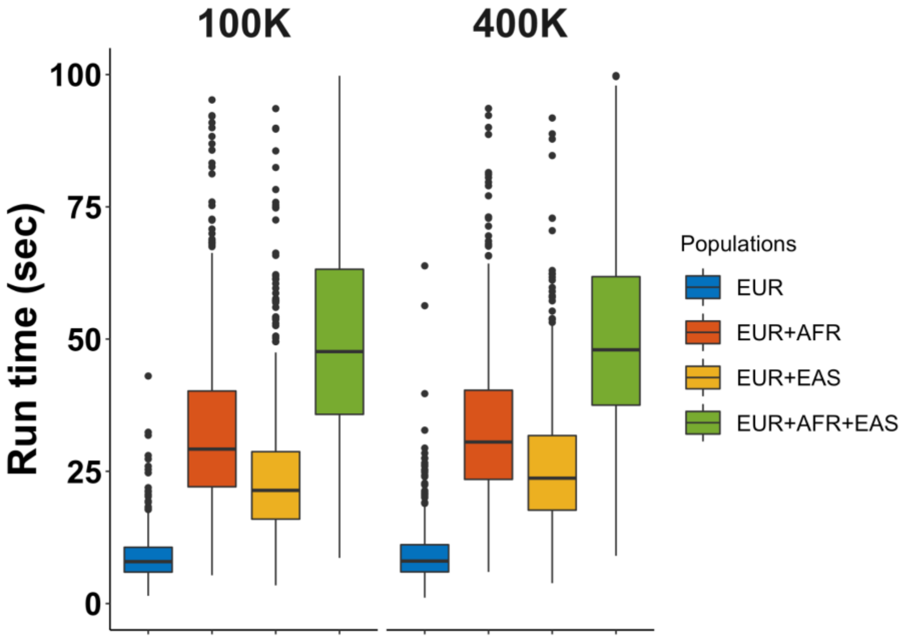


**Supplementary Figure 8: The runtime of SuSiEx under varying sample sizes and population combinations.** The y-axis shows the runtime of SuSiEx, measured in seconds, using a single CPU. Different colors indicate different population combinations. All analyses were conducted under the standard simulation settings. The total sample size is displayed at the top of each subpanel. The combinations of 'EUR+AFR' and 'EUR+EAS' were analyzed with a balanced sample size, while 'EUR+AFR+EAS' was analyzed with a sample size ratio of EUR:AFR:EAS = 2:1:1. Numerical results are available in Supplementary Table 8.

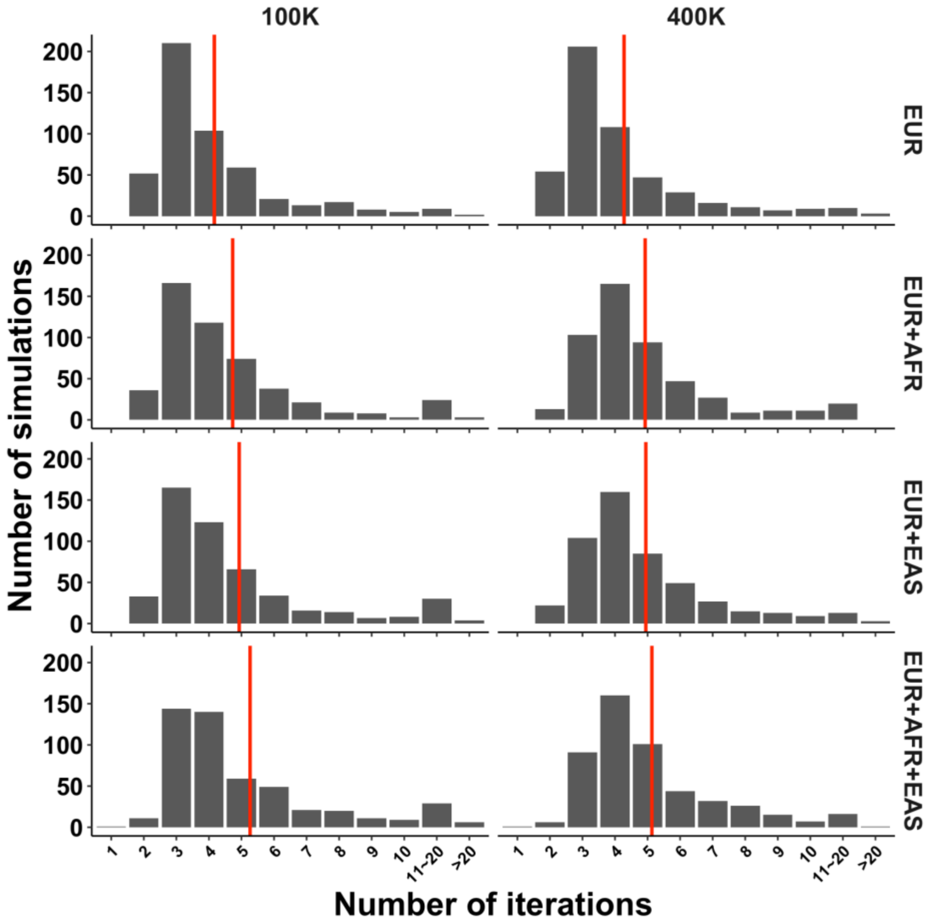


**Supplementary Figure 9: The number of iterations before SuSiEx converged under varying sample sizes and population combinations.** The histograms show the distributions of the number of iterations before SuSiEx converged. The red vertical line represents the average number of iterations. All analyses were conducted under the standard simulation settings. The total sample size is displayed at the top of each subpanel. The combinations of 'EUR+AFR' and 'EUR+EAS' were analyzed with a balanced sample size, while 'EUR+AFR+EAS' was analyzed with a sample size ratio of EUR:AFR:EAS = 2:1:1. Numerical results are available in Supplementary Table 8.

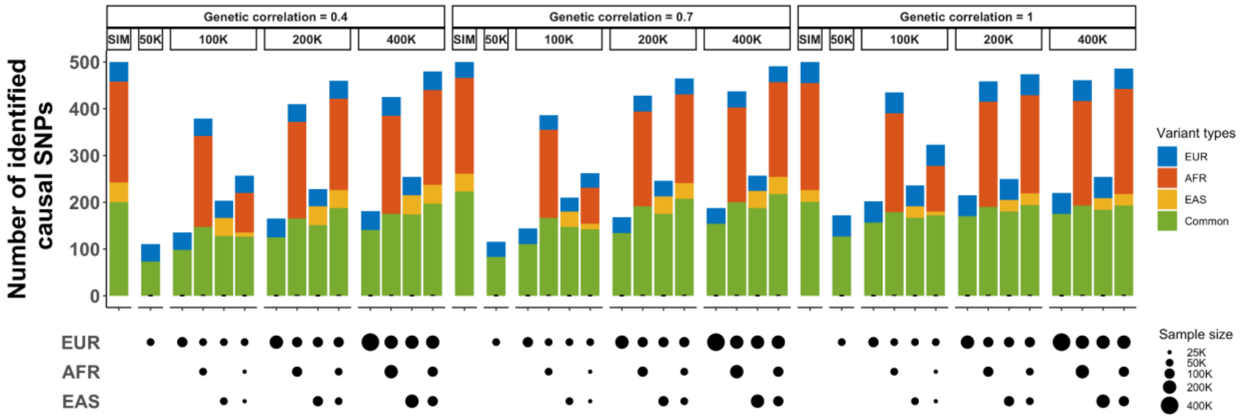


**Supplementary Figure 10: The number of causal SNPs identified by SuSiEx under varying genetic correlations (*$r_g$*).** The top label of each subpanel indicates the total sample size and the genetic correlation. The bottom panels indicate the sample size from each population. Numerical results are available in Supplementary Table 11.

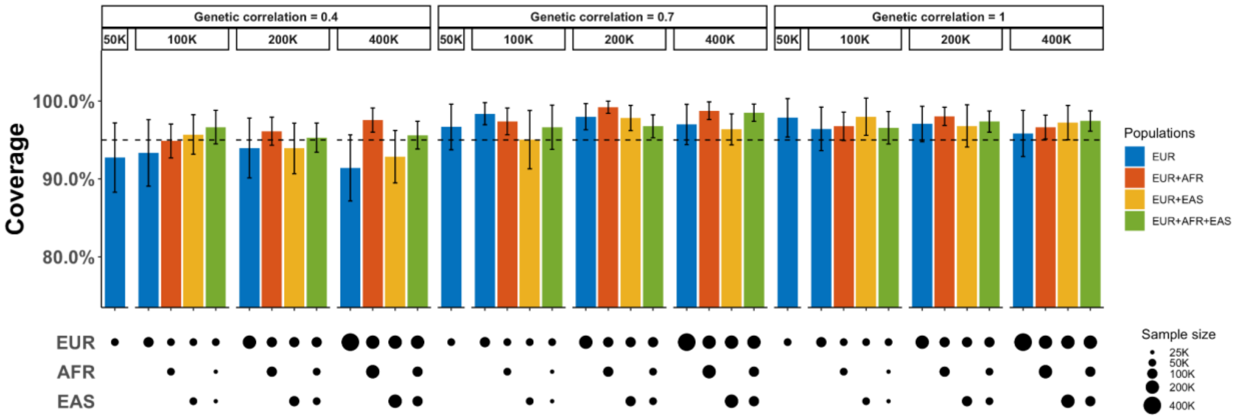


**Supplementary Figure 11: The coverage of SuSiEx under varying genetic correlations ($r_g$).** The dashed line represents the 95% coverage. The error bar represents the 95% confidence interval. The top label of each subpanel indicates the total sample size and the genetic correlation. The bottom panels indicate the sample size from each population. Numerical results are available in Supplementary Table 11.

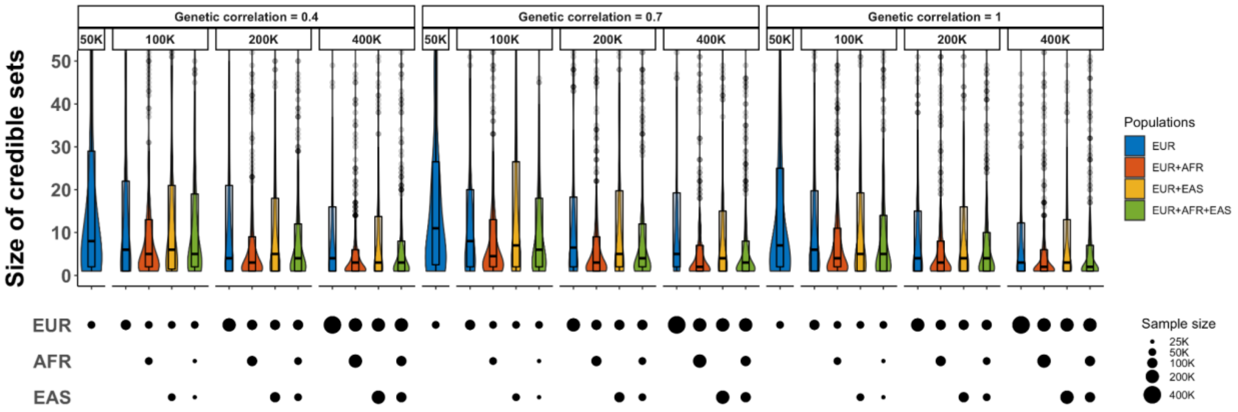


**Supplementary Figure 12: The size of credible sets identified by SuSiEx under varying genetic correlations ($r_g$).** The upper and lower bounds of the box indicate the 75th and 25th percentiles, respectively. The middle line in the box indicates the median. The top label of each subpanel indicates the total sample size and the genetic correlation. The bottom panels indicate the sample size from each population. Numerical results are available in Supplementary Table 11.

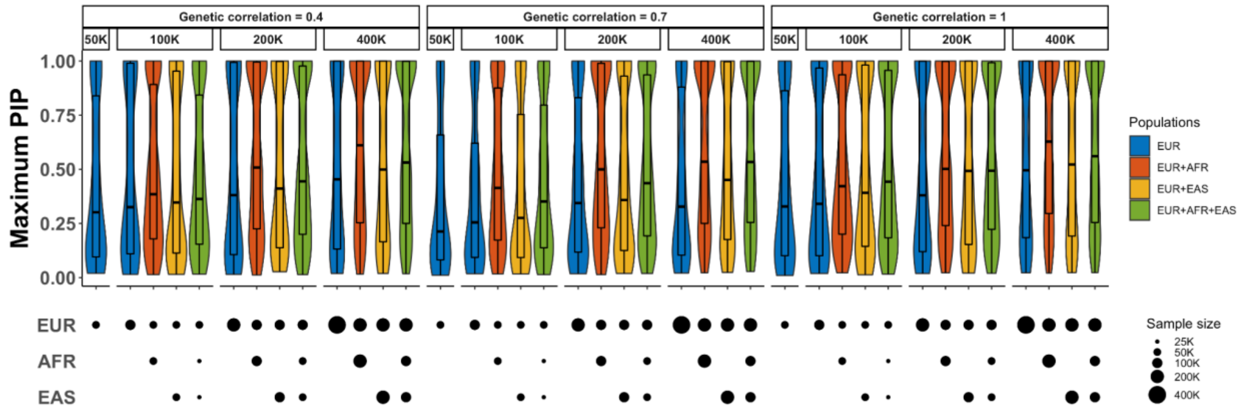


**Supplementary Figure 13: The maximum PIP estimated by SuSiEx under varying genetic correlations ($r_g$).** The upper and lower bounds of the box indicate the 75th and 25th percentiles, respectively. The middle line in the box indicates the median. The top label of each subpanel indicates the total sample size and the genetic correlation. The bottom panels indicate the sample size from each population. Numerical results are available in Supplementary Table 11.

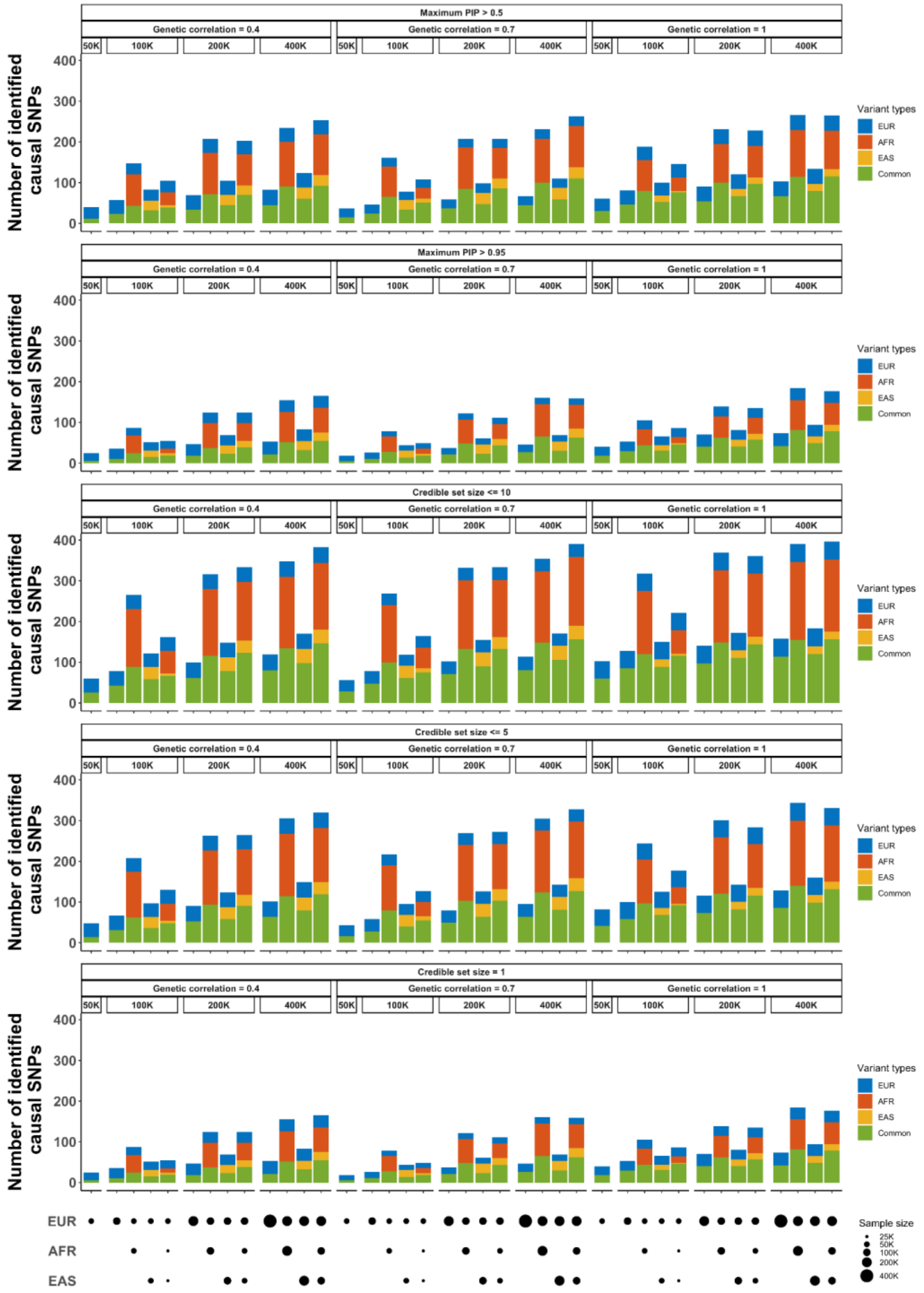


**Supplementary Figure 14: The number of confidently identified causal SNPs under varying genetic correlations ($r_g$).** The top label of each subpanel indicates the total sample size and the confidence level for the credible sets. The bottom panels indicate the sample size from each population. Numerical results are available in Supplementary Table 11.

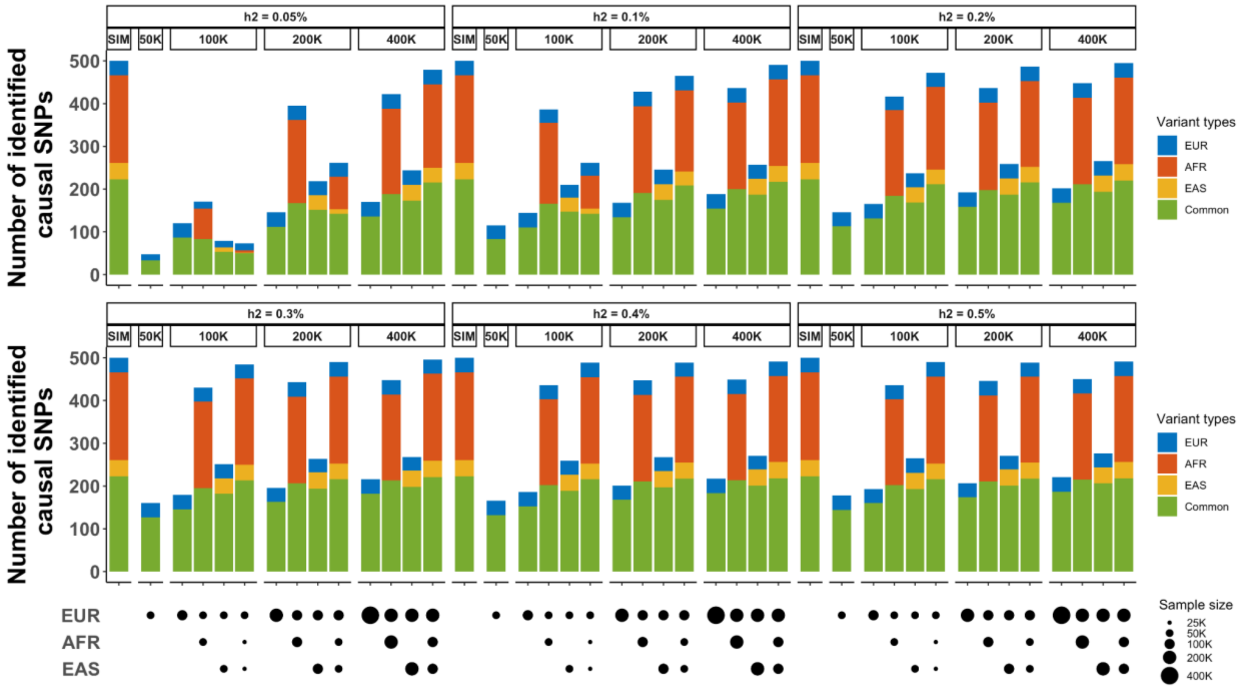


**Supplementary Figure 15: The number of causal SNPs identified by SuSiEx under varying local heritability (*$h^2$*).** The top label of each subpanel indicates the total sample size and the local heritability. The bottom panels indicate the sample size from each population. Numerical results are available in Supplementary Table 12.

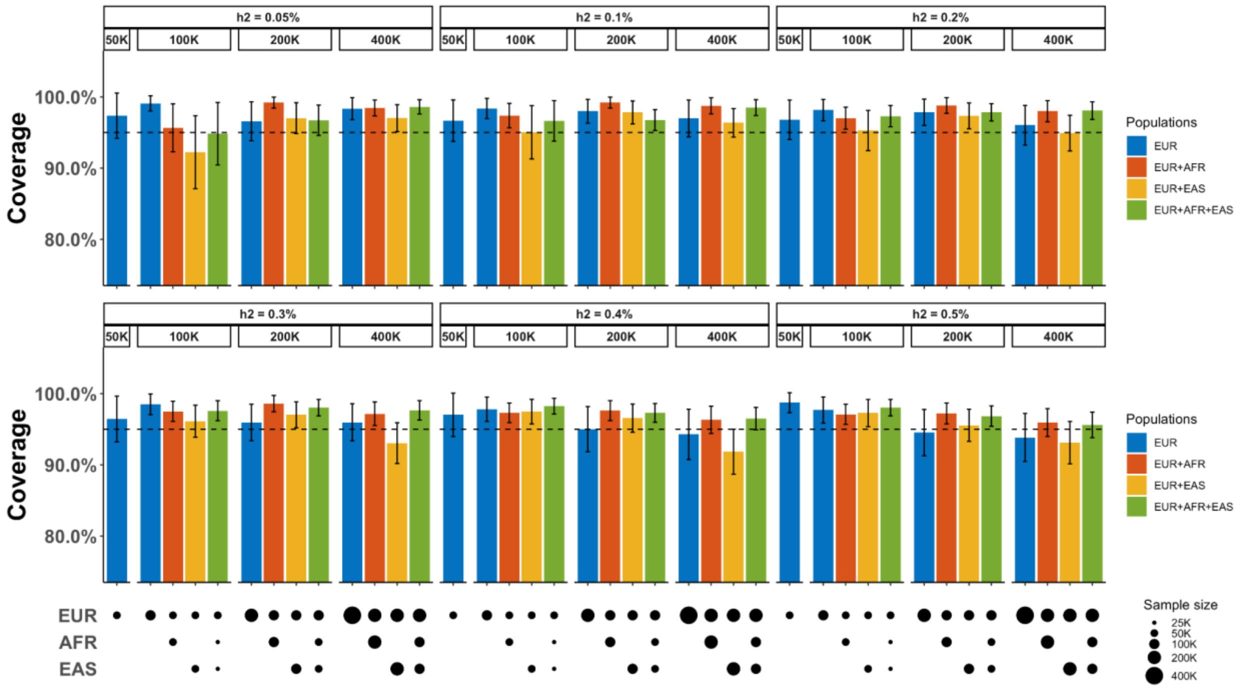


**Supplementary Figure 16: The coverage of SuSiEx under varying local heritability (*$h^2$*).** The dashed line represents the 95% coverage. The error bar represents the 95% confidence interval. The top label of each subpanel indicates the total sample size and the local heritability. The bottom panels indicate the sample size from each population. Numerical results are available in Supplementary Table 12.

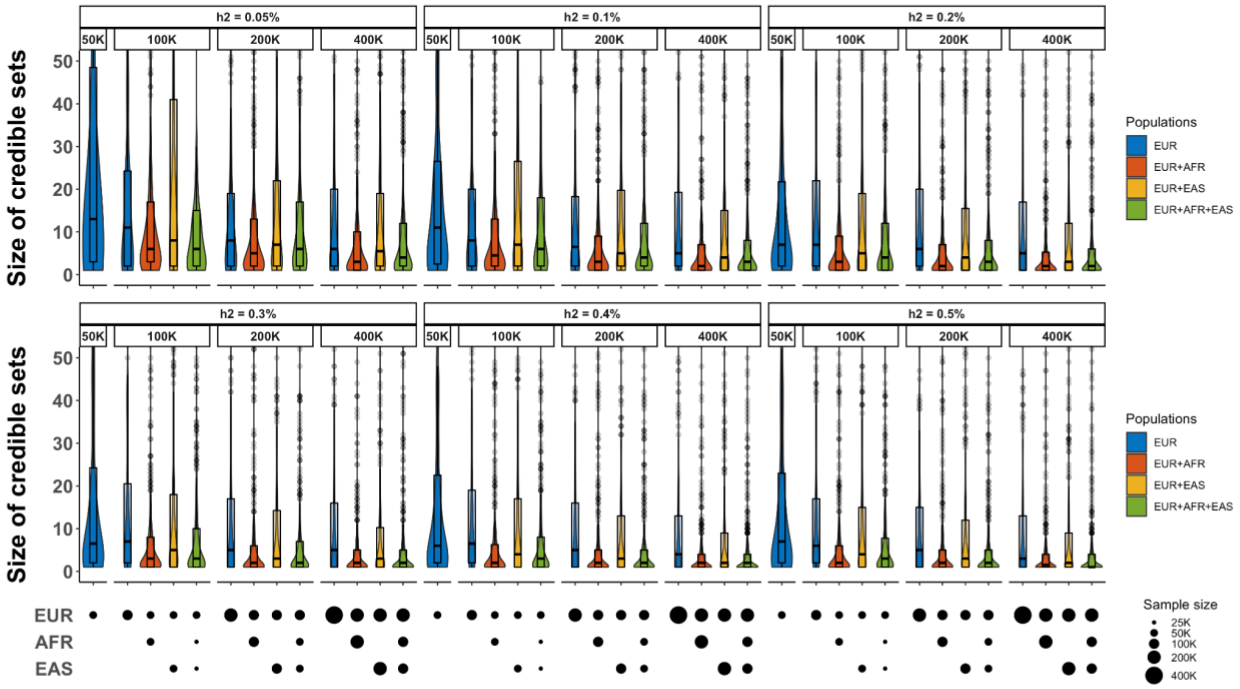


**Supplementary Figure 17: The size of credible sets identified by SuSiEx under varying local heritability ($h^2$).** The upper and lower bounds of the box indicate the 75th and 25th percentiles, respectively. The middle line in the box indicates the median. The top label of each subpanel indicates the total sample size and the local heritability. The bottom panels indicate the sample size from each population. Numerical results are available in Supplementary Table 12.

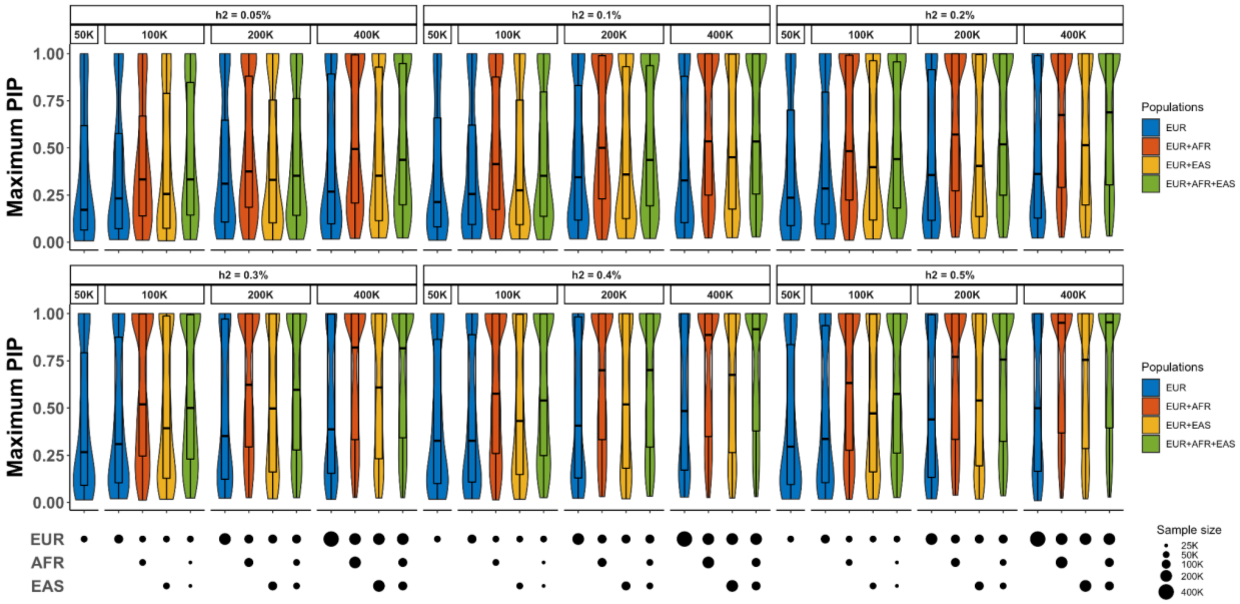


**Supplementary Figure 18: The maximum PIP estimated by SuSiEx under varying local heritability (*$h^2$*).** The upper and lower bounds of the box indicate the 75th and 25th percentiles, respectively. The middle line in the box indicates the median. The top label of each subpanel indicates the total sample size and the local heritability. The bottom panels indicate the sample size from each population. Numerical results are available in Supplementary Table 12.

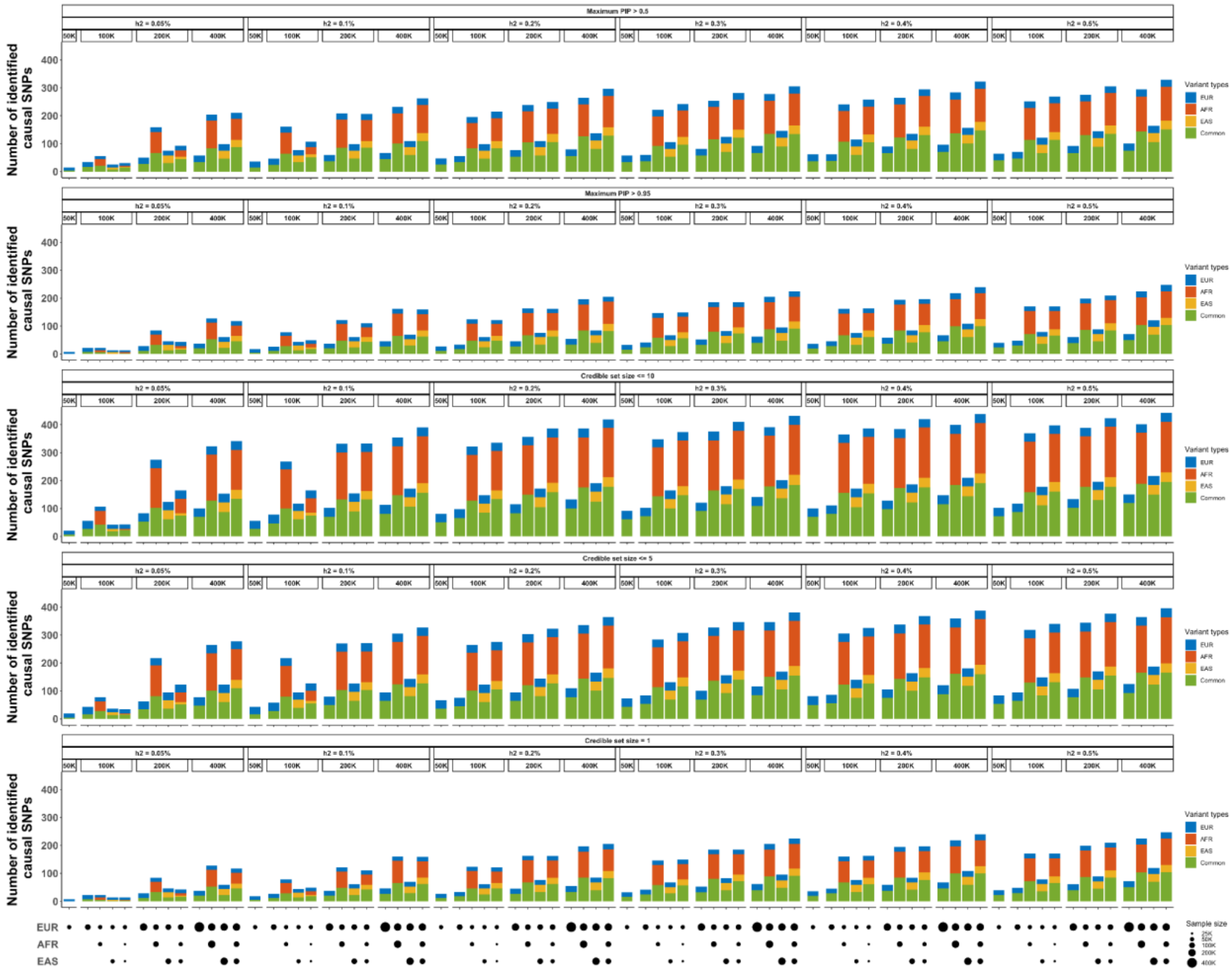

**Supplementary Figure 19: The number of confidently identified causal SNPs under varying local heritability ($h^2$).** The top label of each subpanel indicates the total sample size and the confidence level for the credible sets. The bottom panels indicate the sample size from each population. Numerical results are available in Supplementary Table 12.

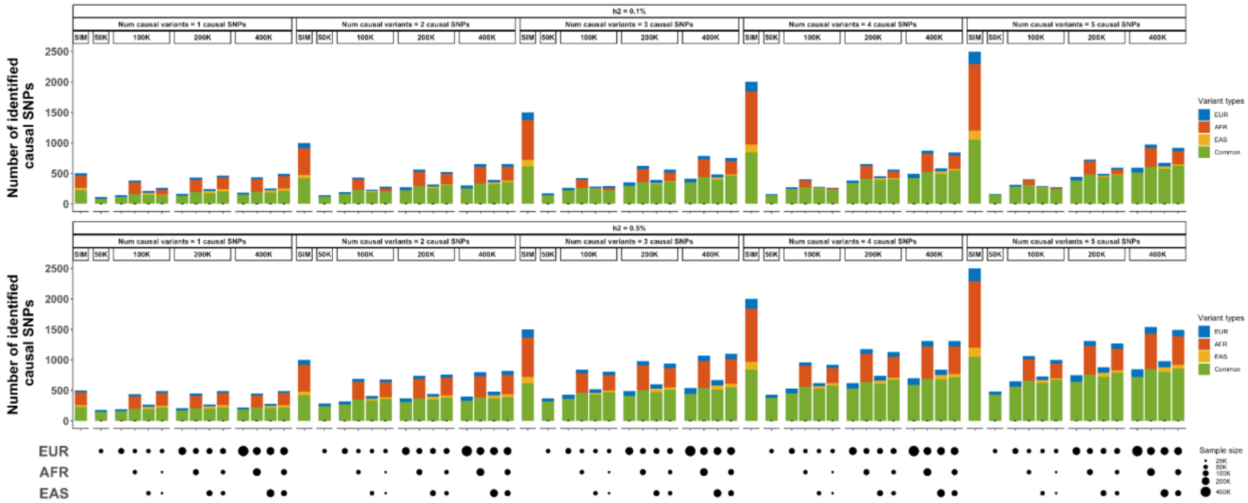


**Supplementary Figure 20: The number of causal SNPs identified by SuSiEx under varying numbers of causal SNPs per locus ($n_{csl}$).** The top label of each subpanel indicates the total sample size, the local heritability and the number of causal SNPs. The bottom panels indicate the sample size from each population. Numerical results are available in Supplementary Table 13.

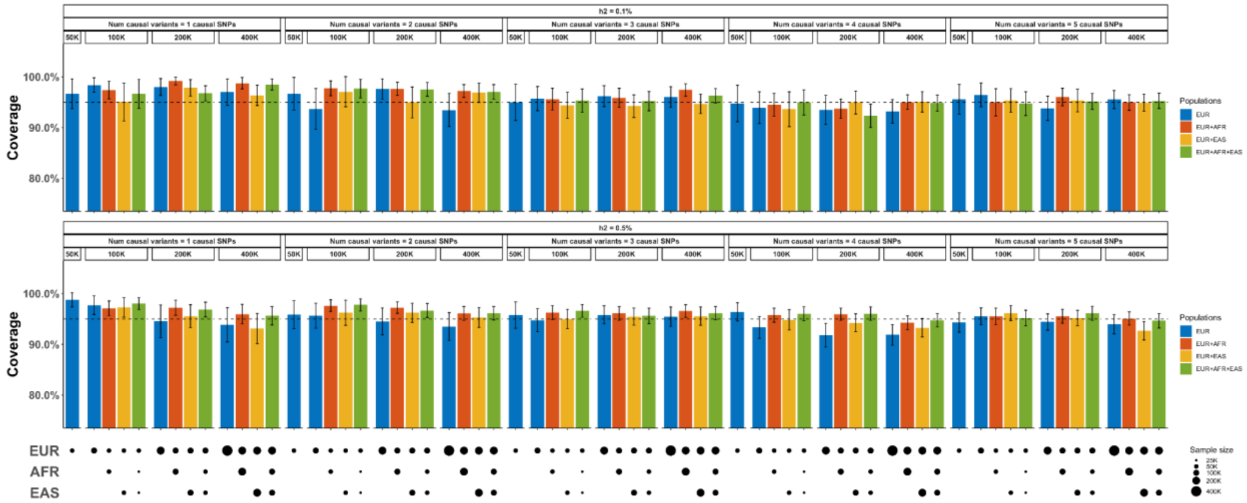


**Supplementary Figure 21: The coverage of SuSiEx under varying numbers of causal SNPs per locus ($n_{csl}$).** The dashed line represents the 95% coverage. The error bar represents the 95% confidence interval. The top label of each subpanel indicates the total sample size, the local heritability and the number of causal SNPs. The bottom panels indicate the sample size from each population. Numerical results are available in Supplementary Table 13.

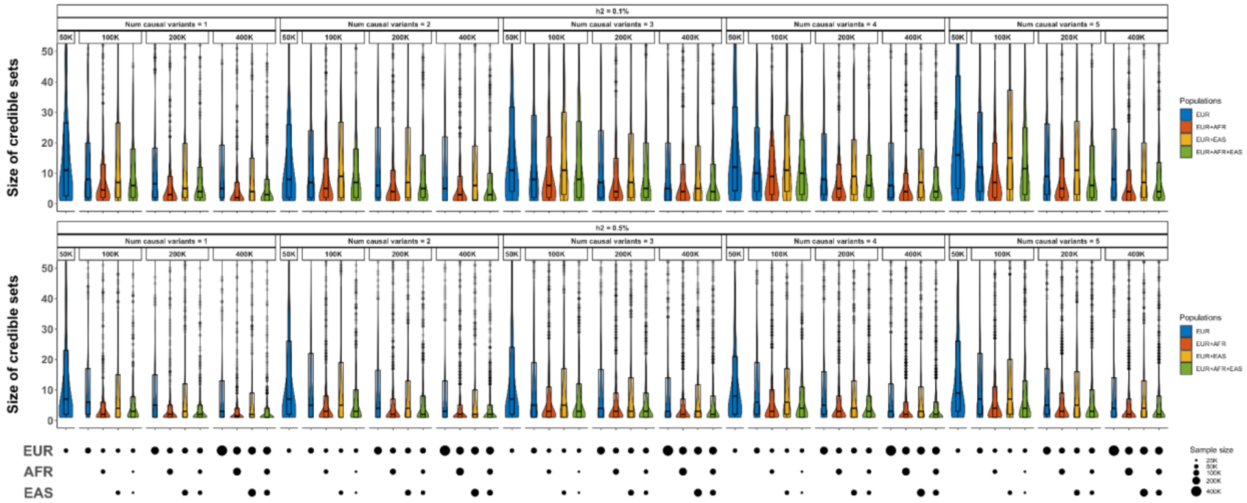


**Supplementary Figure 22: The size of credible sets identified by SuSiEx under varying numbers of causal SNPs per locus ($n_{csl}$).** The upper and lower bounds of the box indicate the 75th and 25th percentiles, respectively. The middle line in the box indicates the median. The top label of each subpanel indicates the total sample size, the local heritability and the number of causal SNPs. The bottom panels indicate the sample size from each population. Numerical results are available in Supplementary Table 13.

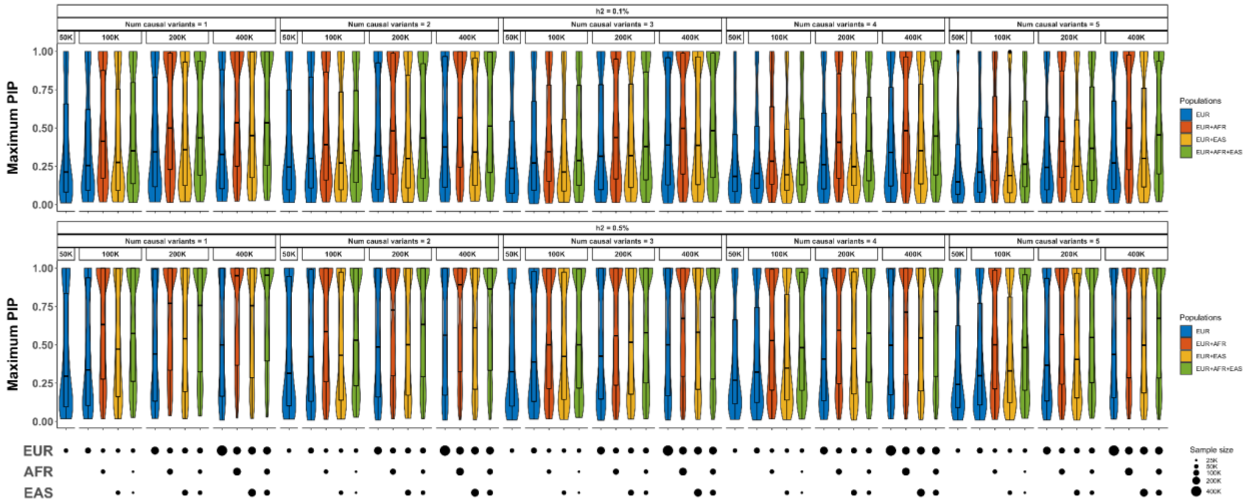


**Supplementary Figure 23: The maximum PIP estimated by SuSiEx under varying numbers of causal SNPs per locus ($n_{csl}$).** The upper and lower bounds of the box indicate the 75th and 25th percentiles, respectively. The middle line in the box indicates the median. The top label of each subpanel indicates the total sample size, the local heritability and the number of causal SNPs. The bottom panels indicate the sample size from each population. Numerical results are available in Supplementary Table 13.

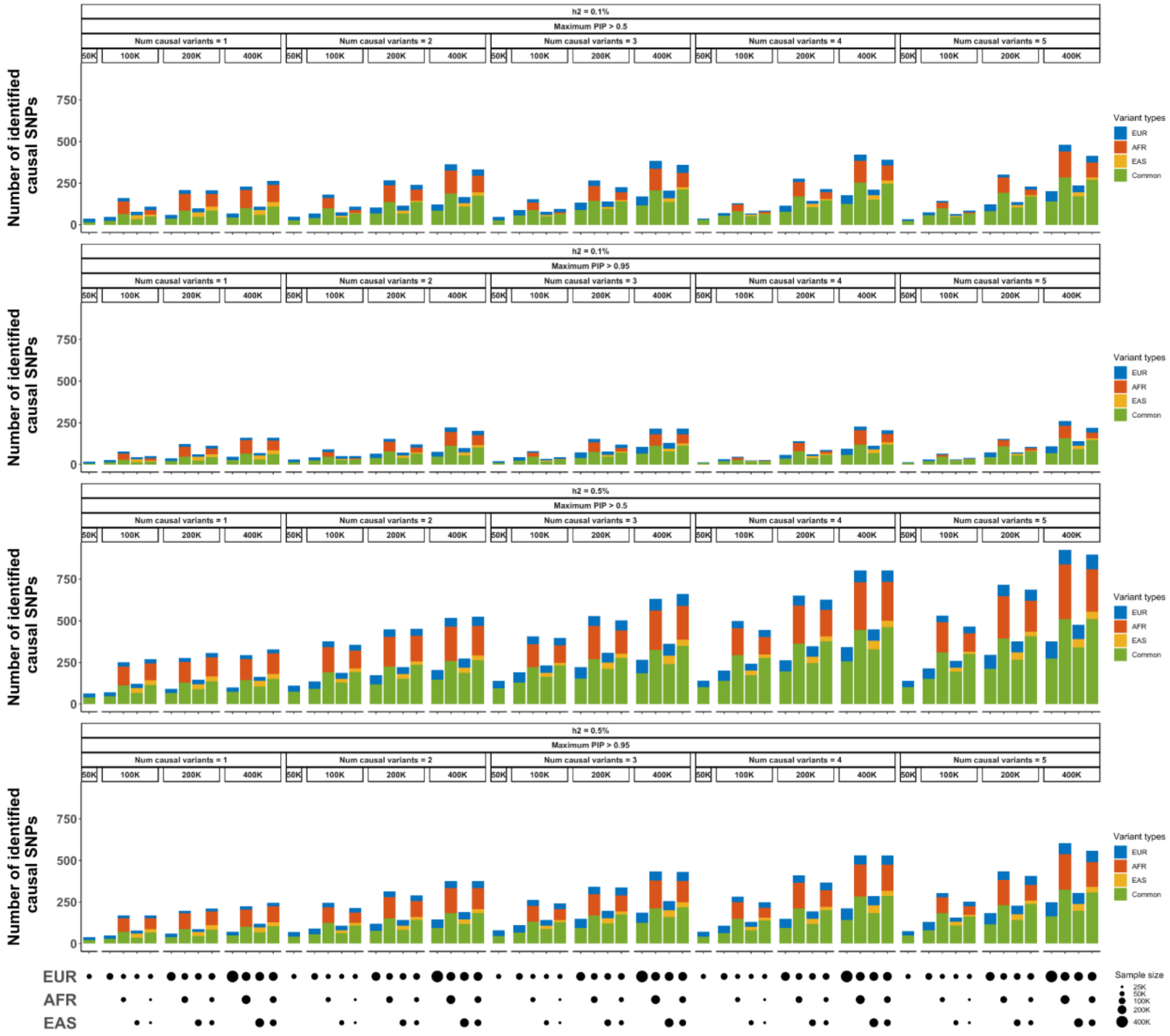


**Supplementary Figure 24: The number of confidently identified causal SNPs under varying numbers of causal SNPs per locus ($n_{csl}$).** The top label of each subpanel indicates the total sample size and the confidence level for the credible sets. The bottom panels indicate the sample size from each population. Numerical results are available in Supplementary Table 13.

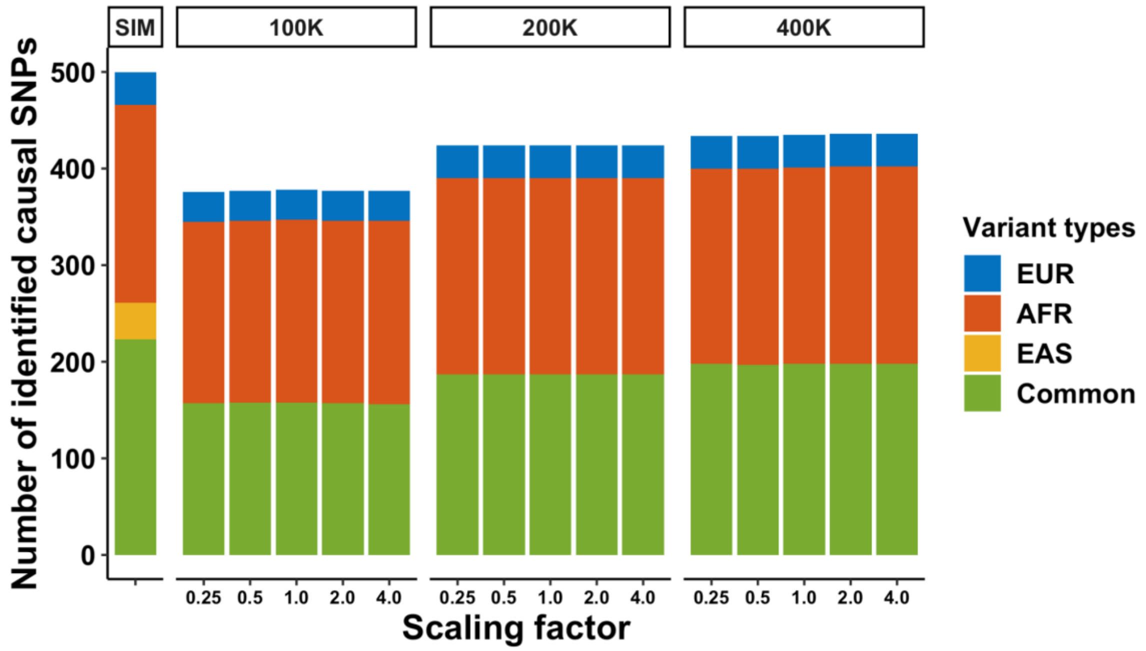


**Supplementary Figure 25: The impact of different $\tau_s^2$ values on the performance of SuSiEx.** The top label on each subpanel indicates the discovery sample size of each population. The x-axis shows the factor by which the $\tau_s^2$ parameter is scaled. The y-axis shows the number of identified true causal variants. Simulations were conducted under the standard parameter setting with a balanced EUR and AFR sample size. The total sample size is shown above each panel. Numerical results are available in Supplementary Table 14.

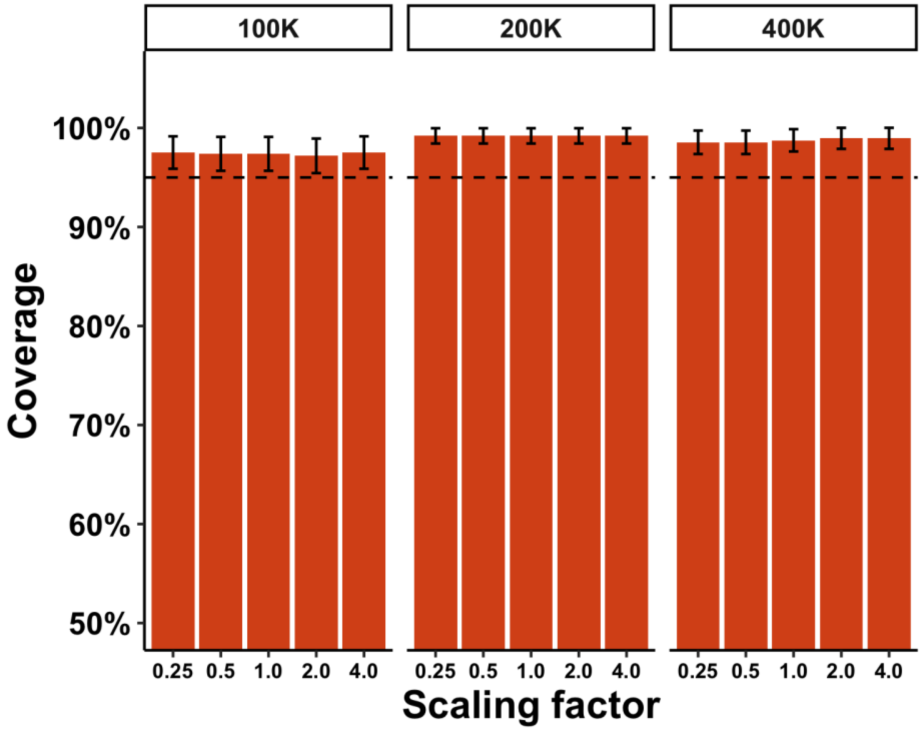


**Supplementary Figure 26: The impact of different $\tau_s^2$ values on the calibration of SuSiEx.** The top label on each subpanel indicates the discovery sample size of each population. The x-axis shows the factor by which the $\tau_s^2$ parameter is scaled. The y-axis shows the coverage. The dashed line represents 95% coverage. The error bar represents the 95% confidence interval. Simulations were conducted under the standard parameter setting with a balanced EUR and AFR sample size. The total sample size is shown above each panel. Numerical results are available in Supplementary Table 14.

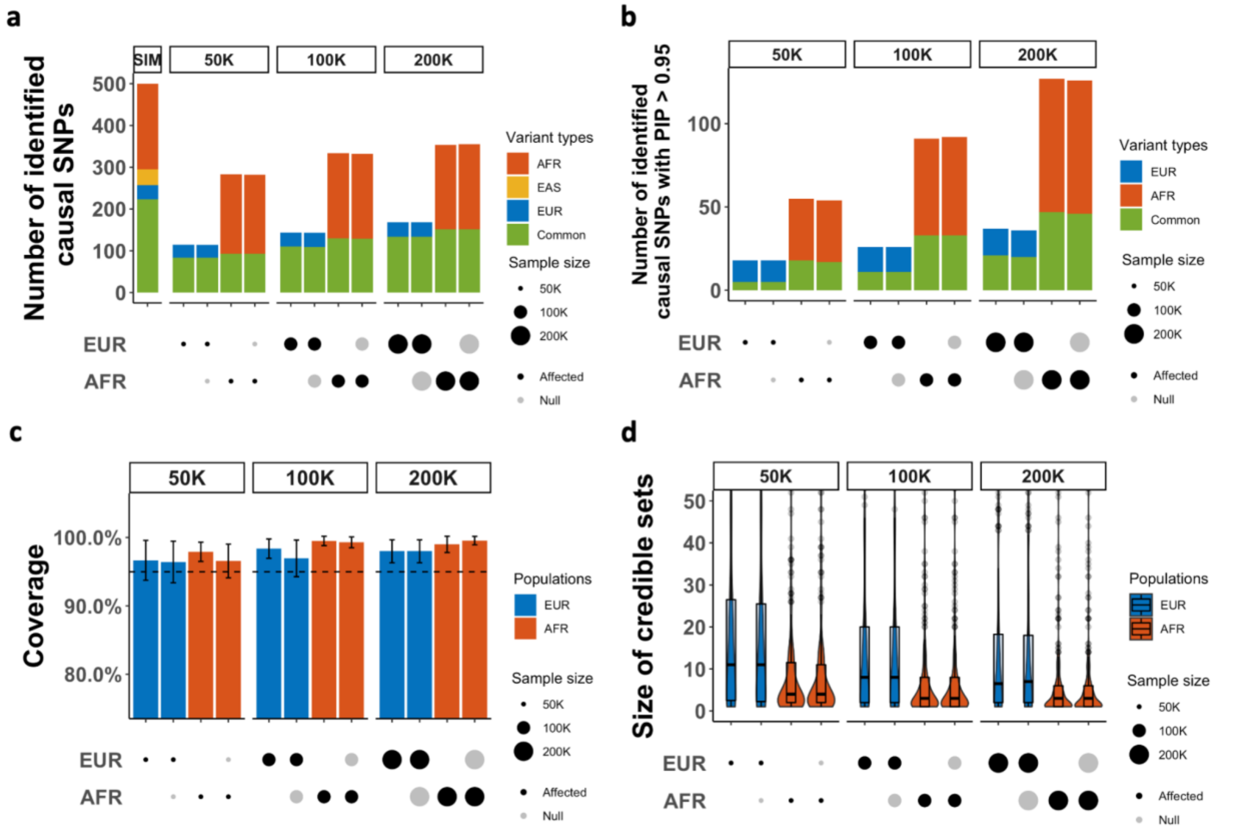


**Supplementary Figure 27: Performance of SuSiEx in the presence of population-specific causal SNPs. a**, The number of identified true causal variants when integrating data from different populations with different sample sizes for fine-mapping. **b**, The number of identified causal variants with PIP >0.95 when integrating data from different populations with different sample sizes for fine-mapping. **c**, The coverage of credible sets. The dashed line represents the 95% coverage. The error bar represents the 95% confidence interval. **d**, Distribution of the size of credible sets. The upper and lower bounds of the box indicate the 75th and 25th percentiles, respectively. The middle line in the box indicates the median. The top label of each subpanel indicates the total sample size in which SNP effects were non-null, and the bottom panels indicate the sample size from each population. Black circles indicate sample size with non-null SNP effects; gray circles indicate the sample size with null SNP effects. Simulated data were generated under the standard simulation setting. Numerical results are available in Supplementary Table 15.

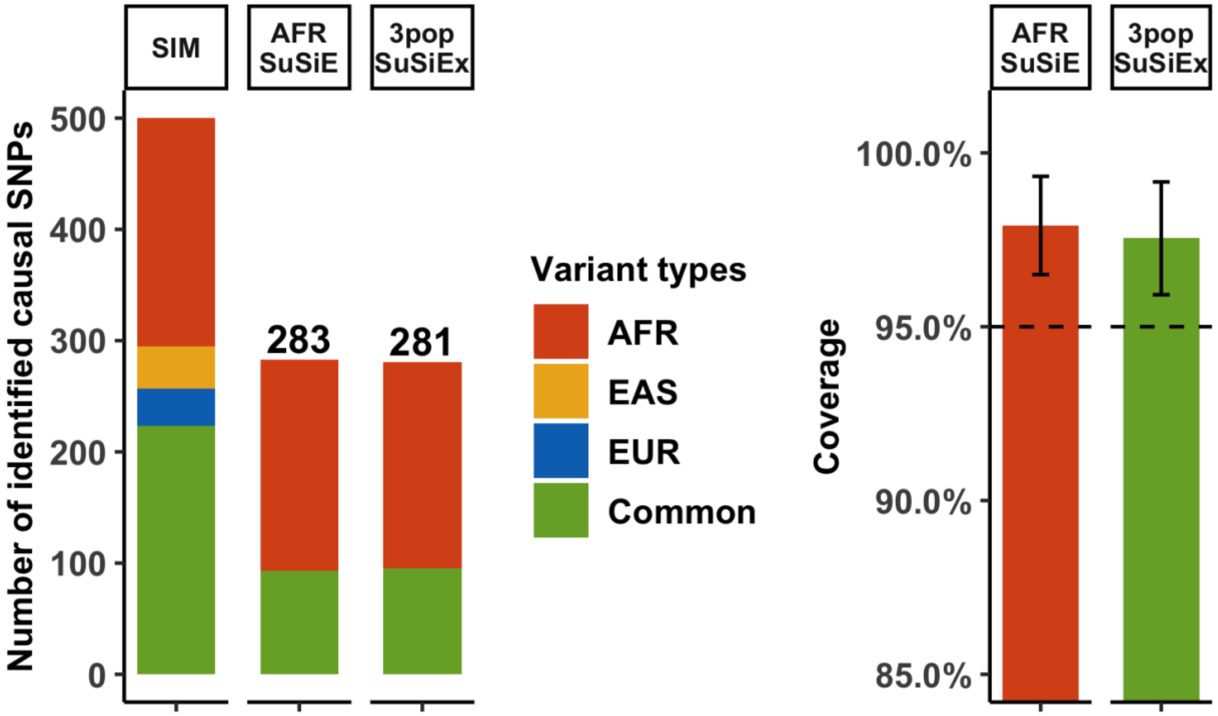


**Supplementary Figure 28: Performance of SuSiEx in the presence of African-specific causal variants.** Left: The number of identified true causal variants when using single-population SuSiE in the AFR population vs. using cross-population SuSiEx that integrates data from the three populations. Right: The coverage of credible sets. The dashed line represents 95% coverage. The error bar represents the 95% confidence interval. Numerical results are available in Supplementary Table 15.

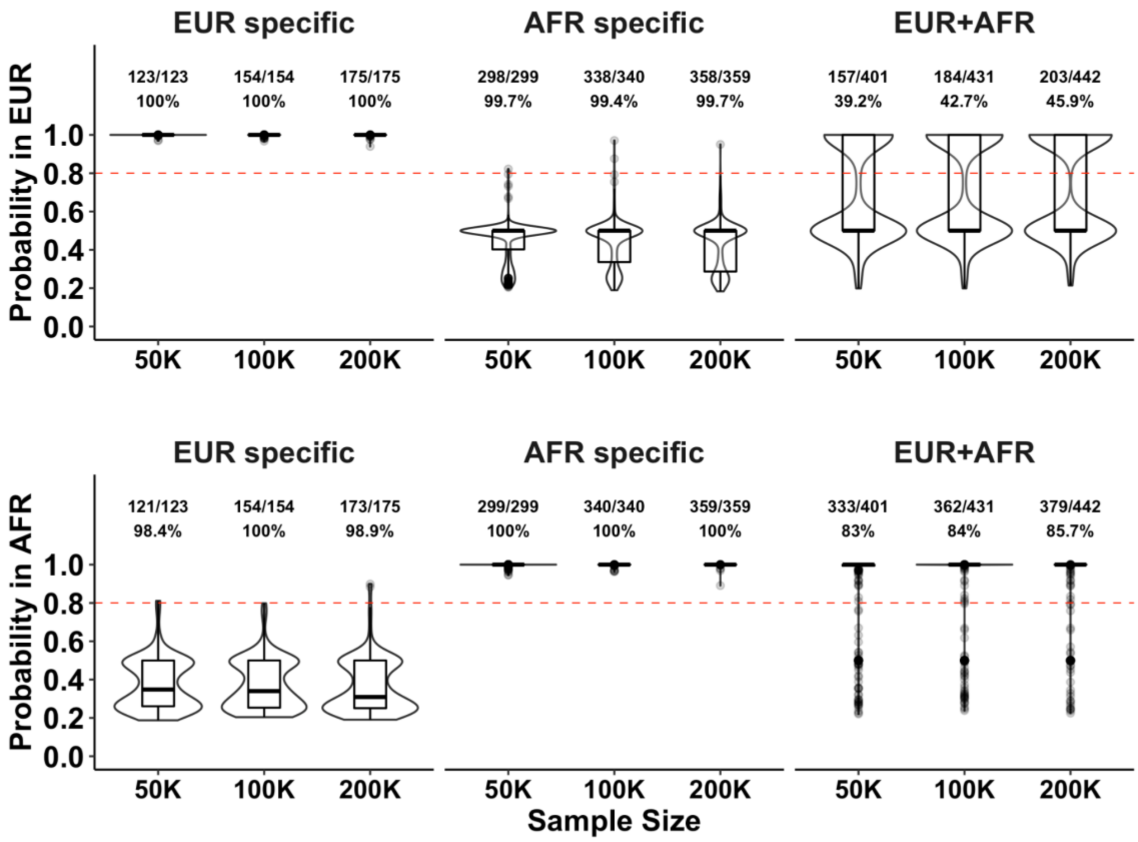


**Supplementary Figure 29: The population-specific causal probability under the standard simulation setting.** Analyses were conducted with a balanced EUR and AFR sample size. The x-axis shows the discovery sample size. The y-axis shows the population-specific causal probability of the identified credible set. The red dashed lines represent the probability of 0.8, which is used as a threshold to infer whether an identified credible set is causal in a population. The label at the top of each panel denotes the ground-truth causal configuration: “EUR specific” indicates that the variant is causal in the EUR population but not in the AFR population; “AFR specific” indicates that the variant is causal in the AFR population but not in the EUR population; “EUR+AFR” indicates that the variant is causal in both EUR and AFR populations. The fractional number on each panel represents “the number of credible sets with correct inference” among “the total number of credible sets identified by SuSiEx”. The percentage represents the accuracy of the inference. Numerical results are available in Supplementary Table 16.

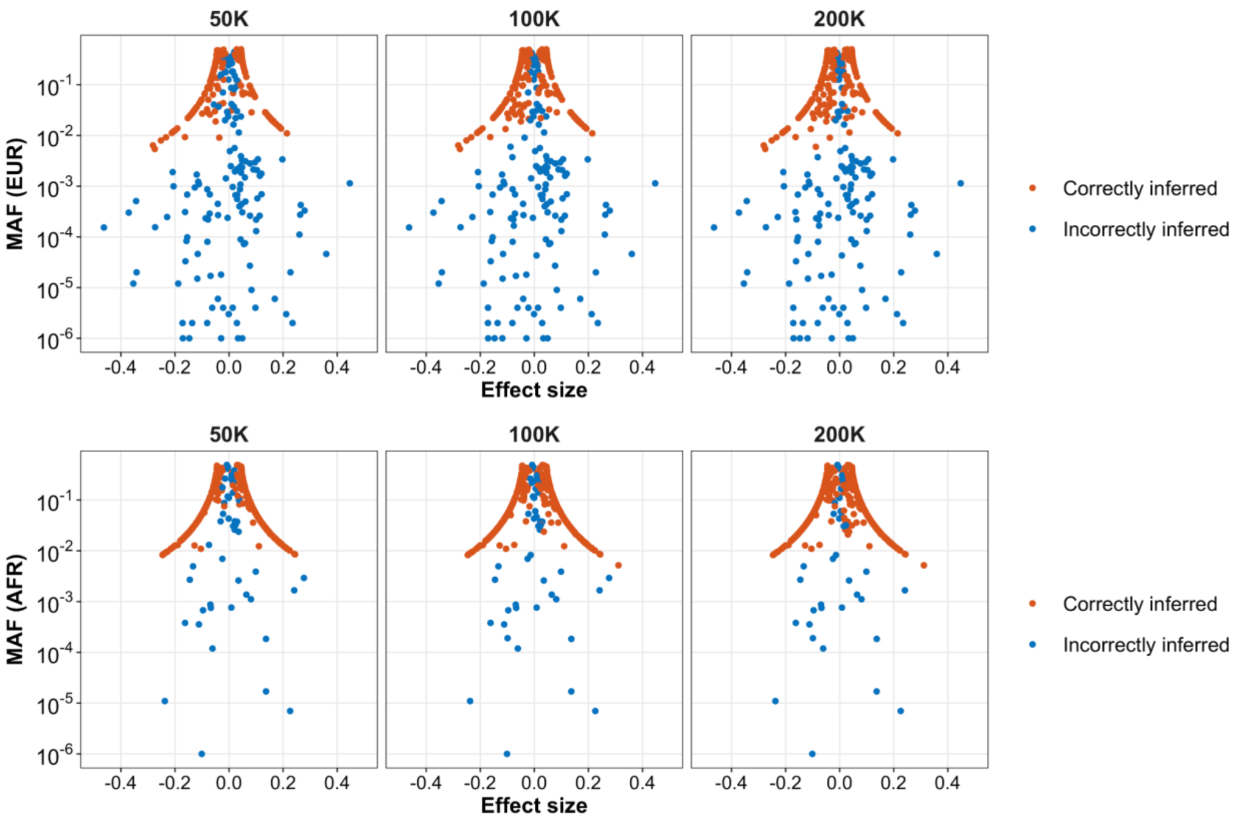


**Supplementary Figure 30: The impact of allele frequency and causal effect size on the classification of population-specific causal variants.** This plot corresponds to the simulation under the “EUR+AFR” scenario in Supplementary Figure 29. The x-axis shows the simulated effect sizes of the causal variants. The y-axis shows the minor allele frequencies (MAF) of the causal variants in a specific population. The label above each panel indicates the discovery sample size. Red dots represent variants that are correctly inferred to be causal in the population. Blue dots represent variants that are incorrectly inferred to be not causal in the population. Incorrectly inferred variants tend to have low MAF or small effect sizes. Numerical results are available in Supplementary Table 16.

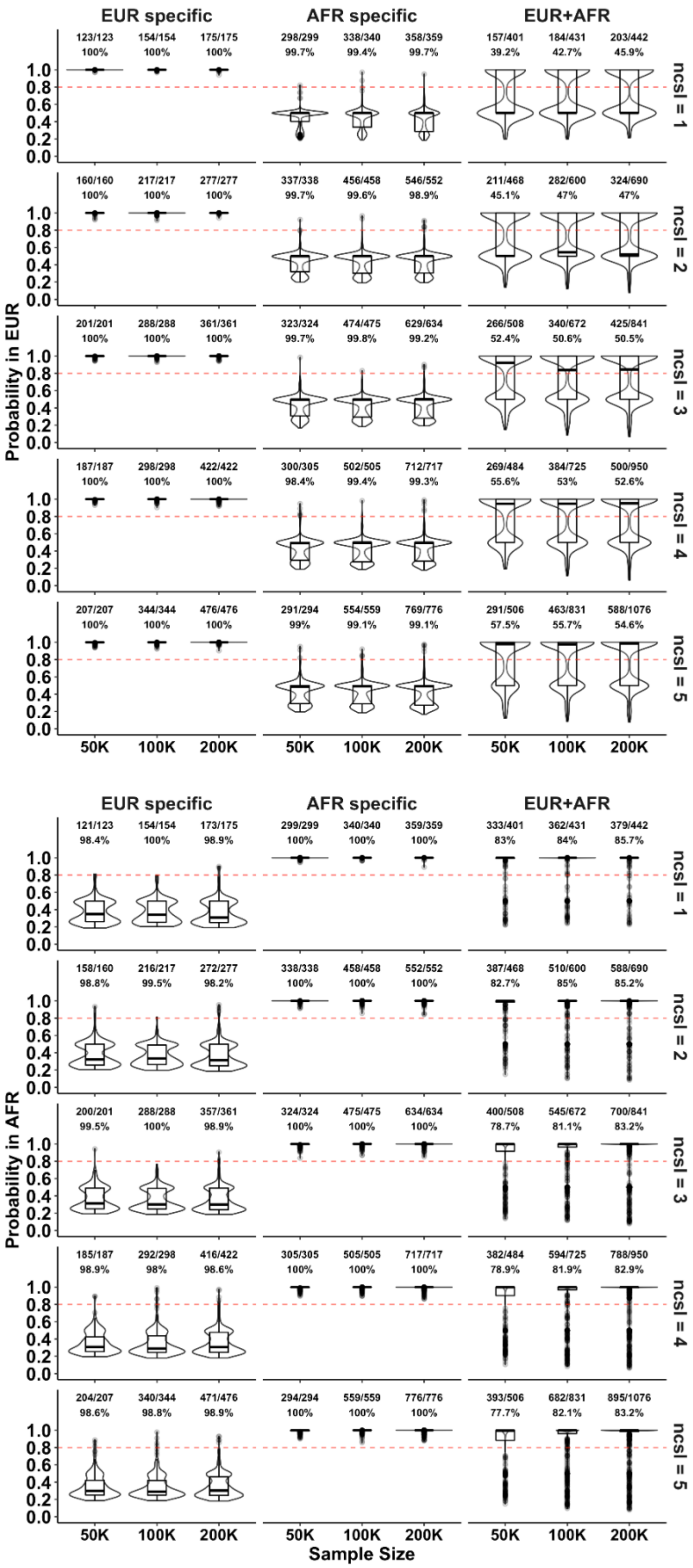
EUR specific
AFR specific
EUR+AFR
123/123 154/154 175/175 298/299 338/340 358/359 157/401 184/431 203/442
100% 100% 100% 99.7% 99.4% 99.7% 39.2% 42.7% 45.9%
ncsl = 1
160/160 217/217 277/277 337/338 456/458 546/552 211/468 282/600 324/690
100% 100% 100% 99.7% 99.6% 98.9% 45.1% 47% 47%
ncsl = 2
201/201 288/288 361/361 323/324 474/475 629/634 266/508 340/672 425/841
100% 100% 100% 99.7% 99.8% 99.2% 52.4% 50.6% 50.5%
ncsl = 3
187/187 298/298 422/422 300/305 502/505 712/717 269/484 384/725 500/950
100% 100% 100% 98.4% 99.4% 99.3% 55.6% 53% 52.6%
ncsl = 4
207/207 344/344 476/476 291/294 554/559 769/776 291/506 463/831 588/1076
100% 100% 100% 99% 99.1% 99.1% 57.5% 55.7% 54.6%
ncsl = 5
Probability in EUR
1.0 0.8 0.6 0.4 0.2 0.0
50K 100K 200K 50K 100K 200K 50K 100K 200K
EUR specific
AFR specific
EUR+AFR
121/123 154/154 173/175 299/299 340/340 359/359 333/401 362/431 379/442
98.4% 100% 98.9% 100% 100% 100% 83% 84% 85.7%
ncsl = 1
158/160 216/217 272/277 338/338 458/458 552/552 387/468 510/600 588/690
98.8% 99.5% 98.2% 100% 100% 100% 82.7% 85% 85.2%
ncsl = 2
200/201 288/288 357/361 324/324 475/475 634/634 400/508 545/672 700/841
99.5% 100% 98.9% 100% 100% 100% 78.7% 81.1% 83.2%
ncsl = 3
185/187 292/298 416/422 305/305 505/505 717/717 382/484 594/725 788/950
98.9% 98% 98.6% 100% 100% 100% 78.9% 81.9% 82.9%
ncsl = 4
204/207 340/344 471/476 294/294 559/559 776/776 393/506 682/831 895/1076
98.6% 98.8% 98.9% 100% 100% 100% 77.7% 82.1% 83.2%
ncsl = 5
Probability in AFR
1.0 0.8 0.6 0.4 0.2 0.0
50K 100K 200K 50K 100K 200K 50K 100K 200K
Sample Size

**Supplementary Figure 31: The population-specific causal probability under varying numbers of causal SNPs per locus ($n_{csl}$) in two-population fine-mapping analysis.** Analyses were conducted with a balanced EUR and AFR sample size. The x-axis shows the discovery sample size. The y-axis shows the population-specific causal probability of the identified credible set. The red dashed lines represent the probability of 0.8, which is used as a threshold to infer whether an identified credible set is causal in a population. The label at the top of each panel denotes the ground-truth causal configuration: “EUR specific” indicates that the variant is causal in the EUR population but not in the AFR population; “AFR specific” indicates that the variant is causal in the AFR population but not in the EUR population; “EUR+AFR” indicates that the variant is causal in both EUR and AFR populations. The label on the right indicates the number of causal SNPs per locus ($n_{csl}$). The fractional number on each panel represents “the number of credible sets with correct inference” among “the total number of credible sets identified by SuSiEx”. The percentage represents the accuracy of the inference. Numerical results are available in Supplementary Table 16.

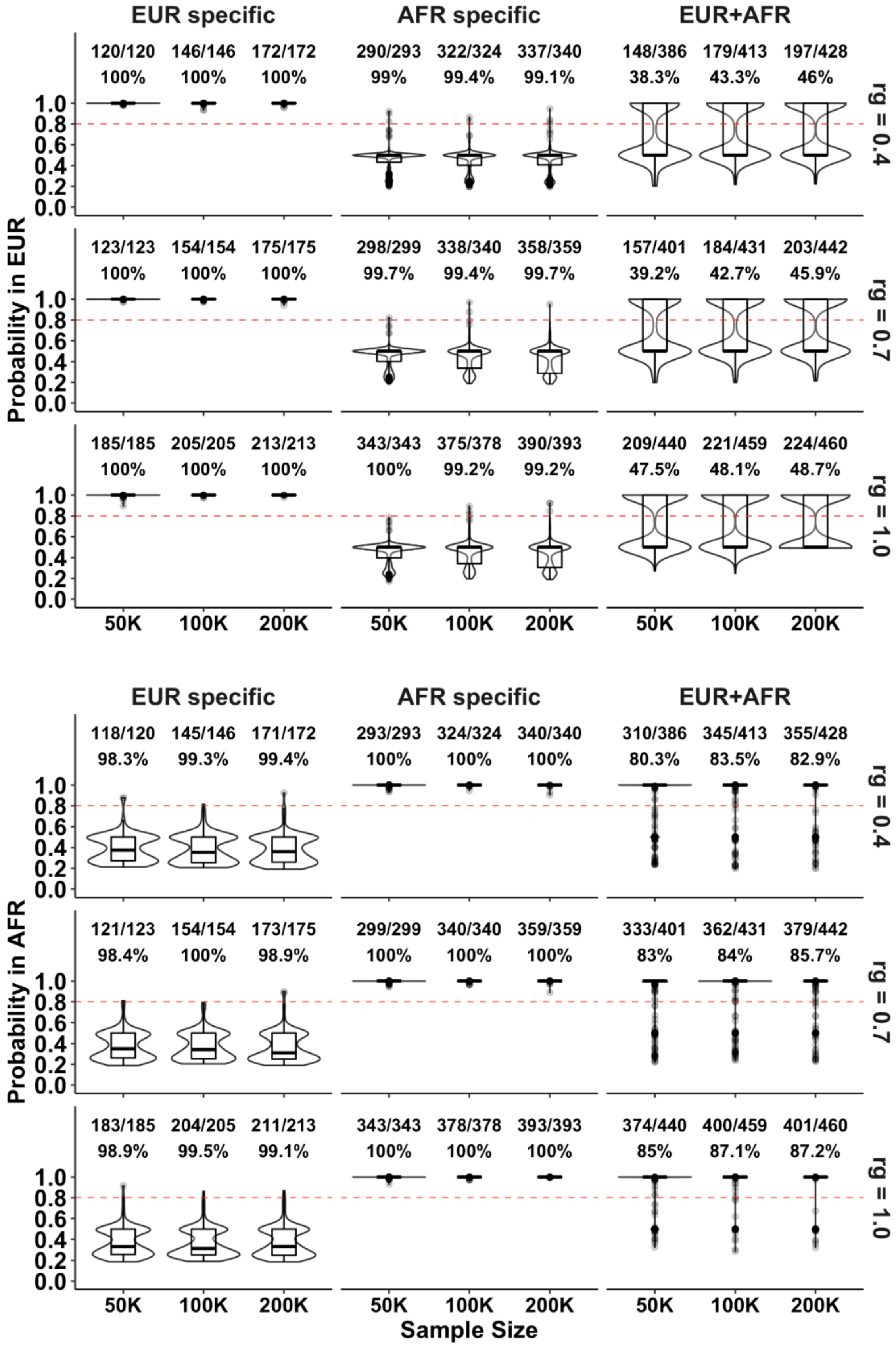
EUR specific
AFR specific
EUR+AFR
120/120 146/146 172/172
100% 100% 100%
290/293 322/324 337/340
99% 99.4% 99.1%
148/386 179/413 197/428
38.3% 43.3% 46%
rg = 0.4
123/123 154/154 175/175
100% 100% 100%
298/299 338/340 358/359
99.7% 99.4% 99.7%
157/401 184/431 203/442
39.2% 42.7% 45.9%
rg = 0.7
185/185 205/205 213/213
100% 100% 100%
343/343 375/378 390/393
100% 99.2% 99.2%
209/440 221/459 224/460
47.5% 48.1% 48.7%
rg = 1.0
Probability in EUR
1.0 0.8 0.6 0.4 0.2 0.0
50K 100K 200K
EUR specific
AFR specific
EUR+AFR
118/120 145/146 171/172
98.3% 99.3% 99.4%
293/293 324/324 340/340
100% 100% 100%
310/386 345/413 355/428
80.3% 83.5% 82.9%
rg = 0.4
121/123 154/154 173/175
98.4% 100% 98.9%
299/299 340/340 359/359
100% 100% 100%
333/401 362/431 379/442
83% 84% 85.7%
rg = 0.7
183/185 204/205 211/213
98.9% 99.5% 99.1%
343/343 378/378 393/393
100% 100% 100%
374/440 400/459 401/460
85% 87.1% 87.2%
rg = 1.0
Probability in AFR
1.0 0.8 0.6 0.4 0.2 0.0
50K 100K 200K
Sample Size

**Supplementary Figure 32: The population-specific causal probability under varying genetic correlations (*$r_g$*) in two-population fine-mapping analysis.** Analyses were conducted with a balanced EUR and AFR sample size. The x-axis shows the discovery sample size. The y-axis shows the population-specific causal probability of the identified credible set. The red dashed lines denote the probability of 0.8, which is used as a threshold to infer whether an identified credible set is causal in a population. The label at the top of each panel denotes the ground-truth causal configuration: “EUR specific” indicates that the variant is causal in the EUR population but not in the AFR population; “AFR specific” indicates that the variant is causal in the AFR population but not in the EUR population; “EUR+AFR” indicates that the variant is causal in both EUR and AFR populations. The label on the right indicates the cross-population genetic correlation ($r_g$). The fractional number on each panel represents “the number of credible sets with correct inference” among “the total number of credible sets identified by SuSiEx”. The percentage represents the accuracy of the inference. Numerical results are available in Supplementary Table 16.

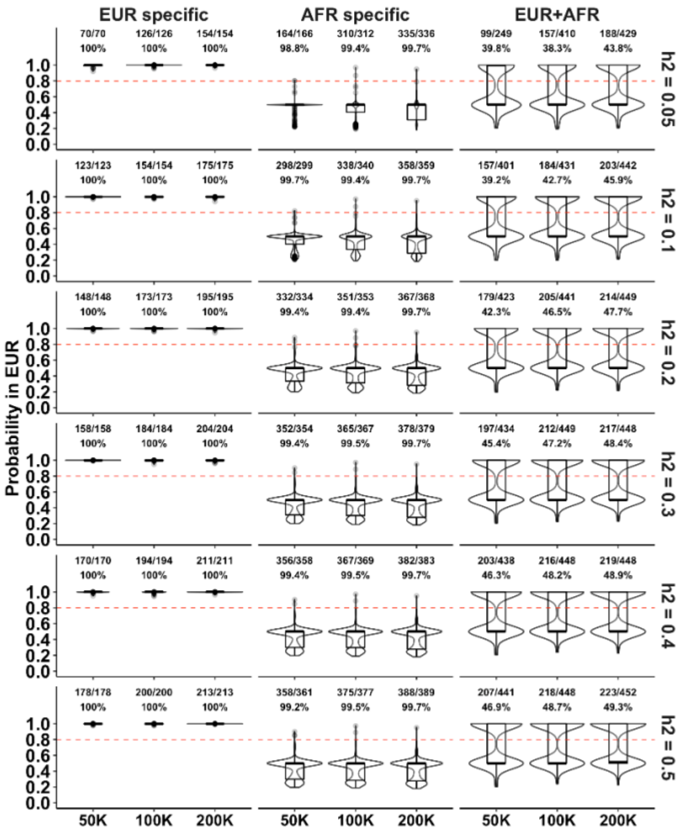
EUR specific
AFR specific
EUR+AFR
70/70 100%
126/126 100%
154/154 100%
164/166 98.8%
310/312 99.4%
335/336 99.7%
99/249 39.8%
157/410 38.3%
188/429 43.8%
h2 = 0.05
123/123 100%
154/154 100%
175/175 100%
298/299 99.7%
338/340 99.4%
358/359 99.7%
157/401 39.2%
184/431 42.7%
203/442 45.9%
h2 = 0.1
148/148 100%
173/173 100%
195/195 100%
332/334 99.4%
351/353 99.4%
367/368 99.7%
179/423 42.3%
205/441 46.5%
214/449 47.7%
h2 = 0.2
158/158 100%
184/184 100%
204/204 100%
352/354 99.4%
365/367 99.5%
378/379 99.7%
197/434 45.4%
212/449 47.2%
217/448 48.4%
h2 = 0.3
170/170 100%
194/194 100%
211/211 100%
356/358 99.4%
367/369 99.5%
382/383 99.7%
203/438 46.3%
216/448 48.2%
219/448 48.9%
h2 = 0.4
178/178 100%
200/200 100%
213/213 100%
358/361 99.2%
375/377 99.5%
388/389 99.7%
207/441 46.9%
218/448 48.7%
223/452 49.3%
h2 = 0.5
Probability in EUR
0.0
0.2
0.4
0.6
0.8
1.0
50K
100K
200K

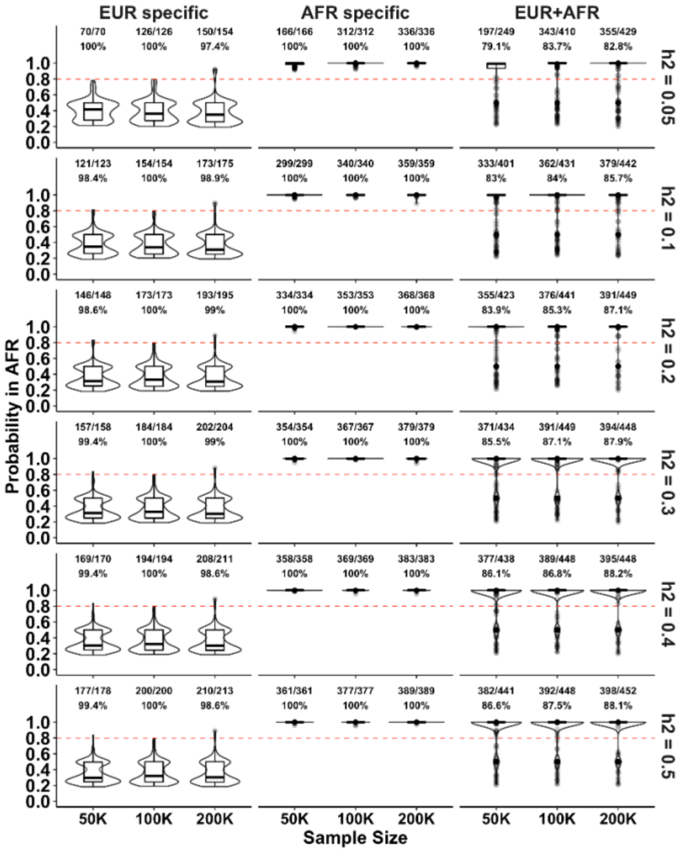
EUR specific
AFR specific
EUR+AFR
70/70 100%
126/126 100%
150/154 97.4%
166/166 100%
312/312 100%
336/336 100%
197/249 79.1%
343/410 83.7%
355/429 82.8%
h2 = 0.05
121/123 98.4%
154/154 100%
173/175 98.9%
299/299 100%
340/340 100%
359/359 100%
333/401 83%
362/431 84%
379/442 85.7%
h2 = 0.1
146/148 98.6%
173/173 100%
193/195 99%
334/334 100%
353/353 100%
368/368 100%
355/423 83.9%
376/441 85.3%
391/449 87.1%
h2 = 0.2
157/158 99.4%
184/184 100%
202/204 99%
354/354 100%
367/367 100%
379/379 100%
371/434 85.5%
391/449 87.1%
394/448 87.9%
h2 = 0.3
169/170 99.4%
194/194 100%
208/211 98.6%
358/358 100%
369/369 100%
383/383 100%
377/438 86.1%
389/448 86.8%
395/448 88.2%
h2 = 0.4
177/178 99.4%
200/200 100%
210/213 98.6%
361/361 100%
377/377 100%
389/389 100%
382/441 86.6%
392/448 87.5%
398/452 88.1%
h2 = 0.5
Probability in AFR
0.0
0.2
0.4
0.6
0.8
1.0
50K
100K
200K
Sample Size

**Supplementary Figure 33: The population-specific causal probability under varying local heritability ($h^2$) in two-population fine-mapping analysis.** Analyses were conducted with a balanced EUR and AFR sample size. The x-axis shows the discovery sample size. The y-axis shows the population-specific causal probability of the identified credible set. The red dashed lines denote the probability of 0.8, which is used as a threshold to infer whether an identified credible set is causal in a population. The label at the top of each panel denotes the ground-truth causal configuration: "EUR specific" indicates that the variant is causal in the EUR population but not in the AFR population; "AFR specific" indicates that the variant is causal in the AFR population but not in the EUR population; "EUR+AFR" indicates that the variant is causal in both EUR and AFR populations. The label on the right indicates the local heritability ($h^2$). The fractional number on each panel represents "the number of credible sets with correct inference" among "the total number of credible sets identified by SuSiEx". The percentage represents the accuracy of the inference. Numerical results are available in Supplementary Table 16.

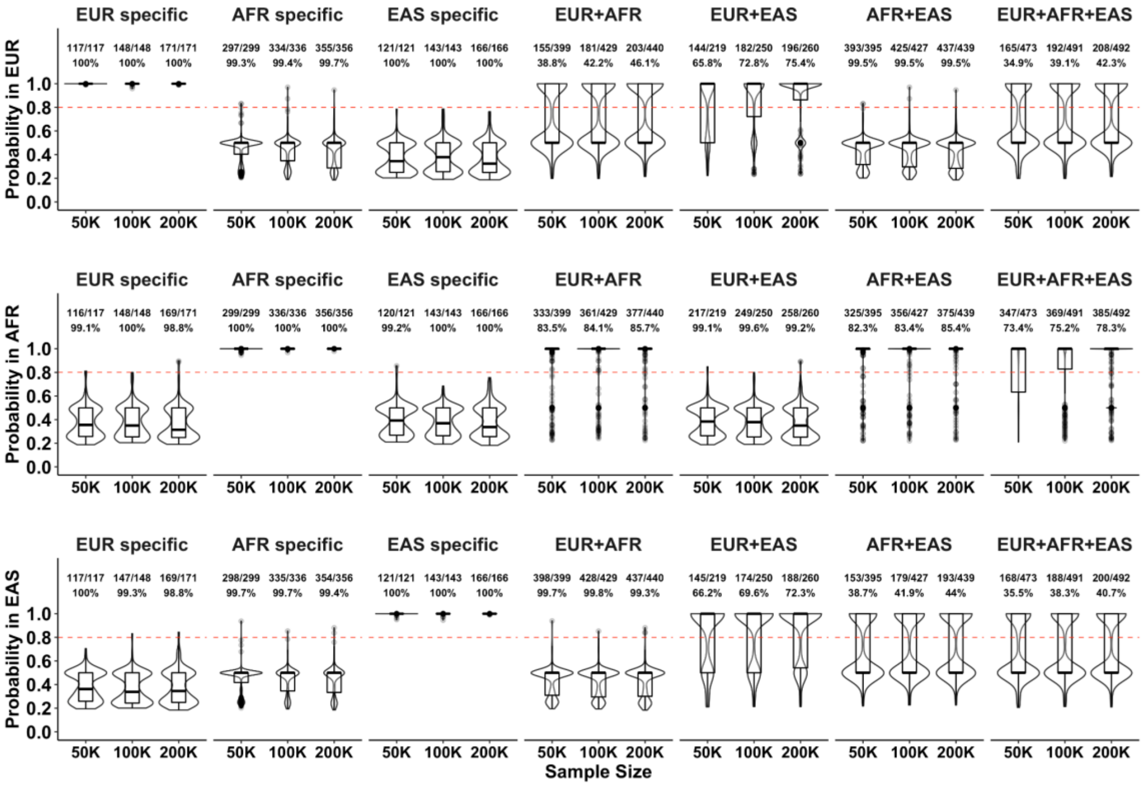


**Supplementary Figure 34: The population-specific causal probability under the standard simulation setting in three-population fine-mapping analysis.** Analyses were conducted with a balanced EUR, AFR and EAS sample size. The x-axis shows the discovery sample size. The y-axis shows the population-specific causal probability of the identified credible set. The red dashed lines denote the probability of 0.8, which is used as a threshold to infer whether an identified credible set is causal in a population. The label at the top of each panel denotes the ground-truth causal configuration: "EUR specific" indicates that the variant is causal in the EUR population but not in the AFR and EAS populations; "AFR specific" indicates that the variant is causal in the AFR population but not in the EUR and EAS populations; "EAS specific" indicates that the variant is causal in the EAS population but not in the EUR and AFR populations; "EUR+AFR+EAS" indicates that the variant is causal in EUR, AFR and EAS populations. The fractional number on each panel represents "the number of credible sets with correct inference" among "the total number of credible sets identified by SuSiEx". The percentage represents the accuracy of the inference. Numerical results are available in Supplementary Table 17.

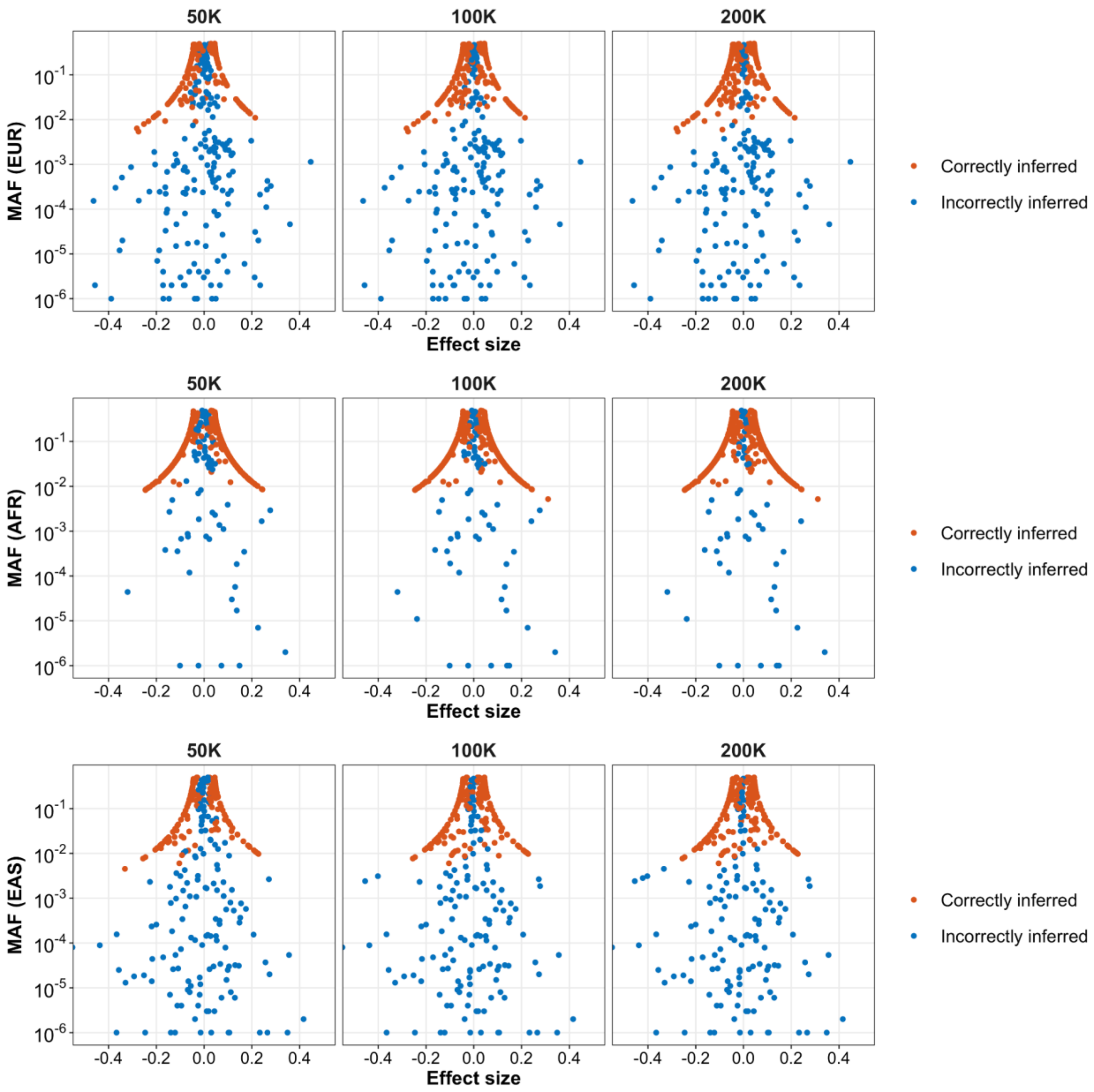


**Supplementary Figure 35: The impact of allele frequency and causal effect size on the classification of population-specific causal variants in three-population fine-mapping analysis.** This plot corresponds to the simulation under the "EUR+AFR+EAS" scenario in Supplementary Figure 34. The x-axis shows the simulated effect sizes of the causal variants. The y-axis shows the minor allele frequencies (MAF) of the causal variants in a specific population. The label above each panel indicates the discovery sample size. Red dots represent variants that are correctly inferred to be causal in the population. Blue dots represent variants that are incorrectly inferred to be not causal in the population. Incorrectly inferred variants tend to have low MAF or small effect sizes. Numerical results are available in Supplementary Table 17.

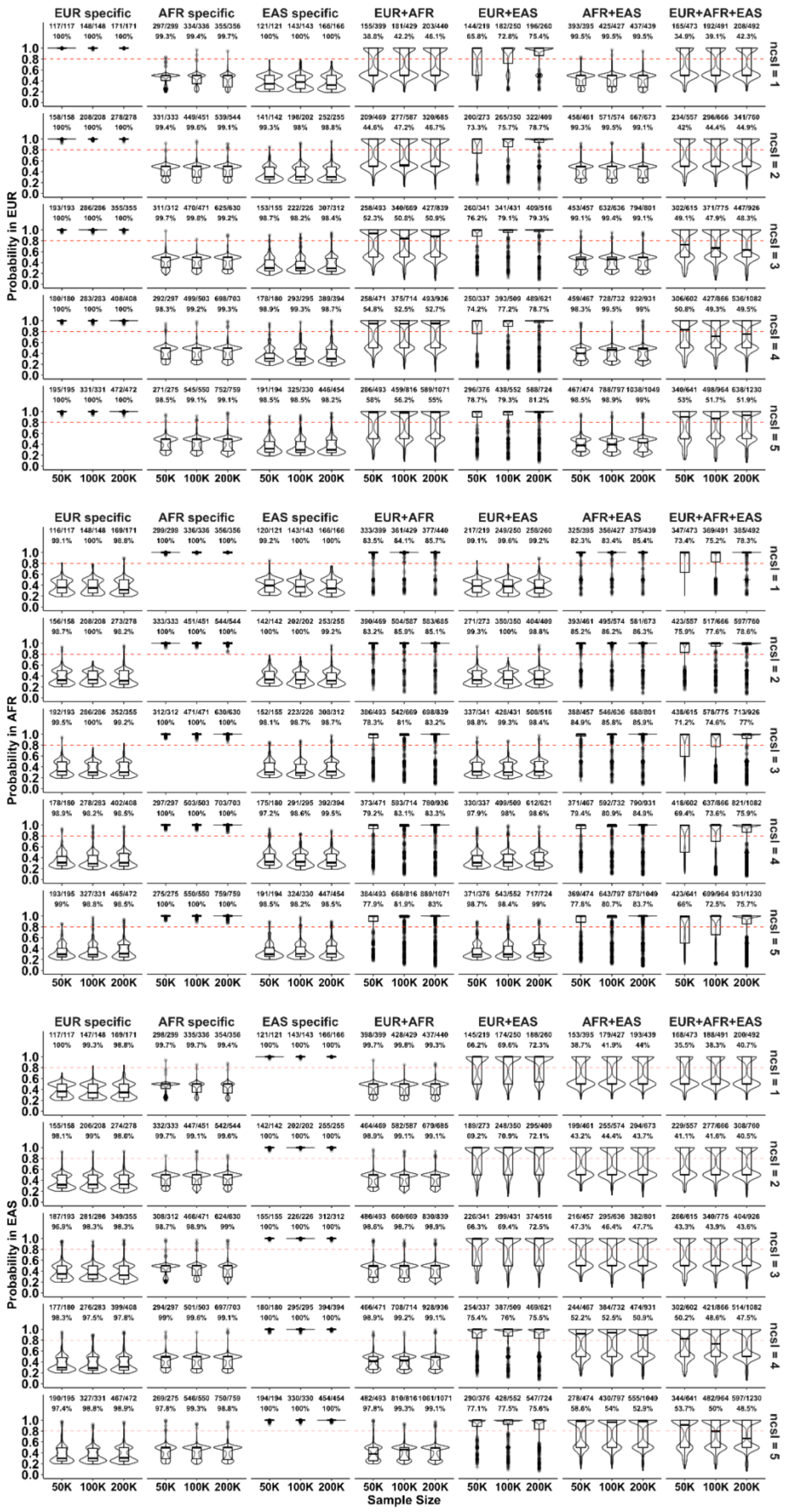

EUR specific
AFR specific
EAS specific
EUR+AFR
EUR+EAS
AFR+EAS
EUR+AFR+EAS
Probability in EUR
Probability in AFR
Probability in EAS
ncsl = 1
ncsl = 2
ncsl = 3
ncsl = 4
ncsl = 5
50K 100K 200K
Sample Size

**Supplementary Figure 36: The population-specific causal probability under varying numbers of causal SNPs per locus ($n_{csl}$) in three-population fine-mapping analysis.** Analyses were conducted with a balanced EUR, AFR and EAS sample size. The x-axis shows the discovery sample size. The y-axis shows the population-specific causal probability of the identified credible set. The red dashed lines denote the probability of 0.8, which is used as a threshold to infer whether an identified credible set is causal in a population. The label at the top of each panel denotes the ground-truth causal configuration: "EUR specific" indicates that the variant is causal in the EUR population but not in the AFR and EAS populations; "AFR specific" indicates that the variant is causal in the AFR population but not in the EUR and EAS populations; "EAS specific" indicates that the variant is causal in the EAS population but not in the EUR and AFR populations; "EUR+AFR+EAS" indicates that the variant is causal in EUR, AFR and EAS populations. The label on the right indicates the number of causal SNPs per locus ($n_{csl}$). The fractional number on each panel represents "the number of credible sets with correct inference" among "the total number of credible sets identified by SuSiEx". The percentage represents the accuracy of the inference. Numerical results are available in Supplementary Table 17.

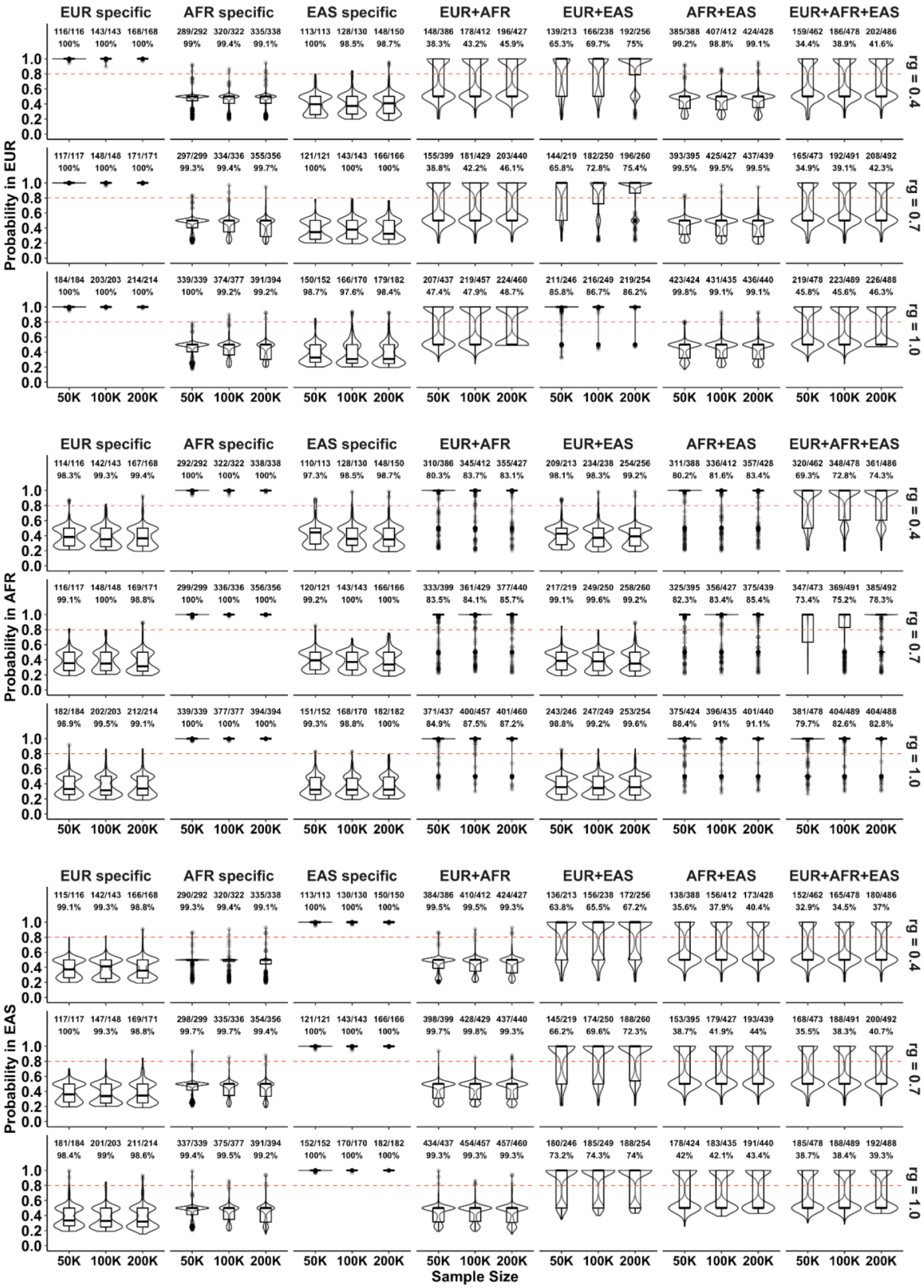

**Supplementary Figure 37: The population-specific causal probability under varying genetic correlations ($r_g$) in three-population fine-mapping analysis.** Analyses were conducted with a balanced EUR, AFR and EAS sample size. The x-axis shows the discovery sample size. The y-axis shows the population-specific causal probability of the identified credible set. The red dashed lines denote the probability of 0.8, which is used as a threshold to infer whether an identified credible set is causal in a population. The label at the top of each panel denotes the ground-truth causal configuration: “EUR specific” indicates that the variant is causal in the EUR population but not in the AFR and EAS populations; “AFR specific” indicates that the variant is causal in the AFR population but not in the EUR and EAS populations; “EAS specific” indicates that the variant is causal in the EAS population but not in the EUR and AFR populations; “EUR+AFR+EAS” indicates that the variant is causal in EUR, AFR and EAS populations. The label on the right indicates the cross-population genetic correlation ($r_g$). The fractional number on each panel represents “the number of credible sets with correct inference” among “the total number of credible sets identified by SuSiEx”. The percentage represents the accuracy of the inference. Numerical results are available in Supplementary Table 17.

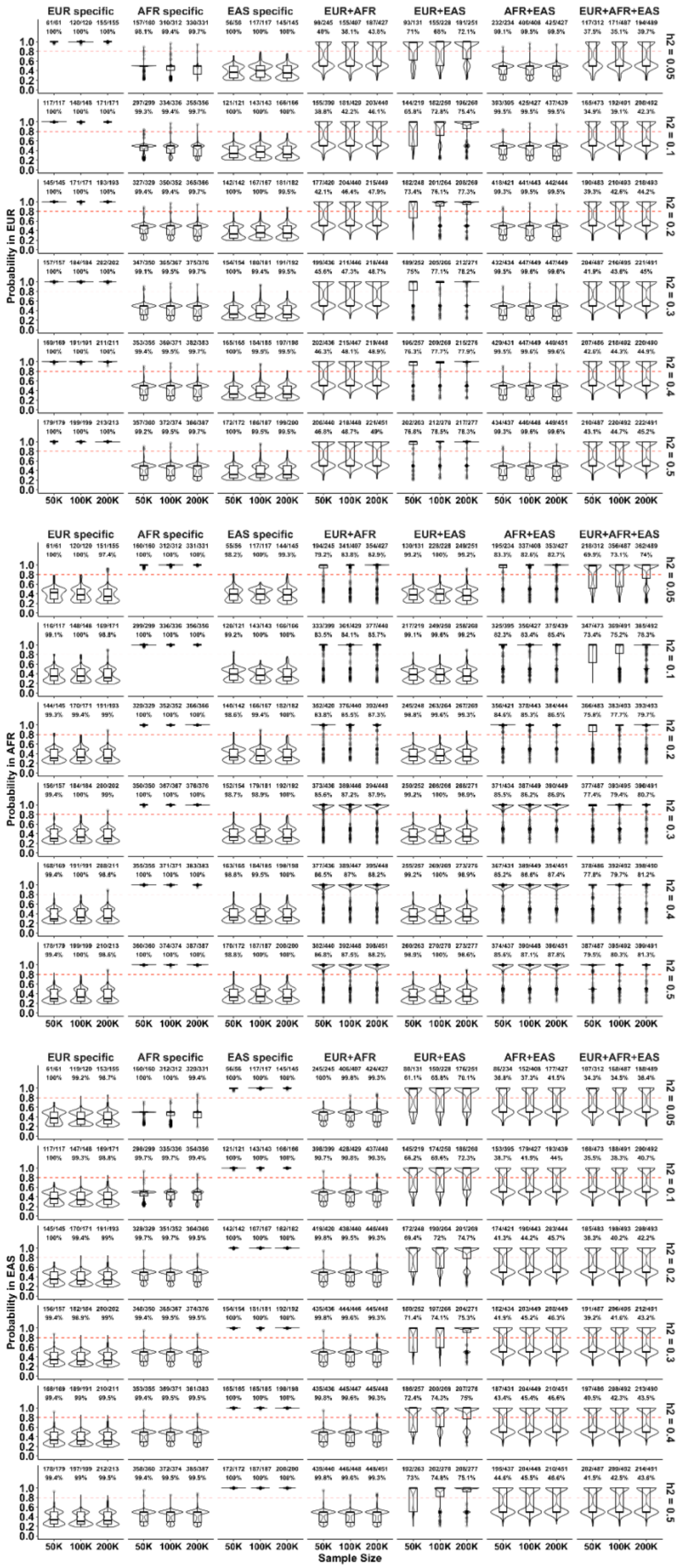
EUR specific
AFR specific
EAS specific
EUR+AFR
EUR+EAS
AFR+EAS
EUR+AFR+EAS
Probability in EUR
Probability in AFR
Probability in EAS
h2 = 0.05
h2 = 0.1
h2 = 0.2
h2 = 0.3
h2 = 0.4
h2 = 0.5
50K 100K 200K
Sample Size

**Supplementary Figure 38: The population-specific causal probability under varying local heritability ($h^2$) in three population fine-mapping analysis.** Analyses were conducted with a balanced EUR, AFR and EAS sample size. The x-axis shows the discovery sample size. The y-axis shows the population-specific causal probability of the identified credible set. The red dashed lines denote the probability of 0.8, which is used as a threshold to infer whether an identified credible set is causal in a population. The label at the top of each panel denotes the ground-truth causal configuration: “EUR specific” indicates that the variant is causal in the EUR population but not in the AFR and EAS populations; “AFR specific” indicates that the variant is causal in the AFR population but not in the EUR and EAS populations; “EAS specific” indicates that the variant is causal in the EAS population but not in the EUR and AFR populations; “EUR+AFR+EAS” indicates that the variant is causal in EUR, AFR and EAS populations. The label on the right indicates the local heritability ($h^2$). The fractional number on each panel represents “the number of credible sets with correct inference” among “the total number of credible sets identified by SuSiEx”. The percentage represents the accuracy of the inference. Numerical results are available in Supplementary Table 17.

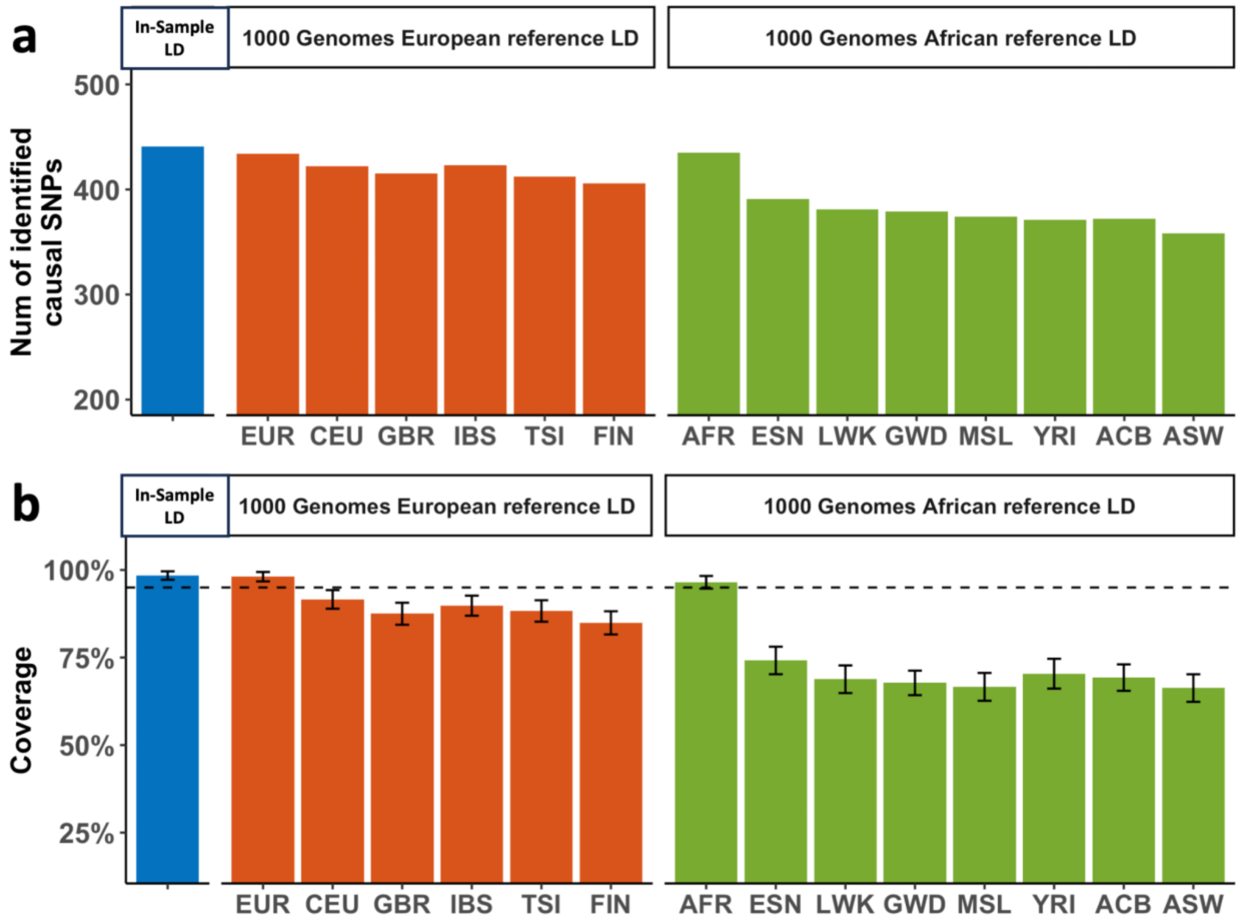


**Supplementary Figure 39: Comparison of fine-mapping analyses between in-sample LD and external reference LD.** Simulation was performed using the standard simulation setting with 200K EUR and 200K AFR samples. **a**, The number of identified true causal variants (true causal variants covered by a credible set). **b**, The coverage of credible sets. The dashed line indicates 95% coverage. The error bar indicates the 95% confidence interval. The top label of each subpanel indicates the reference panel used in the analysis. In-sample LD indicates that in-sample LD was used for both EUR and AFR samples. 1000 Genomes European reference LD indicates that reference LD from 1000 Genomes EUR subpopulations (CEU, GBR, IBS, TSI, FIN) was used for EUR samples but in-sample LD was used for AFR samples. 1000 Genomes African reference LD indicates that reference LD from 1000 Genomes AFR subpopulations (ESN, LWK, GWD, MSL, YRI, ACB, ASW) was used for AFR samples but in-sample LD was used for EUR samples. Numerical results are available in Supplementary Table 18.

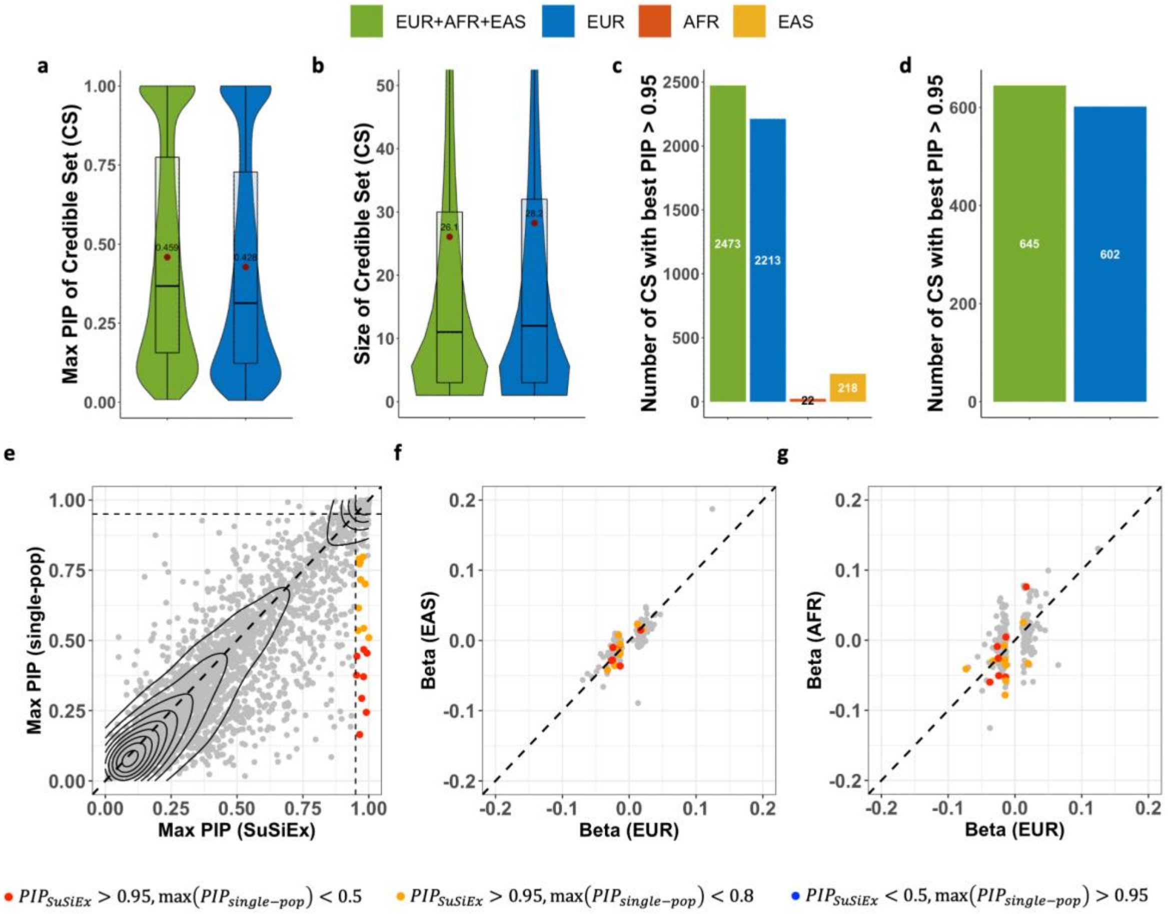


**Supplementary Figure 40: Cross-population fine-mapping analysis after removing the variants with quality issues in biobanks. a**, The distribution of the maximum PIP across 99% credible sets. **b**, The distribution of the size of 99% credible sets. **c**, The number of variants mapped to PIP >95% across 99% credible sets. **d**, The number of variants mapped to PIP >95% in single-credible-set loci. **e**, The maximum PIP from SuSiEx versus the maximum value of the maximum PIP in the three single-population fine-mapping using SuSiE. Only genomic loci with a single credible set aligned across analyses were included. **f** and **g**, The marginal per-allele effect size of the maximum PIP variant in EUR vs. EAS and EUR vs. AFR populations. Variants in single-credible-set loci with PIP >95% estimated by SuSiEx and minor allele frequencies >5% in all populations were included. In **a-b**, red dots represent the mean, the middle line in the box represents the median, and the upper and lower bounds of the box represent the 75th and 25th percentiles, respectively.

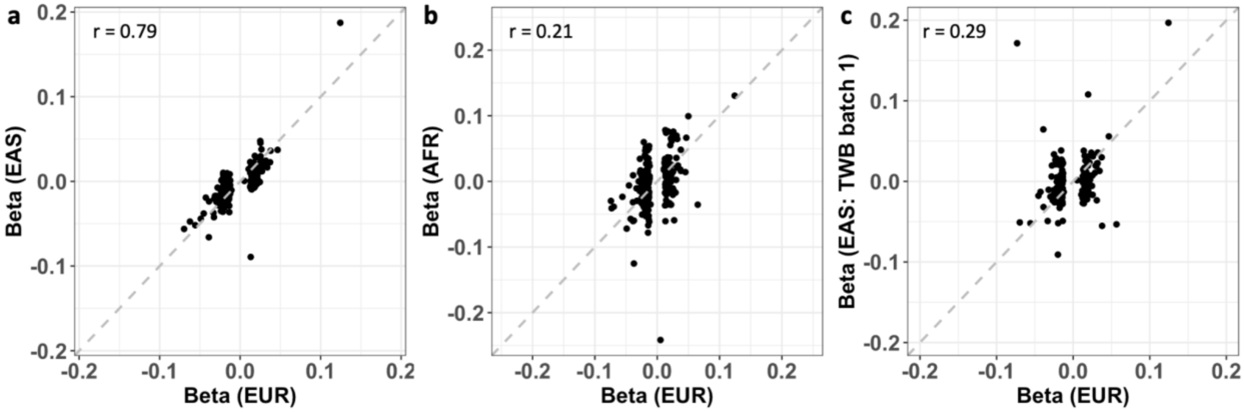


**Supplementary Figure 41: The marginal per-allele effect size of the maximum PIP variant across populations.** We included variants in single-credible-set loci with PIP >95% estimated by SuSiEx and minor allele frequencies >5% in all populations. **a**, EUR vs. EAS; **b**, EUR vs. AFR; **c**, EUR vs. EAS (TWB batch 1).
